# Virologic, clinical, and immunological characteristics of a dengue virus 3 human challenge model

**DOI:** 10.1101/2022.10.24.22281454

**Authors:** Adam T. Waickman, Krista Newell, Joseph Q. Lu, HengSheng Fang, Mitchell Waldran, Chad Gebo, Jeffrey R. Currier, Heather Friberg, Richard G. Jarman, Michelle D. Klick, Lisa A. Ware, Timothy P. Endy, Stephen J. Thomas

## Abstract

Dengue human infection models present an opportunity to explore a vaccine, antiviral, or immuno-compound’s potential for clinical benefit in a controlled setting. Herein, we report the outcome of a phase 1, open-label assessment of a DENV-3 challenge model. In this study, 9 participants received a subcutaneous inoculation with 0.5ml of a 1.4×10^3^ PFU/ml suspension of the DENV-3 strain CH53489. All subjects developed RNAemia within 7 days of inoculation, with peak titers ranging from 3.13×10^4^ to 7.02×10^8^ GE/ml. Symptoms and clinical lab abnormalities consistent with mild dengue infection were observed in all subjects. DENV-3 specific seroconversion was observed by 14 days after inoculation, along with DENV-3 specific memory T cell responses. RNAseq and serum cytokine analysis revealed the presence of an antiviral transcriptional and cytokine response to infection that overlapped with the period of viremia. The magnitude and frequency of clinical and immunologic endpoints correlated with an individual’s peak viral titer.

## INTRODUCTION

Dengue is an expanding global public health threat in predominantly tropical and subtropical regions [1, 2]. Dengue is caused by infection with any of the four DENV types; each being virologically related but genetically and antigenically distinct [3]. *Aedes* mosquito species are the primary vectors of DENV transmission and changes in climate have expanded their habitat allowing for local transmission in historically more temperate regions [4, 5].

It is estimated there are up to 400 million DENV infections every year, with more than 90 million resulting in clinically relevant disease [1, 2, 6]. Infection may result in an asymptomatic or subclinical infection or classic dengue fever [3, 7]. Severe dengue occurs in a minority of infections, approximately 2-4% of people experiencing a sequential infection with a different DENV type more than 18 months following their first infection [8-10]. Severe dengue may manifest as plasma leakage, hemorrhage, end organ dysfunction, and death [8, 9, 11]. Current treatment is primarily based on symptom relief and judicious intravascular volume replacement with fluids [12, 13].

Currently, there are no antivirals approved for the prevention or treatment of DENV infections. There are also no immuno-compounds (i.e., monoclonal antibodies) approved for the prevention or treatment of dengue. A single dengue vaccine (Sanofi Pasteur, Dengvaxia^®^) has been approved for use in numerous countries, but uptake has been low due to the need to assess dengue serostatus prior to vaccination, and an observed safety signal in children who were seronegative at the time of vaccination [14-16]. Takeda Vaccines has recently received approval for use of their vaccine (TAK-003) in Indonesia without the restriction of needing to confirm serostatus prior to vaccination [17-19]. The company is pursuing additional regulatory approvals.

Developing countermeasures against dengue has been challenging for a number of reasons, including; 1) the existence of four DENVs, each capable of causing disease and death, 2) an incomplete understanding of what constitutes an immuno-protective versus immuno-pathologic profile prior to infection; 3) the inability to precisely measure homotypic, non-cross reactive DENV immune responses; and 4) the absence of an animal model of disease which accurately recapitulates the human infection experience [20, 21].

For these reasons, a consortium of investigators has continued over a century of work devoted to developing a safe dengue human infection model (DHIM) [22]. The intent of the DHIM is to expose healthy volunteers to under-attenuated DENVs and create a mild and uncomplicated dengue illness. The objective is to intensely study signs and symptoms of dengue, clinical lab abnormalities, virologic, and innate and adaptive immune responses. Knowing the exact infecting DENV type and time of exposure allows investigators to explore, in detail, the kinetics of clinical, virologic, serologic, and immunologic responses to infection.

The DHIM is not intended to study moderate or severe dengue, but to drastically improve upon the limitations of current small animal and non-human primate models of uncomplicated dengue disease. A safe and consistently performing DHIM representing each of the four DENV types would be a powerful tool supporting the development of dengue vaccines, treatments, and non-vaccine prophylactics. A DHIM capable of assessing, early in development, the potential for clinical benefit of a countermeasure could prevent thousands of volunteers having to be exposed to ineffective countermeasure candidates.

In this manuscript we describe our clinical characterization of a DENV-3 DHIM viral strain. In this study, we screened and enrolled 9 flavivirus naïve, healthy adults. Each volunteer was subcutaneously inoculated with 0.5ml of a 1.4×10^3^ PFU/ml suspension of the DENV-3 strain CH53489 manufactured under Good Manufacturing Practices. The trial was conducted following U.S. Food and Drug Administration acceptance of an Investigational New Drug application and local Institutional Review Board approval. Following inoculation, volunteers were assessed daily or every other day for 28 days. Follow-up visits also occurred at more remote timepoints out to 180 days. Blood samples were collected to assess safety and virologic, serologic, and immunologic responses to infection.

## RESULTS

### Virologic characteristics of CH53489 strain DENV-3 challenge

Nine participants (**Supplemental Table 1**) were enrolled in this study to receive a single subcutaneous inoculation with 0.5ml of a 1.4×10^3^ PFU/ml suspension of the DENV-3 strain CH53489 [23, 24]. Participants in this study were evaluated daily for the first 14 days after inoculation either in-person or by phone, then every other day until 28 days post virus inoculation with additional visits on days 90 and 180 post infection (**Supplemental Figure 1**). Participants were admitted to SUNY Upstate Medical University Hospital for observation if they met pre-defined hospitalization criteria. Six of nine study participants were admitted for observation during the course of the study. As with previous DENV-1 challenge studies, all participants who became ill were managed with oral fluids, acetaminophen, and an anti-nausea medication if required. No intravenous access was required. In the event of hospitalization, study participants were shifted to an alternative sample collection schedule where safety and research blood draws were collected daily for the duration of the hospital stay and additional research draws collected three- and seven-days post discharge. All nine enrolled participants completed the study per protocol and were included in the subsequent virologic and immunologic analyses.

All study participants developed detectable RNAemia and viremia as assessed by qRT-PCR and Vero cell-based plaque assay, respectively, while 8 of 9 study participants experienced quantifiable NS1 antigenemia (**Figure 1A, Figure. 1B, Figure 1C)**. The incubation period (time from infection to detectable RNAemia) ranged from 3 to 6 days (mean 4.1 days post challenge) (**Figure 1D**), with peak viral loads occurring 6 to 8 days post challenge (mean 6.7 days post challenge) (**Figure 1E**). Peak viral load ranged from 3.13×10^4^ to 7.02×10^8^ genome equivalents (GE)/mL, or 5.3×10^3^ to 3.0×10^7^ plaque forming units (PFU)/mL of serum, with detectible viremia (day of first detection until first day below detection) lasting 4 to 11 days (mean of 7.3 days). DENV NS1 antigenemia was detected on average 3.1 days later than RNAemia or viremia, with the day of detectible NS1 antigenemia ranging between 6 to 12 days post challenge (mean 7.3 days), persisting for an average (mean) of 12.25 days across all NS1 antigen positive study participants (**Fig. 1C**).

**Figure 1.**
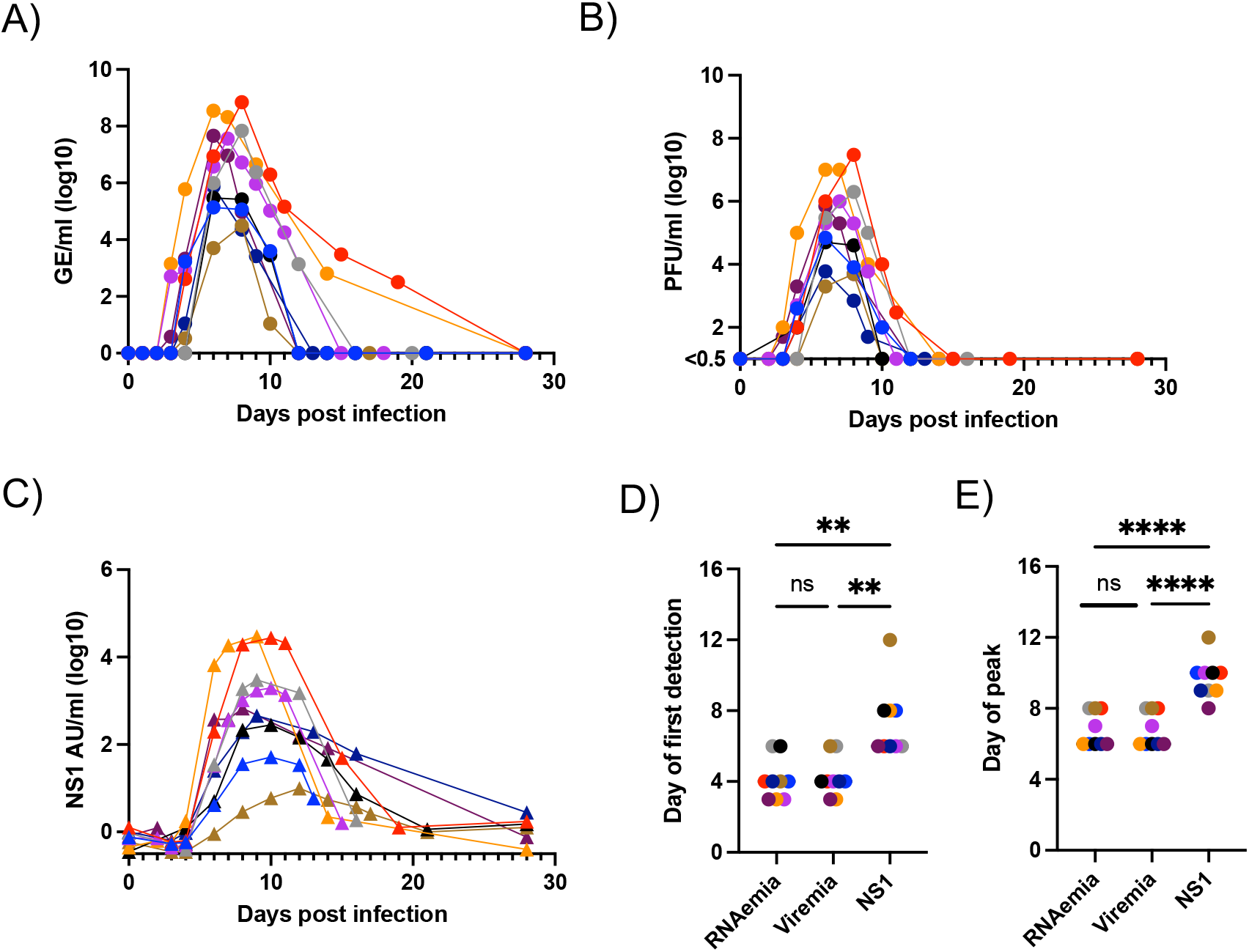
Viral kinetics of CH53489 challenge. **A)** DENV-3 RNA content in serum as assessed by qRT-PCR, **B)** Infectious DENV content in serum as assessed Vero cell plaque assay., **C)** NS1 protein content in serum as assessed by ELISA, **D)** Comparison of RNAemia, viremia, and NS1 antigenemia onset in all study participants, **E)** Comparison of peak RNAemia, viremia, and NS1 antigenemia in all study participants. ns not significant, ** p < 0.01, **** p< 0.0001, paired 1 way ANOVA with correction for multiple comparisons

### Clinical characteristics of CH53489 strain DENV-3 challenge

Within the first 14 days following inoculation 9/9 study participants reported at least one solicited systemic AE. Rash was observed in 7/9 subjects, which was evident for between 3 and 20 days per individual (median of 9 days), with 4/9 reporting severe rash based on the percent of body surface area involved (**Figure 2A**). Fatigue was described by 7/9 subjects, lasting for 3-12 days (median of 6 days) (**Figure 2B**). Headache was described by 8/9 subjects and was experienced for 1 to 11 days per subject (median of 3 days) (**Figure 2C**). Fever – defined as a temperature equal to or greater than 100.4°C - was observed in 8/9 individuals, lasting for 1 to 5 days per subject (median 2 days), with one subject experiencing severe (≥ 102.1°C) fever (**Figure 2D**).

**Figure 2.**
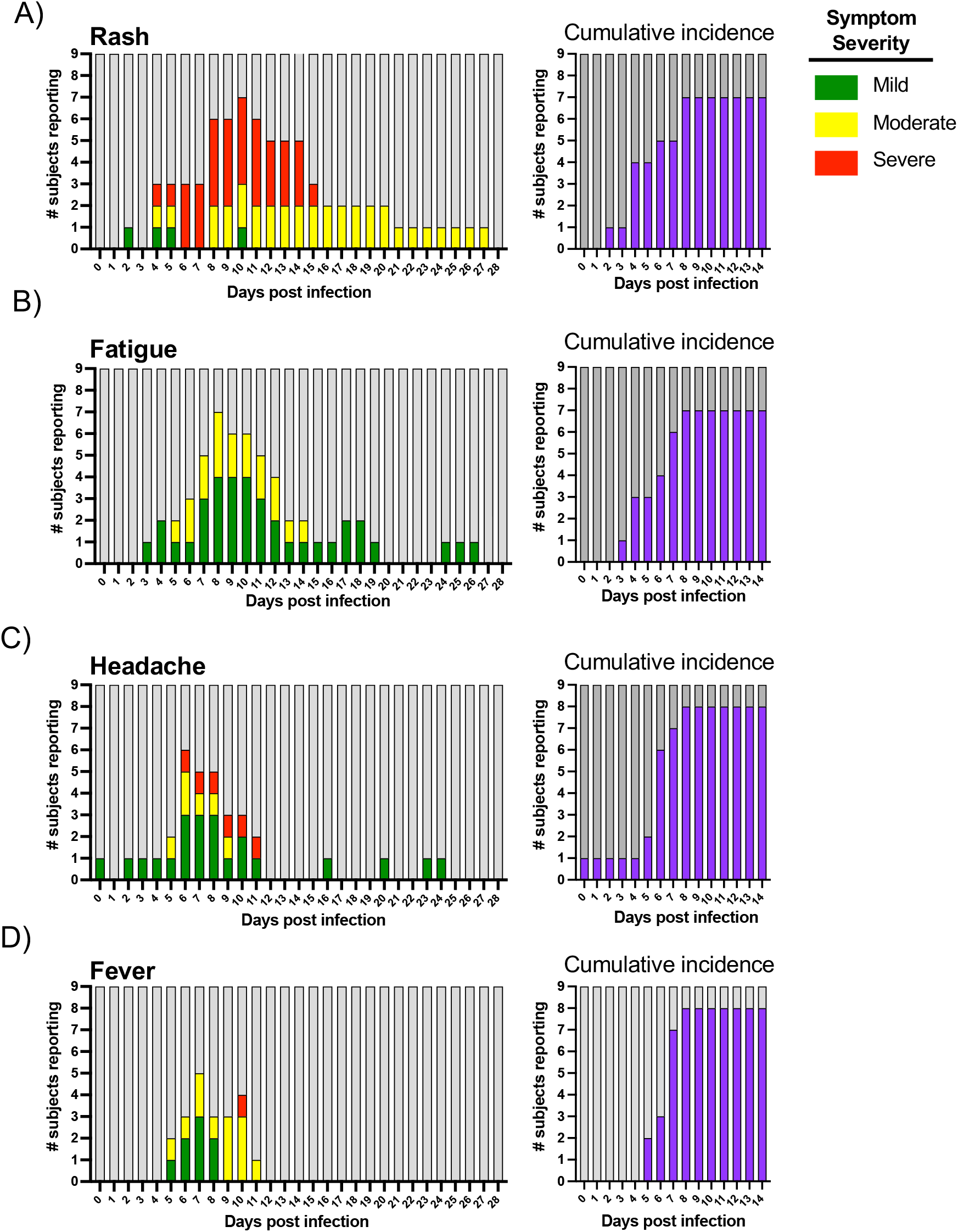
Timing and severity of clinically significant adverse events in response to CH53489 challenge. **A)** Rash, **B)** Fatigue, **C)** Headache, **D)** Fever

All study participants had at least 1 laboratory abnormality between days 1 and 28 post DENV-3 strain CH53489 challenge, the majority of which were mild or moderate. At least 1 severe laboratory abnormality was reported by 5 of the 9 participants. Elevated liver enzyme levels (ALT/AST) were observed in 5/9 study participants with 2 individuals classified as severe and all returning to normal ranges within 30 days post challenge (**Figure 3A, Figure 3B**). No hemoconcentration was observed in any subject during the course of the study (**Figure 3C**). Lymphopenia was observed in 9/9 subjects (3 severe), while neutropenia was observed in 7/9 subjects (2 severe), and thrombocytopenia observed in 4/9 study participants (1 severe) (**Figure 3D, Figure 3E, Figure 3F**).

**Figure 3.**
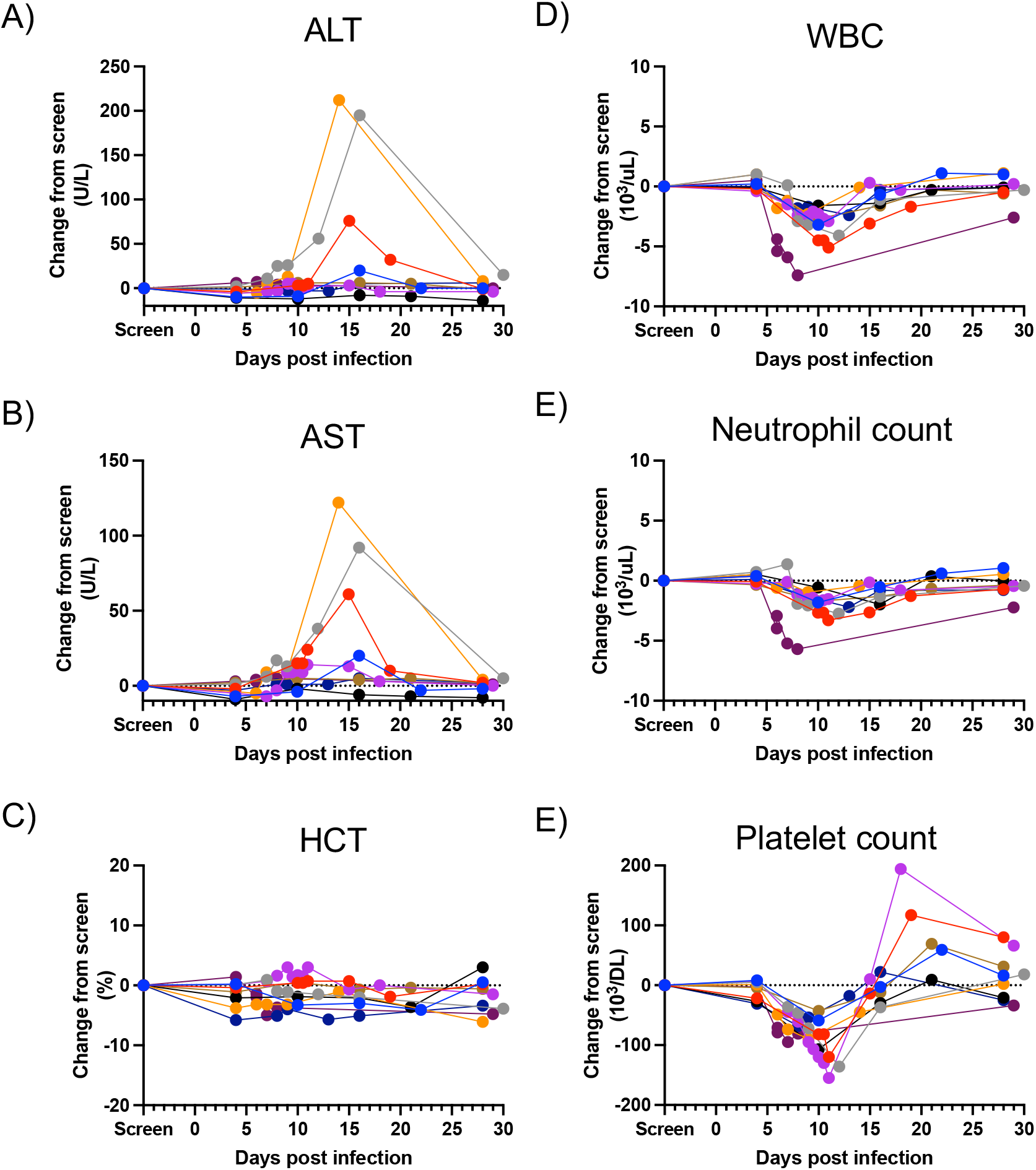
Timing and magnitude of clinical lab abnormalities following CH53489 challenge. **A)** ALT, **B)** AST, **C)** HCT, **D)** WBC, **E)** Neutrophil count, **F)** Platelet count. All results shown relative to screening visit value

### Kinetics and specificity of DENV-3 elicited humoral immunity

Having established the virologic and clinical features of CH53489 human challenge we next assessed the induction of DENV-3 specific adaptive immunity. Using a virion-capture ELISA assay we observed the induction of a robust DENV-3 specific IgM response in all participants first detectible between days 9 and 14 post infection and persisting above baseline in 8/9 subjects for at least 90 days (**Figure 4A, Figure 4B**). A transient DENV-3 specific IgA response was also observed starting between days 12 and 15 post challenge, dropping below the threshold of detection in 7/9 subjects by day 90 post infection (**Figure 4A, Figure 4B)**. DENV-3 specific IgG was detected in circulation observed starting between days 14 and 28 post challenge, remaining above the threshold of detection in 8/9 for at least 90 days post infection (**Figure 4A, Figure 4B)**.

**Figure 4.**
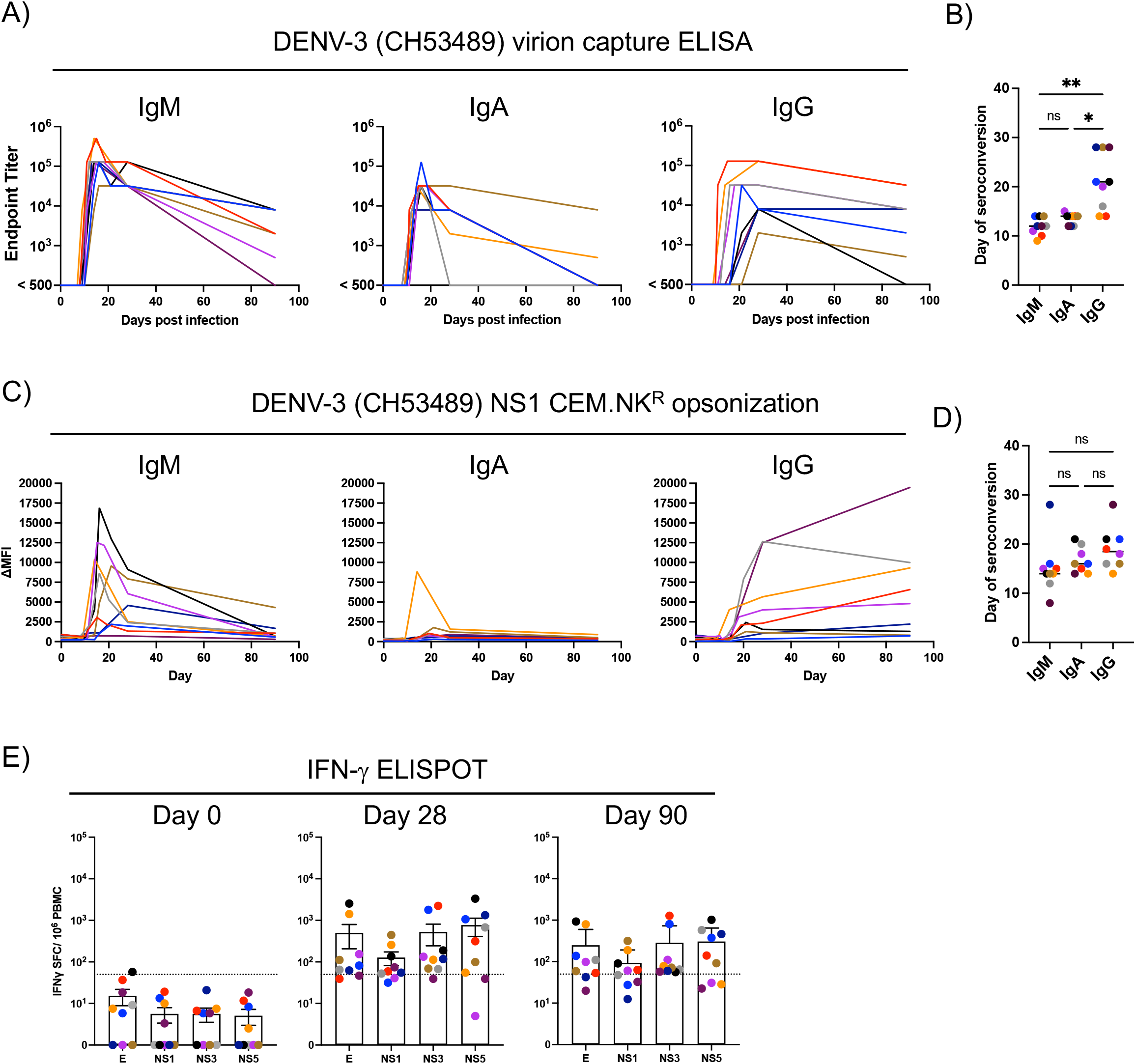
Kinetics and specificity of DENV-3 specific immunity elicited by CH53489 challenge. **A)** Antibody endpoint titers using virion-capture ELISA. **B)** Day of virion-specific IgM, IgA, and IgG seroconversion. **C)** Opsonizing anti-NS1 antibody staining using DENV-2 NS1 expressing CEM.NK^R^ cells **D)** Day of DENV-2 NS1-specific IgM, IgA, and IgG seroconversion. **E)** Antigen specific breakdown of days 0, 28, and 90 DENV-3 specific cellular immunity. Dashed line indicates 50 SFC/10^6^ PBMC.

In addition to assessing the abundance of virion-specific antibodies, the abundance of NS1-specific antibodies elicited by CH53489 infection were quantified using cell-based NS1 opsonization assay. We utilized a CEM.NK^R^ cell line that was engineered to stably express DENV-3 NS1 and quantified the abundance of anti-NS1 opsonizing antibodies using flow cytometry (**Supplemental Figure 3**). Robust induction of anti-NS1 IgM antibodies was observed in all study participants first starting between days 8 and 28 post infection, with IgG isotype responses appearing later and persisting through day 90 post infection (**Figure 4C, Figure 4D**). Only modest and transient anti-NS1 IgA antibody titers were observed following CH53489 challenge (**Figure 4C, Figure 4D**).

Consistent with the robust humoral immune response elicited by CH53489 challenge, IFN-γ ELISPOT analysis of PBMC collected from study participants on days 0, 28 and 90 post infection demonstrated the presence of a significant adaptive cellular immune response to infection. Stimulation with overlapping peptide pools spanning the E, NS1, NS3, and NS5 proteins demonstrated the induction of significant cellular immune responses against all antigens included in the analysis, which NS3, NS5 and E proteins exhibiting higher responses on average than NS1(**Figure 4E, Supplemental Figure 4**). In their totality these results demonstrate that CH53489 challenge results in the development of robust DENV-3 specific humoral and cellular immunity that persists for at least 90 days post infection.

### Transcriptional profiling of CH53489-elicited immune activation

In light of the robust adaptive immune response elicited by CH53489 challenge we next endeavored to define the early transcriptional profile of DENV-3 elicited inflammation. Accordingly, we performed RNAseq analysis on whole blood samples collected on or around days 0, 6, 8, 10, 14 and 28 post CH53489 challenge in all 9 study participants. These time points were selected to capture both the peak of viral replication (days 6-8 post infection) as well as the days when the induction of the adaptive immune response was predicted to occur (day 10-14). Due to the adaptive nature of the study protocol sample collections were not synchronous across all study participants. Samples were binned into the nearest common study day (0, 6, 8, 10, 14, or 28) for subsequent analysis (**Supplemental Table 2**).

Robust transcriptional deviations away from the day 0 transcriptional baseline were observed on days 6, 8, 10, and 14 post infection, with all participants returning to a baseline transcriptional state by day 28 post infection (**Figure 5A**). A total of 205 (196 upregulated, 9 downregulated), 302 (247 upregulated, 55 downregulated), 245 (195 upregulated, 50 downregulated) and 295 (267 upregulated, 28 downregulated) differentially expressed genes were observed on days 6, 8, 10, and 14, receptively (**Figures 5B-E, Supplemental Tables 3-6**). No differentially expressed gene were observed on day 28 relative to day 0. Significant overlap was observed between the differentially expressed genes detected on days 6, 8, and 10, while day 14 was characterized by a mostly independent set of differentially expressed gene products (DEGs) (**Figure 5F, Supplemental Figure 5**).

**Figure 5.**
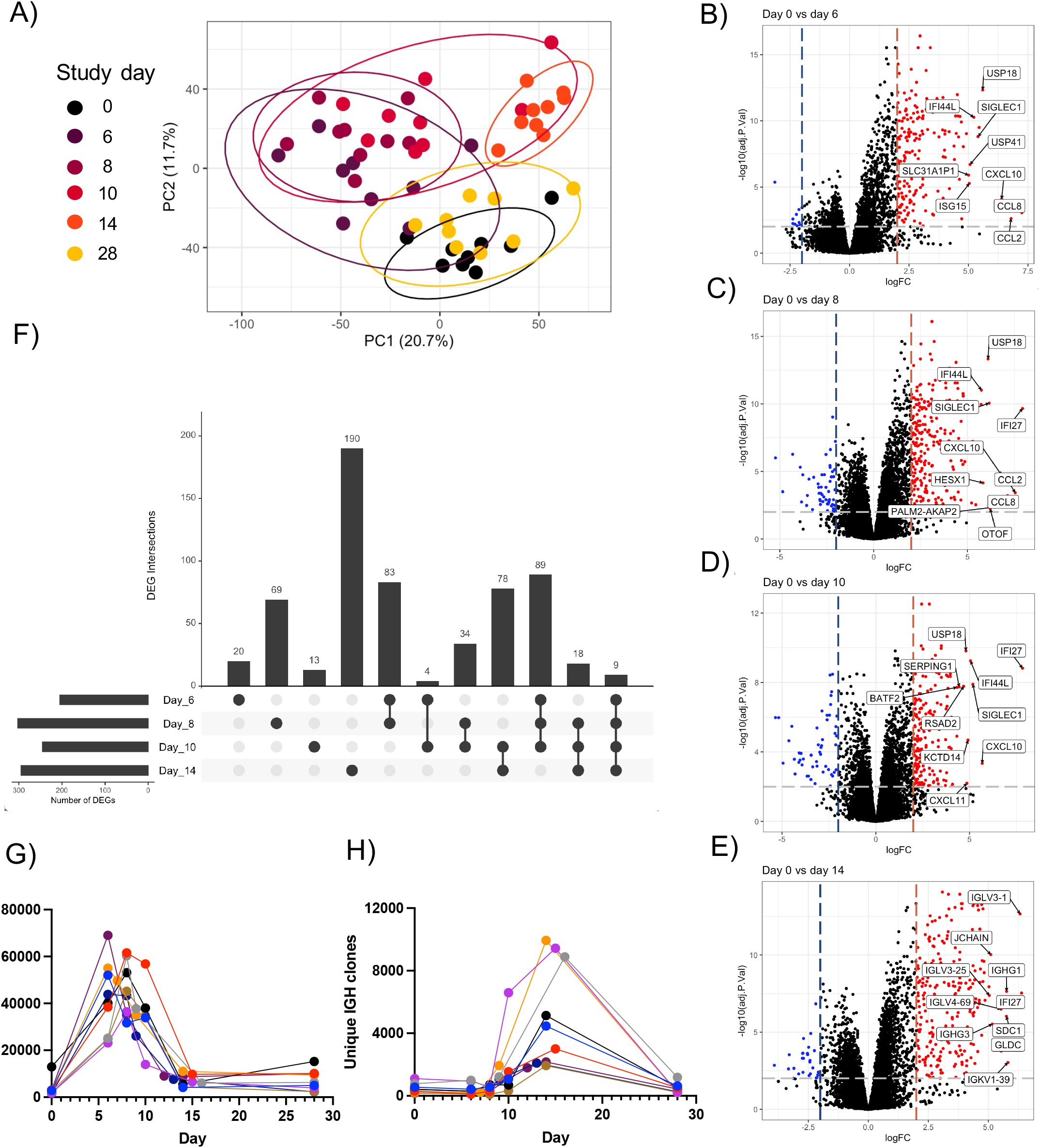
Kinetics and composition of DENV-3 elicited inflammation. **A)** PCA plot of RNAseq analysis of whole blood obtained on days 0, 6, 8, 10, 14, and 28 post CH53489 infection. Points colored by sample collection day. **B)** Volcano plot showing differential gene expression day 0 vs day 6, **C)** day 0 vs day 8, **D)** day 0 vs day 10, and **E)** day 0 vs day 14 post infection with select statistically and biologically significant genes highlighted. Genes with a log_2_ fold change of >2 and an adjusted p-value < 0.01 were considered significant. **F)** Overlap of Differentially Expressed Genes (DEG) observed on days 6, 8, 10 and 14 post CH53489 infection. **G)** Kinetics of previously defined inflammatory gene module expression across all 9 study participants and 6 time points. **H)** Identification and quantification of unique IGH clones across all 9 study participants and 6 time points from the unenriched RNAseq data using MiXCR.

Ingenuity Pathway Analysis (IPA) of the DEGs identified in this study revealed that days 6, 8, and 10 were dominated by the expression of gene products associated with interferon signaling, chemokine/cytokine production, and the induction of antiviral responses (**Table 1, Supplemental Figure 6, Supplemental Tables 7-9**). Gene pathways consistent with nascent B cell activation and antibody production were also observed on days 10 and 14 post DENV-3 infection, with the day 14 gene profile dominated by terms associated with mitotic cell division and lymphocyte proliferation (**Table 1, Supplemental Figure 6, Supplemental Tables 9-10**). Consistent with these transcriptional profiles, elevated levels of IL-1Ra, IL-8, IFN-β, IL-10, and IFN-α2a were observed in serum collected from subjects on days 6, 8 and 10 post challenge (**Supplemental Figure 7**), while IFN-γ, IL-10, and IL-9 were additionally detected starting on day 10 post infection (**Supplemental Figure 8**). No statistically significant differences in VEGFA, TNFα, MIP-1a, GM-CSF, MCP1, IL-1β, IL-12p70, G-CSF, IL-6, IL-4 or IL-5 levels were observed (**Supplemental Figure 9**).

**Table 1.** IPA analysis of differentially expressed genes defined by RNAseq on days 6, 8, 10, and 14 post CH58349 infection.

| Comparison | Ingenuity Canonical Pathways | -log(p-value) | Ratio | z-score |
| --- | --- | --- | --- | --- |
| Day 0 vs Day 6 | Role of Hypercytokinemia/hyperchemokine in the Pathogenesis of Influenza | 1.98E+01 | 1.98E-01 | 3.638 |
| Day 0 vs Day 6 | Interferon Signaling | 1.71E+01 | 3.33E-01 | 2.887 |
| Day 0 vs Day 6 | Activation of IRF by Cytosolic Pattern Recognition Receptors | 9.22E+00 | 1.38E-01 | 1.667 |
| Day 0 vs Day 6 | Role of Pattern Recognition Receptors in Recognition of Bacteria and Viruses | 7.97E+00 | 7.05E-02 | 2 |
| Day 0 vs Day 6 | Coronavirus Pathogenesis Pathway | 6.80E+00 | 5.42E-02 | -2.111 |
| Day 0 vs Day 6 | Role of PKR in Interferon Induction and Antiviral Response | 5.35E+00 | 5.88E-02 | 2.646 |
| Day 0 vs Day 6 | Granulocyte Adhesion and Diapedesis | 4.31E+00 | 4.23E-02 | NaN |
| Day 0 vs Day 6 | Death Receptor Signaling | 4.29E+00 | 6.25E-02 | 2.449 |
| Day 0 vs Day 6 | Retinoic acid Mediated Apoptosis Signaling | 4.25E+00 | 8.33E-02 | 2.236 |
| Day 0 vs Day 6 | Agranulocyte Adhesion and Diapedesis | 3.94E+00 | 3.74E-02 | NaN |
| Day 0 vs Day 8 | Role of Hypercytokinemia/hyperchemokine in the Pathogenesis of Influenza | 1.70E+01 | 1.98E-01 | 4.123 |
| Day 0 vs Day 8 | Interferon Signaling | 1.35E+01 | 3.06E-01 | 2.714 |
| Day 0 vs Day 8 | Role of Pattern Recognition Receptors in Recognition of Bacteria and Viruses | 1.03E+01 | 9.62E-02 | 2.828 |
| Day 0 vs Day 8 | Activation of IRF by Cytosolic Pattern Recognition Receptors | 7.81E+00 | 1.38E-01 | 1.667 |
| Day 0 vs Day 8 | Granulocyte Adhesion and Diapedesis | 6.36E+00 | 6.35E-02 | NaN |
| Day 0 vs Day 8 | Coronavirus Pathogenesis Pathway | 6.03E+00 | 5.91E-02 | -2.111 |
| Day 0 vs Day 8 | Complement System | 4.51E+00 | 1.35E-01 | 1 |
| Day 0 vs Day 8 | Agranulocyte Adhesion and Diapedesis | 4.24E+00 | 4.67E-02 | NaN |
| Day 0 vs Day 8 | Role of PKR in Interferon Induction and Antiviral Response | 4.19E+00 | 5.88E-02 | 2.449 |
| Day 0 vs Day 8 | Role of Hypercytokinemia/hyperchemokine in the Pathogenesis of Influenza | 1.70E+01 | 1.98E-01 | 4.123 |
| Day 0 vs Day 10 | Systemic Lupus Erythematosus In B Cell Signaling Pathway | 2.14E+01 | 5.67E-02 | 2.828 |
| Day 0 vs Day 10 | IL-15 Signaling | 2.04E+01 | 6.63E-02 | NaN |
| Day 0 vs Day 10 | B Cell Receptor Signaling | 1.79E+01 | 5.55E-02 | NaN |
| Day 0 vs Day 10 | Communication between Innate and Adaptive Immune Cells | 1.35E+01 | 3.88E-02 | NaN |
| Day 0 vs Day 10 | Interferon Signaling | 1.24E+01 | 2.78E-01 | 2.53 |
| Day 0 vs Day 10 | Role of Hypercytokinemia/hyperchemokine in the Pathogenesis of Influenza | 1.10E+01 | 1.40E-01 | 3.464 |
| Day 0 vs Day 10 | Primary Immunodeficiency Signaling | 7.59E+00 | 1.43E-01 | NaN |
| Day 0 vs Day 10 | Role of Pattern Recognition Receptors in Recognition of Bacteria and Viruses | 4.20E+00 | 5.13E-02 | 2 |
| Day 0 vs Day 10 | Activation of IRF by Cytosolic Pattern Recognition Receptors | 3.61E+00 | 7.69E-02 | 0.447 |
| Day 0 vs Day 10 | Pyrimidine Deoxyribonucleotides De Novo Biosynthesis I | 3.01E+00 | 1.30E-01 | NaN |
| Day 0 vs Day 14 | IL-15 Signaling | 1.08E+02 | 2.05E-01 | NaN |
| Day 0 vs Day 14 | B Cell Receptor Signaling | 9.91E+01 | 1.71E-01 | NaN |
| Day 0 vs Day 14 | Systemic Lupus Erythematosus In B Cell Signaling Pathway | 9.24E+01 | 1.49E-01 | NaN |
| Day 0 vs Day 14 | Communication between Innate and Adaptive Immune Cells | 8.17E+01 | 1.17E-01 | NaN |
| Day 0 vs Day 14 | Kinetochore Metaphase Signaling Pathway | 1.54E+01 | 1.62E-01 | 3.207 |
| Day 0 vs Day 14 | Primary Immunodeficiency Signaling | 1.06E+01 | 1.96E-01 | NaN |
| Day 0 vs Day 14 | Estrogen-mediated S-phase Entry | 7.95E+00 | 2.69E-01 | 2.646 |
| Day 0 vs Day 14 | Mitotic Roles of Polo-Like Kinase | 7.29E+00 | 1.36E-01 | 1.414 |
| Day 0 vs Day 14 | Cyclins and Cell Cycle Regulation | 6.37E+00 | 1.07E-01 | 3 |
| Day 0 vs Day 14 | Role of CHK Proteins in Cell Cycle Checkpoint Control | 5.48E+00 | 1.23E-01 | -2.236 |

To provide additional insight into the kinetics and relative magnitude of DENV-3 elicited inflammation captured in the study we utilize a previously described DENV transcriptional inflammation index to score each visit for each subject. Using this metric, the transcriptional signature of DENV-elicited inflammation peaked in all individuals between days 6 and 8 post challenge, returning to near baseline levels by day 14 post challenge mirroring the kinetics of viremia observed in this study (**Figure 4G**). Furthermore, the abundance of unique IGH clones assembled from the unenriched RNAseq data – a surrogate indicator of activated B cell frequency – exhibited a significant increase from baseline starting on day 10 post infection, peaking in all individuals on day 14, and returning to baseline by day 28 (**Figure 5H**).

### DENV-3 elicited plasmablast profiling

In light of the robust B cell activation profile observed by RNAseq following CH53489 infection we endeavored to better define the clonal and transcriptional diversity of activated B cells (plasmablasts) elicited by DENV-3 infection in the study. To this end, CD38+CD27+ plasmablast-phenotype CD19+ B cells were sorted from the PBMC of four subjects with the most robust B cell activation profile by as defined by RNAseq analysis (**Figure 6A, supplemental Figure 10**). After sorting, these purified plasmablasts were subjected to scRNAseq analysis using the 10xGenomics platform. This resulted in the capture of 15,717 DENV-elicited plasmablasts with full-length/paired immunoglobulin sequences.

**Figure 6.**
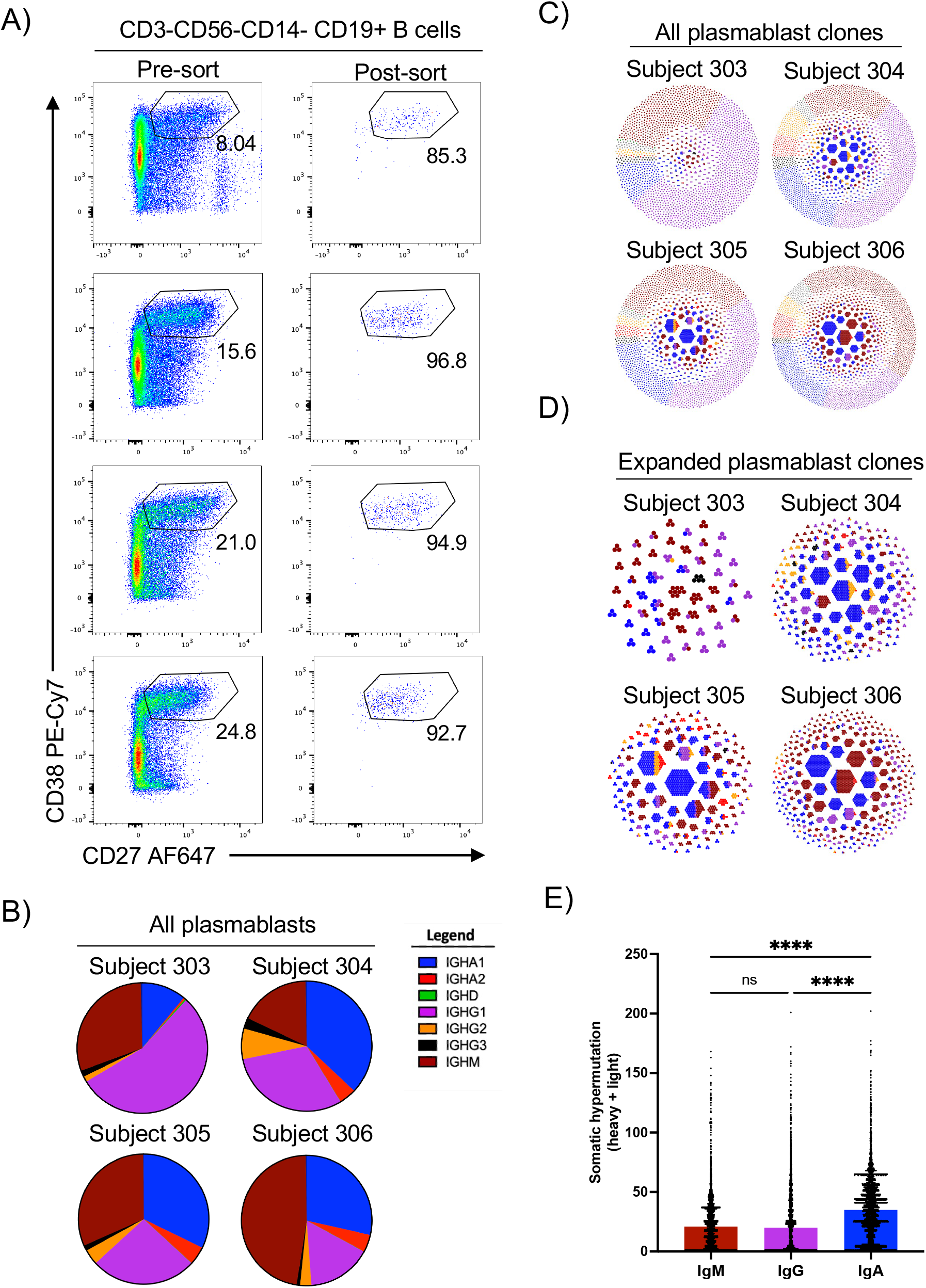
Isolation and clonal characterization of DENV-3 elicited plasmablasts. **A)** Identification and post-sort purity of DENV-3 elicited plasmablasts **B)** Isotype distribution of DENV-3 elicited plasmablasts **C)** Clonal diversity and isotype distribution of all DENV-3 elicited plasmablasts **D**) Clonal diversity and isotype distribution of expanded (n > 3) DENV-3 elicited plasmablasts **E)** Somatic hypermutation burden of DENV-3 elicited plasmablasts separated by isotype. **** p < 0.0001 one-way ANOVA with correction for multiple comparisons.

Consistent with our previously published report analyzing the plasmablast profile elicited by natural primary DENV infection [25], three of the four subjects included in this analysis exhibited a significant IgA1/IgM isotype bias with numerous large/expanded clonal families, while one subject exhibited a more IgG1/IgM isotype bias with significantly less clonal expansion (**Figure 6B, Figure 6C, Figure 6D**). Many clonal families were observed with members with IgM, IgA, and IgG Fc regions (**Figure 6C, Figure 6D**). Relatively low levels of somatic hypermutation (SHM) were observed in IgM and IgG isotype antibodies captured in this analysis, with IgA isotype antibodies on average exhibited significantly higher SHM burdens than their IgM and IgG counterparts.

### Virologic and immunologic correlates of clinical dengue

We next endeavored to clarify the correlative relationships between the individual virologic, clinical, and immunologic parameters captured in this analysis. We selected three virologic parameters (peak RNAemia, viremia, and NS1 antigenemia), four clinical parameters (days with rash, fatigue, headache, or fever), five clinical labs (AUC of ALT, AST, WBC, neutrophil and platelet counts outside of normal range), seven immunologic parameters (DENV-specific IgM/IgA/IgG AUC, NS1-specific IgM/IgA/IgG AUC, and day 90 ELISPOT counts), five serum cytokine features (IL-1Ra, IP-10, IFN-A2a, IFNg, and IL-10 AUC), and two transcriptional features (inflammation score and number of peak annotated IGH clones) for inclusion in this analysis (**Figure 7A**).

**Figure 7.**
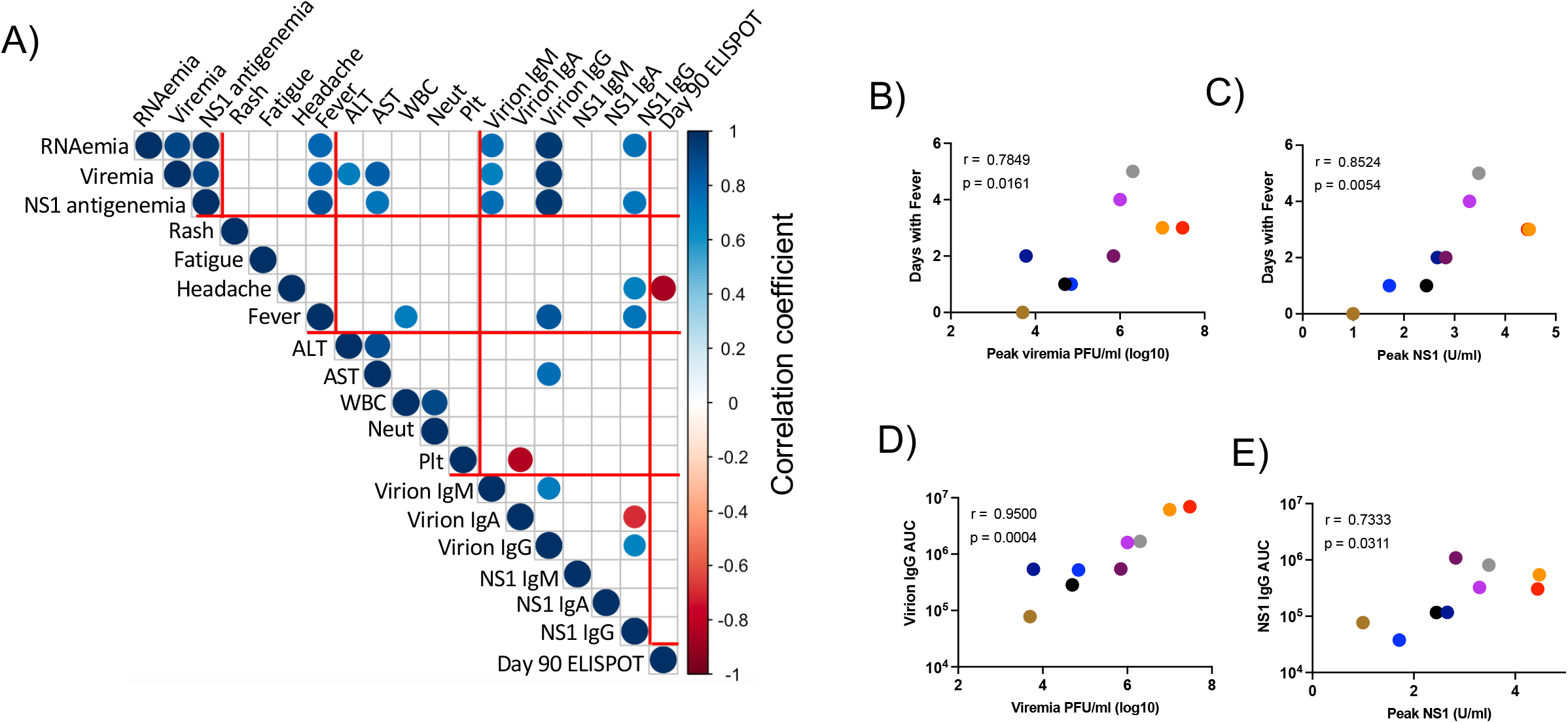
Correlation analysis of selection virologic, clinical, and immunologic features post CH53489 challenge. **A)** Correlation matrix. Spearman correlation. Only interactions with a p < 0.05 shown. **B**) Peak viremia vs total days with fever for each subject. **C)** Peak NS1 antigen levels vs total days with fever for each subject **D)** Peak viremia vs virion-specific IgG AUC **E**) Peak NS1 antigen levels vs NS1-specific IgG AUC

All virologic parameters exhibited a high degree of correlation with multiple clinical parameters such as number of days with fever and ALT/AST levels (**Figure 7A, Figure 7B, Figure 7C**). In addition, peak viral loads correlated with DENV-specific IgM and IgG titers, and anti-NS1 IgG titers (**Figure 7A, Figure 7D, Figure 7E**). These results support a model wherein the incidence and severity of mild DENV-elicited clinical pathology – as well as the magnitude of DENV-elicited adaptive immunity – is directly proportional to viral burden in previously flavivirus naïve individuals.

## DISCUSSION

In this study we precisely define the virologic, immunologic, and clinical features associated with a primary DENV-3 infection in flavivirus-naïve adults. This includes the kinetics and composition of the innate, humoral, cellular, and transcriptional responses elicited by experimental DENV-3 infection, as well as the virologic and clinical features of DENV-3 infection in 9 individuals inoculated with the under-attenuated DENV-3 strain CH53489. A notable feature of the results presented in our current study is the strong correlation observed between peak viral load and the frequency and magnitude of the clinical and immunological features elicited by DENV-3 infection. While the relationship between peak viral load, symptom severity, and subsequent anti-DENV immune profiles has been well established in individuals experiencing clinically apparent dengue [7], to our knowledge this is the first time this relationship has been extended to include individuals with mild or asymptomatic primary DENV infections. Importantly, these results support the utility of dengue human challenge models in evaluating the potential clinical benefit of candidate anti-dengue countermeasures where reduction in disease burden through the reduction in viral replication is an objective.

In addition to the spectrum of virologic and clinical features of primary DENV-3 infection captured in this analysis, several notable immunologic responses to CH53489 infection were observed. This includes a robust IgA1-biased plasmablast response after the resolution of acute viremia and infection-elicited inflammation. The presence of an IgA-dominated plasmablast response following primary DENV infection has now been described by multiple groups using several different analytic approaches [25-28]. However, the origin of this IgA biased response remains unclear. The extensive hypermutation burden present in these IgA class-switched plasmablasts might suggest that these cells arose from a pool of pre-existing IgA1 class-switched memory B cells that cross-react with DENV antigens. However, data presented in this study suggest this might not be the case, as B cell clones that have class-switched to IgM, IgG, and IgA are observed. These results are more consistent with a model wherein the IgA1 isotype plasmablasts arose from the same population of naïve B cells as IgM and IgG1 isotype cells. In addition, the transient nature of this DENV-elicited IgA response suggests that these cells are unable to take up residency in the bone marrow as long-lived plasma cells. While the biological significance of this observation is unclear, the short-lived nature of serum-derive DENV-specific IgA produced following both natural and experimental DENV infection has interesting implications from an epidemiological and serosurveillance perspective.

There are some limitations of this study to consider. Firstly, the relativity small number of subjects included in this analysis limits the generalizability of the observations. Secondly, the virus used in this study is delivered by intradermal injection rather than by an infected mosquito. The presence of mosquito saliva and/or salivary proteins (MSPs) has been shown to impact the kinetics and magnitude of DENV viremia in several experimental models of DENV infection [29], and it is currently unclear how the absence of this signal impacts the virologic and clinical progression of DENV infection in humans.

In conclusion, we have demonstrated the ability to safely infect flavivirus naïve volunteers with an under-attenuated DENV-3 strain CH53489. This infection results in a dengue-like illness with symptoms, clinical lab abnormalities, and immune responses consistent with an uncomplicated natural DENV infection. We demonstrated the magnitude and frequency of symptoms and immunologic features correlated with peak viremia titers. The acute/innate immune response to CH53489 is evident as early as 6 days post infection and persists through at least 10 days post infection, with the induction of an adaptive anti-DENV immune response evident 14 days post infection. These results provide hitherto unachievable insight into the kinetics and composition of DENV-3 elicited immune responses in flavivirus naïve individuals and provide an array of DENV-elicited biomarkers that can inform the development and characterization of anti-DENV countermeasures.

## MATERIALS AND METHODS

### Dengue human infection model

The Dengue Human Infection Model and associated analysis was approved by the State University of New York Upstate Medical University (SUNY-UMU) and the Department of Defense’s Human Research Protection Office. This phase 1, open-label study (ClinicalTrials.gov identifier: NCT04298138) was conducted between August 2020 and July 2021 at the State University of New York, Upstate Medical University (SUNY-UMU) in Syracuse, New York. Participants received a single subcutaneous inoculation of 0.7 × 10^3^ PFU (0.5ml of a 0.7 × 10^3^ PFU solution) of the CH53489 DENV-3 infection strain virus manufactured at the WRAIR Pilot Bioproduction Facility, Silver Spring, MD (US FDA Investigational New Drug 19321). All participants were pre-screened to ensure an absence of preexisting flavivirus using the Euroimmun dengue, West Nile, and Zika IgG ELISA kits (Lübeck, Germany). Subjects were monitored in an outpatient setting unless the hospitalization criteria were met. Criteria for subject hospitalization included fever of > 101 at 2 time points at least 4 hours apart, headache ≥ grade 2, eye pain ≥ grade 2, bone pain > grade 2, joint pain > grade 2, abdominal pain > grade 2, muscle pain > grade 2, nausea and/or vomiting > grade 2, liver function tests (ALT, AST) > grade 2, leukopenia > grade 2, thrombocytopenia ≥ grade 2, fatigue ≥ grade 2, or the presence any symptoms determined by the PI to warrant hospital admission. Study participants were discharged when the following criteria were met: no fever (< 100.4°F), laboratory parameters resolving and were ≤ grade 2 (at clinician discretion), and clinical symptoms were resolving and were ≤ grade 2 (at clinician discretion). Quantitative DENV-3 specific reverse-transcriptase polymerase chain reaction (RT-PCR) was performed using previously published techniques [30]. Serum NS1 antigen levels were quantified using a Euroimmun dengue NS1 ELISA kit (Lübeck, Germany).

### DENV-3 virion-capture ELISA

DENV-3 reactive serum IgM/IgA/IgG levels were assessed using a flavivirus capture ELISA protocol. In short, 96 well NUNC MaxiSorb flat-bottom plates were coated with 2 μg/ml flavivirus group-reactive mouse monoclonal antibody 4G2 (Envigo Bioproducts, Inc.) diluted in borate saline buffer. Plates were washed and blocked with 0.25% BSA +1% Normal Goat Serum in PBS after overnight incubation. DENV-3 (strain CH53489) was captured for 2 hours, followed by extensive washing. Serum samples were serially diluted four-fold, plated and incubated for 1 hour at room temperature on the captured virus. DENV-specific IgM/IgG/IgA levels were quantified using anti-human IgM HRP (Seracare, 5220-0328), anti-human IgG HRP (Southern BioTech, 2044-0), and anti-human IgA HRP (Biolegend, 411,002). Secondary antibody binding was quantified using the TMB Microwell Peroxidase Substrate System (KPL, cat. #50-76-00) and Synergy HT plate reader. DENV-3 (strain CH53489) was propagated in Vero cells and purified by ultracentrifugation through a 30% sucrose solution. End-point titers were determined as the reciprocal of the final dilution at which the optical density (OD) was greater than 2× of a pool of control flavivirus naïve serum at the same dilution. Day of seroconversion was defined as the day at which a participant’s end-point titer exceeded that of their respective day 0 sample

### Anti-NS1 opsonization assay

DENV-3 NS1 expressing CEM.NK^R^ cells (**fig. S3**) were stained with a 1:50 dilution of heat-inactivated serum diluted in PBS at room temperature for 30 min. Cells were extensively washed, then stained with goat anti-human IgA AF647 (2050-31, Southern Biotech), goat anti-human IgG AF467 (2040-31, Southern Biotech), or goat anti-human IgM AF647(2020-31, Southern Biotech). Flow cytometry analysis was performed on a BD LSRII instrument, and data analyzed using FlowJo v10.2 software (Treestar). Reported MFI values are background subtracted, with background defined as the signal observed staining DENV-3 NS1 expressing CEM.NK^R^ cells in the absence of serum. Day of seroconversion was defined as the day at which a participant’s background-subtracted NS1 specific MFI increased by at least 2× over their respective day 0 value.

### Serum cytokine analysis

The abundance of cytokines/chemokines in patient serum was assessed using the MSD human U-PLEX Viral Combo 1 kit and the MESO QuickPlex SQ 120 instrument (MESO Scale Diagnostics, Rockville, MD) according to the manufacture’s recommendation.

### IFN-γ ELISPOT

Cryopreserved PBMC were thawed, washed twice, and placed in RPMI 1640 medium (Corning, Tewksbury, MA, USA) supplemented with 10% heat-inactivated fetal calf serum (Corning, 35-010-CV), L-glutamine (Lonza, Basel, Switzerland), and Penicillin/Streptomycin (Gibco, Waltham, MA, USA). Cellular viability was assessed by trypan blue exclusion and cells were resuspended at a concentration of 5 × 10^6^/mL and rested overnight at 37 °C. After resting, viable PBMC were washed, counted, and resuspended at a concentration of 1 × 10^6^/mL in complete cell culture media. Next, 100 μL of this cell suspension was mixed with 100 μL of the individual peptide pools listed in **Supplemental Table 11** and diluted to a final concentration 1 μg/mL/peptide (DMSO concentration 0.5%) in complete cell culture media. This cell and peptide mixture was loaded onto a 96-well PVDF plate coated with anti-IFN-γ (3420-2HW-Plus, Mabtech, Nacka, Sweden) and cultured overnight. Controls for each participant included 0.5% DMSO alone (negative) and anti-CD3 (positive). After overnight incubation, the ELISPOT plates were washed and stained with anti-IFN-γ-biotin followed by streptavidin-conjugated HRP (3420-2HW-Plus, Mabtech). Plates were developed using TMB substrate and read using a CTL-ImmunoSpot^®^ S6 Analyzer (Cellular Technology Limited, Shaker Heights, OH, USA). All peptide pools were tested in duplicate, and the adjusted mean was reported as the mean of the duplicate experimental wells after subtracting the mean value of the negative (DMSO only) control wells. Individuals were considered reactive to a peptide pool when the background-subtracted response was >50 spot forming cells (SFC)/10^6^ PBMC. All data were normalized based on the number of cells plated per well and are presented herein as SFC/10^6^ PBMC.

### RNA sequencing library preparation and sequencing

Whole blood was collected from all study participants using PAXgene RNA collection tubes (BD) and frozen at -20° C until analyzed. RNA was recovered from the collection tubes using the Qiagen PAXgene Blood RNA isolation Kit and sequencing libraries created using Illumina Stranded Total RNA Prep with Ribo-Zero Plus and IDT-Ilmn RNA UD Indexes SetA. Final library QC and quantification was performed using a Bioanalyizer (Agilent) and DNA 1000 reagents. Libraries were pooled at an equimolar ratio and sequenced on a 300 cycle Novaseq 6000 instrument using v1.5 S4 reagent set.

### RNA sequencing gene expression analysis

Raw reads from demultiplexed FASTQ files were mapped to the human reference transcriptome (Ensembl, Home sapiens, GRCh38) using Kallisto [31] version 0.46.2. Transcript-level counts and abundance data were imported and summarized in R (version 4.0.2) using the TxImport package [32] and TMM normalized using the package EdgeR [33, 34]. Differential gene expression analysis performed using linear modeling and Bayesian statistics in the R package Limma [35]. Genes with a log_2_ fold change of >2 and a Benjamini-Hochberg adjusted p-value < 0.01 were considered significant. Gene module scores were calculated by summing the TMM normalized abundance (TPM) of the genes highlighted as belonging to either Module 1 or Module 2 for all samples.

### BCR clonotype identification and annotation from bulk RNAseq

Raw FASTQ files were filtered to contain only pair-end reads and to remove any Illumina adaptor contamination and low-quality reads using Trimmomatic (v0.39) [36]. Pair-end reads were subsequently analyzed using MiXCR (v3.0.3) using the RNA-seq/non-targeted genomic analysis pipeline [37, 38].

### Flow cytometry and cell sorting

Cryopreserved PBMC were thawed and placed in RPMI 1640 medium (Corning, Tewksbury, MA, USA) supplemented with 10% heat-inactivated fetal calf serum (Corning, 35-010-CV), L-glutamine (Lonza, Basel, Switzerland), and Penicillin/Streptomycin (Gibco, Waltham, MA, USA). Cellular viability was assessed by trypan blue exclusion. Surface staining for flow cytometry analysis and cell sorting was performed in PBS supplemented with 2% FBS at room temperature. Aqua Live/Dead (ThermoFisher, L34957) was used to exclude dead cells in all experiments. Antibodies and dilutions used for flow cytometry analysis are listed in **Supplemental Table 12**. Cell sorting was performed on a BD FACSAria III instrument, with cells sorted directly into 50 ul of complete cell culture media supplemented with 20% FCS. Data analyzed using FlowJo v10.2 software (Treestar).

### Single-cell RNA sequencing library generation

Flow-sorted viable CD3-CD56-CD14-CD19+CD27+CD38+ plasmablast phenotype B cells were prepared for single-cell RNA sequencing using the Chromium NextGEM Single-Cell 5′ Reagent version 2 kit and Chromium Single-Cell Controller (10x Genomics, CA) [39]. Approximately 2000–8000 cells per reaction suspended at a density of 50–500 cells/μL in PBS plus 0.5% FBS were loaded for gel bead-in-emulsion (GEM) generation and barcoding. Reverse transcription, RT-cleanup, and cDNA amplification were performed to isolate and amplify cDNA for enriched V(D)J library construction according to the manufacturer’s protocol. Libraries were constructed using the Chromium Single-Cell 5′ reagent kit, V(D)J Human B Cell Enrichment Kit, and Dual Index Kit (10x Genomics, CA) according to the manufacturer’s protocol.

### Single-cell RNA sequencing library Sequencing

scRNAseq BCR V(D)J enriched libraries were sequenced on an Illumina NextSeq 2000 instrument using the 300 cycle P1 reagent kit. Libraries were balanced to allow for ∼10,000 reads/cell for BCR V(D)J enriched libraries. Sequencing parameters were set for 150 cycles for Read1, 10 cycles for Index1, 10 cycles for Index2, and 150 cycles for Read2. Prior to sequencing, library quality and concentration were assessed using an Agilent 4200 TapeStation with High Sensitivity D5000 ScreenTape Assay according to the manufacturer’s recommendations.

### Immunoglobulin sequence analysis

Sorted B cell immunoglobulin clonotype identification, alignment, and annotation was performed using the 10x Genomics Cell Ranger pipeline [39] Sample demultiplexing and clonotype alignment was performed using the Cell Ranger software package (10x Genomics, CA, v5.0.0) and bcl2fastq2 (Illumina, CA, v2.20) according to the manufacturer’s recommendations, using the default settings and mkfastq/vdj commands, respectively. Immunoglobulin clonotype alignment was performed against a filtered human V(D)J reference library. Paired immunoglobulin hypermutation burden was assessed using the software package BRILIA [40], and clonal lineages defined and visualized using the enclone software suite from 10xGenomics.

### Statistical analysis

All statistical analyses other than RNAseq gene expression analysis was performed using GraphPad Prism 9 Software (GraphPad Software, La Jolla, CA). A P-value□<□0.05 was considered significant.

## Data Availability

The authors declare that all data supporting the findings of this study are available within this article and its Supplementary Information files, or from the corresponding author upon reasonable request. RNAseq gene expression data have been deposited in the NCBI Gene Expression Omnibus (GEO GSE216328).

## ACKNOWLEDGEMENTS

We gratefully acknowledge excellent technical assistance provided by Karen Gentile of the Upstate Medical University Molecular Analysis Core (MAC), Lisa Phelps of the SUNY Upstate Medical University Flow Cytometry Core, Holly Chanatry, and the members of the Institute for Global Health and Translational Science (IGHTS) of SUNY Upstate Medical University. We also acknowledge Calli Rooney from the US Army. We also wish to thank all the study participants for making this study possible. The following reagents were obtained through BEI Resources, NIAID, NIH: Peptide Array, DENV-3 Sleman/1978, E protein (NR-511), DENV-3 Philippines/H87/1956, E protein (NR-9228), DENV-1 Philippines/H87/1956, NS1 protein (NR-2753), DENV-3 Philippines/H87/1956, NS3 protein (NR-2754), DENV-3 Philippines/H87/1956, NS5 protein (NR-4204). The opinions or assertions contained herein are the private views of the authors and are not to be construed as reflecting the official views of the US Army or the US Department of Defense. Material has been reviewed by the Walter Reed Army Institute of Research. There is no objection to its presentation and/or publication. The investigators have adhered to the policies for protection of human participants as prescribed in AR 70-25.

## Author contributions

Conceptualization: L.W., M.D.K, R.G.J., T.P.E., and S.J.T. Formal analysis: L.W., A.T.W. Funding acquisition: H.F, R.G.J., T.P.E., and S.J.T. Investigation: A.T.W., J.Q.L., K.N., H.S.F., M.W., C.G., M.D.K., L.W., T.P.E., S.J.T. Resources: H.F., R.G.J., J.R.C. Visualization: K.N., A.T.W. Writing – original draft: A.T.W., S.J.T. Writing – review & editing: all authors

## References

1. Bhatt, S., et al., The global distribution and burden of dengue. Nature, 2013. 496(7446): p. 504–7.

2. Shepard, D.S., et al., Economic impact of dengue illness in the Americas. Am J Trop Med Hyg, 2011. 84(2): p. 200–7.

3. Guzman, M.G., et al., Dengue infection. Nat Rev Dis Primers, 2016. 2: p. 16055.

4. Gubler, D.J., Aedes aegypti and Aedes aegypti-borne disease control in the 1990s: top down or bottom up. Charles Franklin Craig Lecture. Am J Trop Med Hyg, 1989. 40(6): p. 571–8.

5. Iwamura, T., A. Guzman-Holst, and K.A. Murray, Accelerating invasion potential of disease vector Aedes aegypti under climate change. Nat Commun, 2020. 11(1): p. 2130.

6. World mosquito program. Facts sheet Dengue. [cited 2021 27 April]; Available from: https://www.worldmosquitoprogram.org/sites/default/files/2020-11/WMP%20dengue_0.pdf.

7. Vaughn, D.W., et al., Dengue viremia titer, antibody response pattern, and virus serotype correlate with disease severity. J Infect Dis, 2000. 181(1): p. 2–9.

8. Anderson, K.B., et al., A shorter time interval between first and second dengue infections is associated with protection from clinical illness in a school-based cohort in Thailand. J Infect Dis, 2014. 209(3): p. 360–8.

9. Montoya, M., et al., Symptomatic versus inapparent outcome in repeat dengue virus infections is influenced by the time interval between infections and study year. PLoS Negl Trop Dis, 2013. 7(8): p. e2357.

10. Guzman, M.G., M. Alvarez, and S.B. Halstead, Secondary infection as a risk factor for dengue hemorrhagic fever/dengue shock syndrome: an historical perspective and role of antibody-dependent enhancement of infection. Arch Virol, 2013. 158(7): p. 1445–59.

11. Srikiatkhachorn, A., et al., Natural history of plasma leakage in dengue hemorrhagic fever: a serial ultrasonographic study. Pediatr Infect Dis J, 2007. 26(4): p. 283-90; discussion 291-2.

12. Rajapakse, S., et al., Prophylactic and therapeutic interventions for bleeding in dengue: a systematic review. Trans R Soc Trop Med Hyg, 2017. 111(10): p. 433–439.

13. Wong, J.G., et al., Identifying Adult Dengue Patients at Low Risk for Clinically Significant Bleeding. PLoS One, 2016. 11(2): p. e0148579.

14. Capeding, M.R., et al., Clinical efficacy and safety of a novel tetravalent dengue vaccine in healthy children in Asia: a phase 3, randomised, observer-masked, placebo-controlled trial. Lancet, 2014. 384(9951): p. 1358–65.

15. Villar, L., et al., Efficacy of a tetravalent dengue vaccine in children in Latin America. N Engl J Med, 2015. 372(2): p. 113–23.

16. World Health, O., Dengue vaccine: WHO position paper, July 2016 - recommendations. Vaccine, 2017. 35(9): p. 1200–1201.

17. Biswal, S., et al., Efficacy of a Tetravalent Dengue Vaccine in Healthy Children and Adolescents. N Engl J Med, 2019. 381(21): p. 2009–2019.

18. Biswal, S., et al., Efficacy of a tetravalent dengue vaccine in healthy children aged 4-16 years: a randomised, placebo-controlled, phase 3 trial. Lancet, 2020. 395(10234): p. 1423–1433.

19. Takeda’s QDENGA (Dengue Tetravalent Vaccine [Live, Attenuated]) Approved in Indonesia for Use Regardless of Prior Dengue Exposure. August 22, 2022:[Available from:https://www.takeda.com/newsroom/newsreleases/2022/takedas-qdenga-dengue-tetravalent-vaccine-live-attenuated-approved-in-indonesia-for-use-regardless-of-prior-dengue-exposure/.

20. Thomas, S.J. and T.P. Endy, Current issues in dengue vaccination. Curr Opin Infect Dis, 2013. 26(5): p. 429–34.

21. Low, J.G., E.E. Ooi, and S.G. Vasudevan, Current Status of Dengue Therapeutics Research and Development. J Infect Dis, 2017. 215(Suppl_2): p. S96–S102.

22. Endy, T.P., et al., A Phase 1, Open-Label Assessment of a Dengue Virus-1 Live Virus Human Challenge Strain. J Infect Dis, 2021. 223(2): p. 258–267.

23. Mammen, M.P., et al., Evaluation of dengue virus strains for human challenge studies. Vaccine, 2014. 32(13): p. 1488–94.

24. Sun, W., et al., Experimental dengue virus challenge of human subjects previously vaccinated with live attenuated tetravalent dengue vaccines. J Infect Dis, 2013. 207(5): p. 700–8.

25. Waickman, A.T., et al., Transcriptional and clonal characterization of B cell plasmablast diversity following primary and secondary natural DENV infection. EBioMedicine, 2020. 54: p. 102733.

26. Waickman, A.T., et al., Temporally integrated single cell RNA sequencing analysis of PBMC from experimental and natural primary human DENV-1 infections. PLoS Pathog, 2021. 17(1): p. e1009240.

27. Rouers, A., et al., CD27(hi)CD38(hi) plasmablasts are activated B cells of mixed origin with distinct function. iScience, 2021. 24(5): p. 102482.

28. Robinson, M.L., et al., Magnitude and kinetics of the human immune cell response associated with severe dengue progression by single-cell proteomics. bioRxiv, 2022: p. 2022.09.21.508901.

29. McCracken, M.K., et al., Route of inoculation and mosquito vector exposure modulate dengue virus replication kinetics and immune responses in rhesus macaques. PLoS Negl Trop Dis, 2020. 14(4): p. e0008191.

30. Houng, H.S., et al., Development of a fluorogenic RT-PCR system for quantitative identification of dengue virus serotypes 1-4 using conserved and serotype-specific 3’ noncoding sequences. J Virol Methods, 2001. 95(1-2): p. 19–32.

31. Bray, N.L., et al., Erratum: Near-optimal probabilistic RNA-seq quantification. Nat Biotechnol, 2016. 34(8): p. 888.

32. Soneson, C., M.I. Love, and M.D. Robinson, Differential analyses for RNA-seq: transcript-level estimates improve gene-level inferences. F1000Res, 2015. 4: p. 1521.

33. Robinson, M.D., D.J. McCarthy, and G.K. Smyth, edgeR: a Bioconductor package for differential expression analysis of digital gene expression data. Bioinformatics, 2010. 26(1): p. 139–40.

34. McCarthy, D.J., Y. Chen, and G.K. Smyth, Differential expression analysis of multifactor RNA-Seq experiments with respect to biological variation. Nucleic Acids Res, 2012. 40(10): p. 4288–97.

35. Ritchie, M.E., et al., limma powers differential expression analyses for RNA-sequencing and microarray studies. Nucleic Acids Res, 2015. 43(7): p. e47.

36. Bolger, A.M., M. Lohse, and B. Usadel, Trimmomatic: a flexible trimmer for Illumina sequence data. Bioinformatics, 2014. 30(15): p. 2114–20.

37. Bolotin, D.A., et al., MiXCR: software for comprehensive adaptive immunity profiling. Nat Methods, 2015. 12(5): p. 380–1.

38. Bolotin, D.A., et al., Antigen receptor repertoire profiling from RNA-seq data. Nat Biotechnol, 2017. 35(10): p. 908–911.

39. Zheng, G.X., et al., Massively parallel digital transcriptional profiling of single cells. Nat Commun, 2017. 8: p. 14049.

40. Lee, D.W., et al., BRILIA: Integrated Tool for High-Throughput Annotation and Lineage Tree Assembly of B-Cell Repertoires. Front Immunol, 2016. 7: p. 681.

