## Supplemental Figures for "Virologic, clinical, and immunological characteristics of a dengue virus 3 human challenge model"

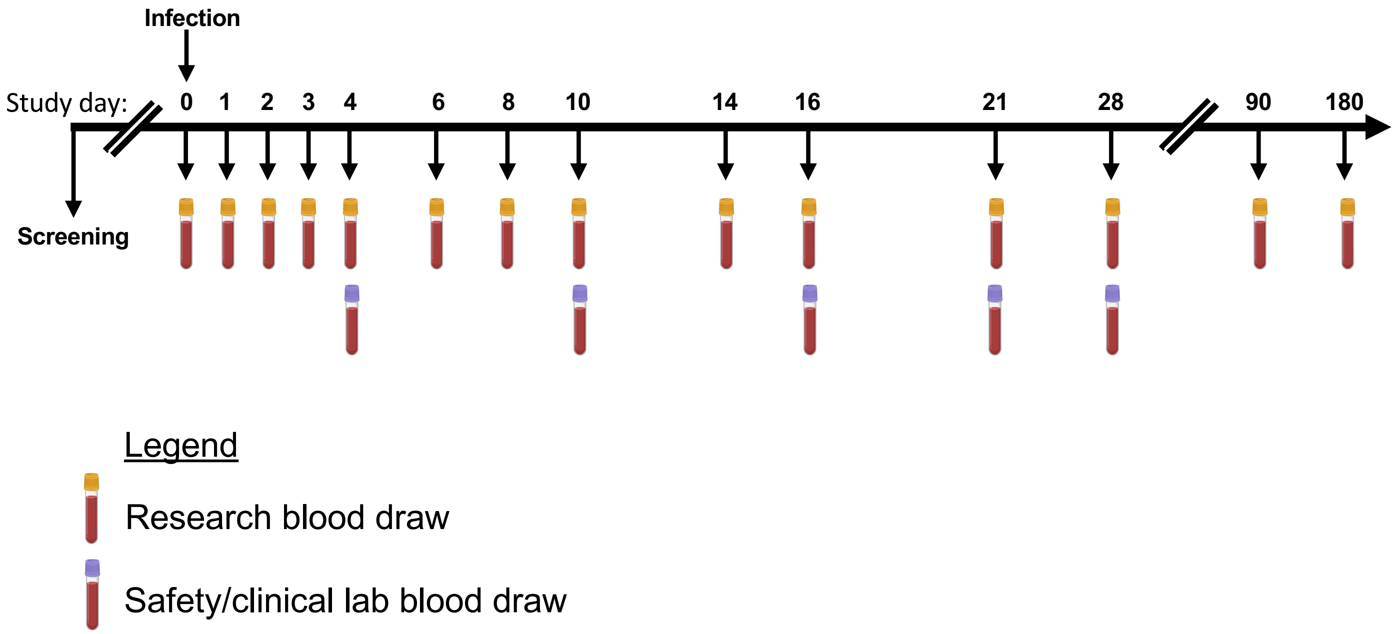


**Supplemental Figure 1.** Study schematic

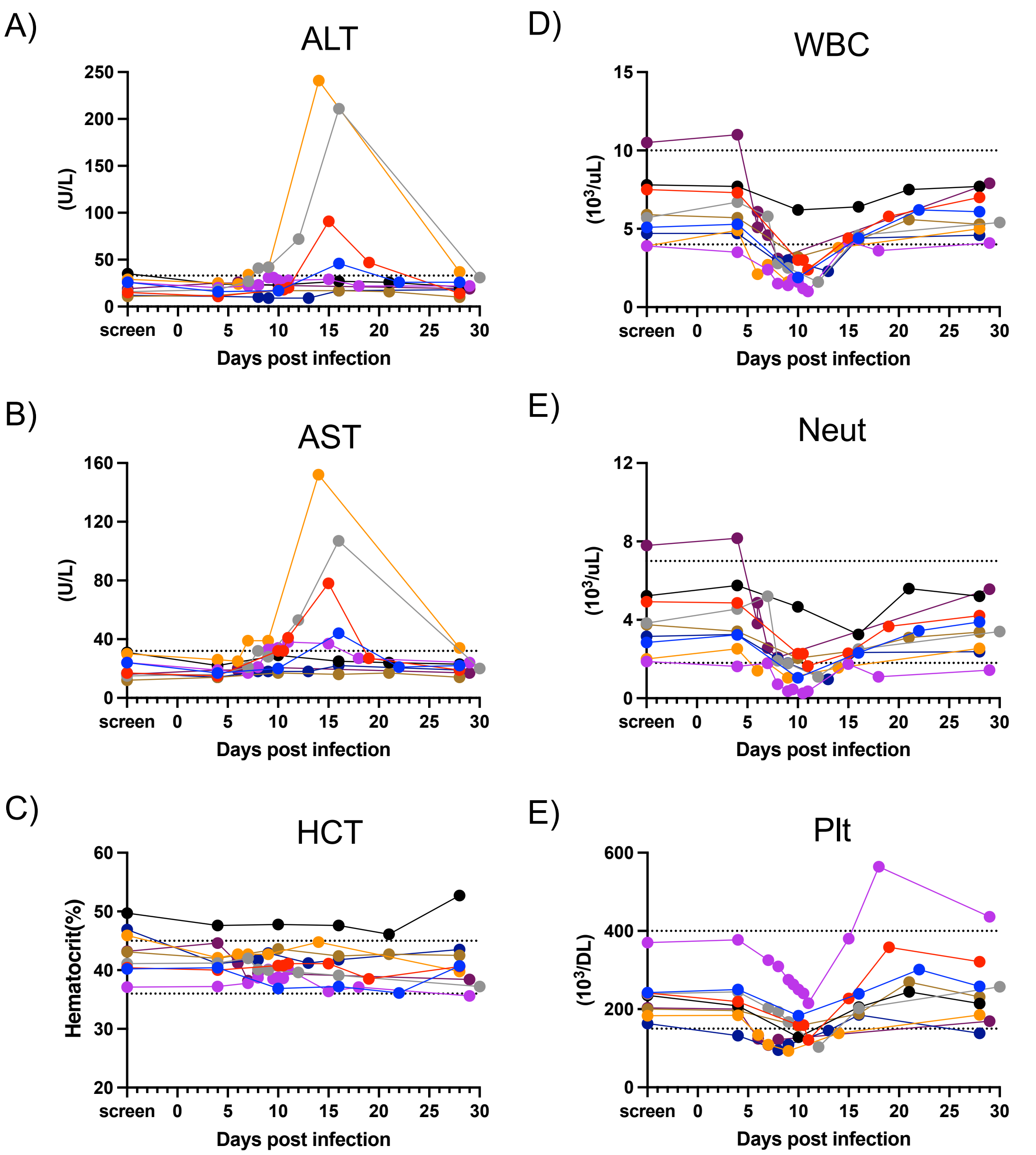


**Supplemental Figure 2.** Timing and severity of clinically significant adverse events in response to CH53489 challenge. Raw lab values. Normal high and normal low for each lab shown in dotted lines

A)

```
MNSKNTSMSFSCIAIGIITLYLGAVVQADMGCVINWKGKELKCGSGIFVTNEVHTWTEQYKFQADSPKRLATAIAGAWENG
VCGIRSTTRMENLLWKQIANELNYILWENNIKLTVVVGDIIGVLEQGKRTLTPQPMELKYSWKTWGKAKIVTAETQNSSFI
IDGPHTPECPRASRAWNVWEVEDYGFGVFTTNIWLKLREEYTQLCDHRLMSAAVKDERAVHADMGYWIESQKNGSWKLEKA
SLIEVKTCTWPKSHTLWSNGVLESDMIIPKSLAGPISQHNHRPGYHTQTAGPWHLGKLELDFNYCEGTTVVITENCGTRGP
SLRTTTVSGKLIHEWCCRSCTLPPLRYMGEDGCWYGMEIRPINEKEENMVKSLASAGSGKVDNFTMGFLCLAILFEEVMRG
KFGKKHMIAGVFLTFLLLLSGQITWRDMAHTLIMIGSNASDRMGMGVTYLALIATFKIQPFLALGFFLRKLTSRENFLLGV
GLAMATTLQLPEDIEQMANGIALGLMALKLITQFETYQLWTALVSLTCSNTIFTLTVAWRTATLILAGVSLLPVCQSSSMR
KTDWLPMAVAAMGVPPLPLFIFSLKDTLKRR
```

B)

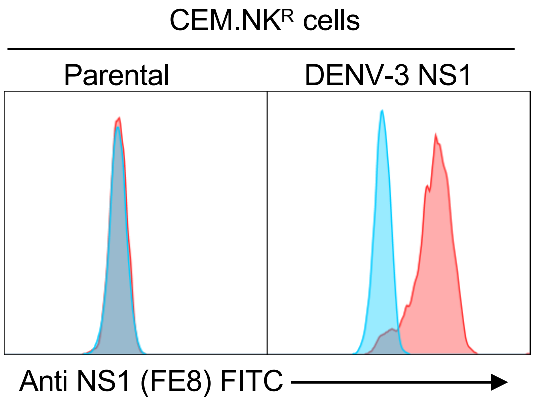


C)

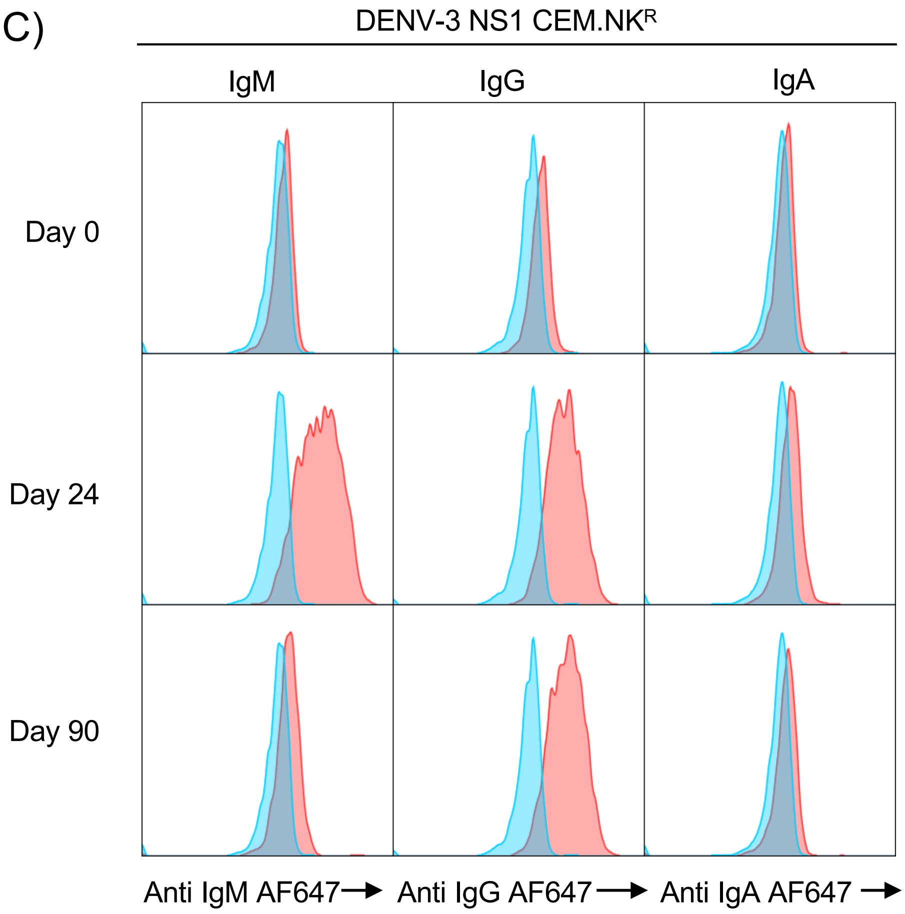


**Supplemental Figure 3. NS1 opsonization assay. A)** Sequence and annotation of the DENV-3 NS1 expression construct used in this study. Yellow = signal peptide, green = NS1, blue = NS2A. **B)** Anti-NS1 staining mAb staining (clone FE8) of parental CEM.NKR cell line and DENV-3 NS1 expressing CEM.NKR cells. Unstained cells shown in blue, FE8 stained shown in red **C)** Representative NS1 opsonizing activity of serum collected at days 0, 24, and 90 days post DENV-3 challenge. Unstained cells shown in blue, serum stained shown in red

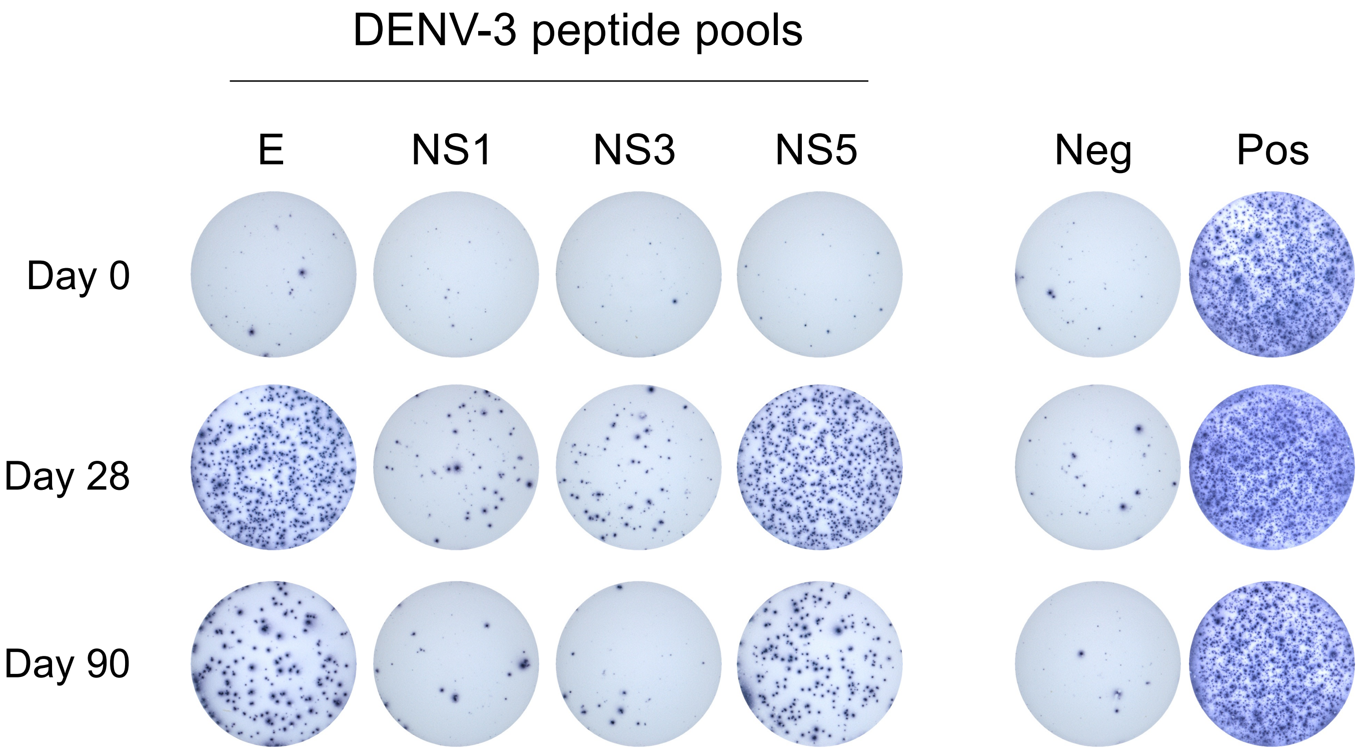


**Supplemental Figure 4.** Representative IFN-γ ELISPOT images

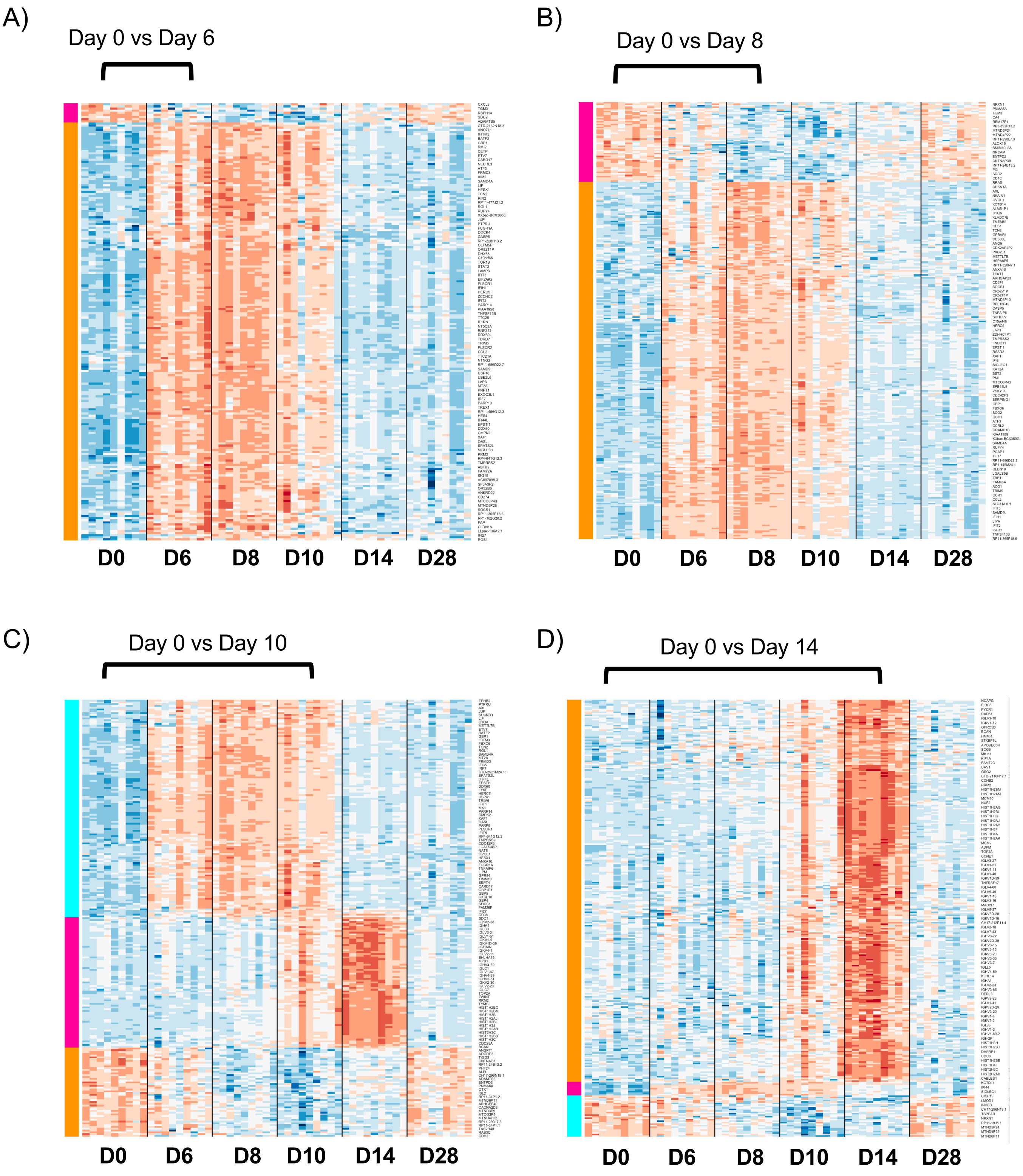


**Supplemental Figure 5.** Identification of gene modules with coordinated expression changes following DENV-3 challenge

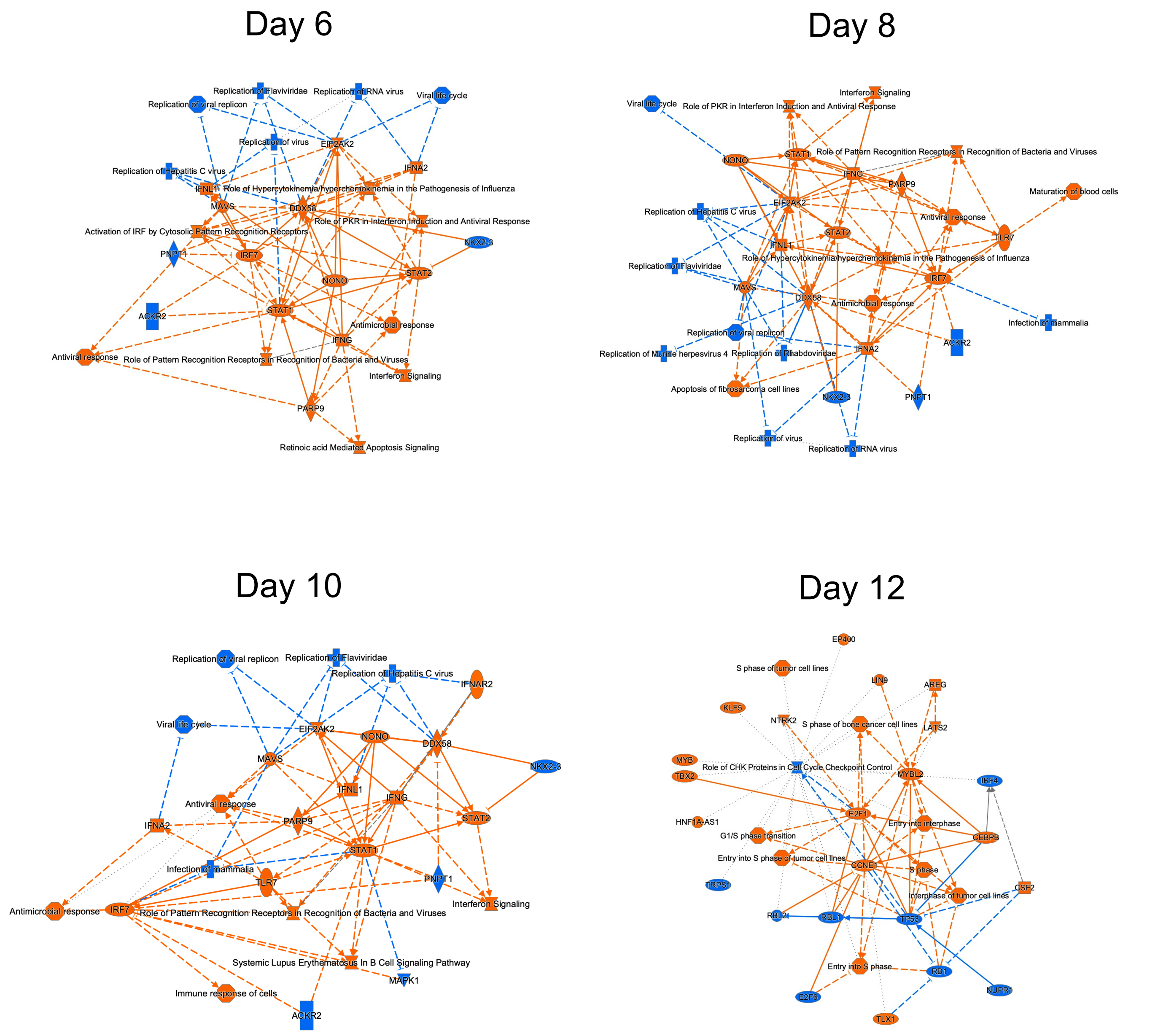


**Supplemental Figure 6.** Graphical network summary of IPA terms identified across differentially expressed genes on days 6, 8, 10, and 14 post DENV-3 infection

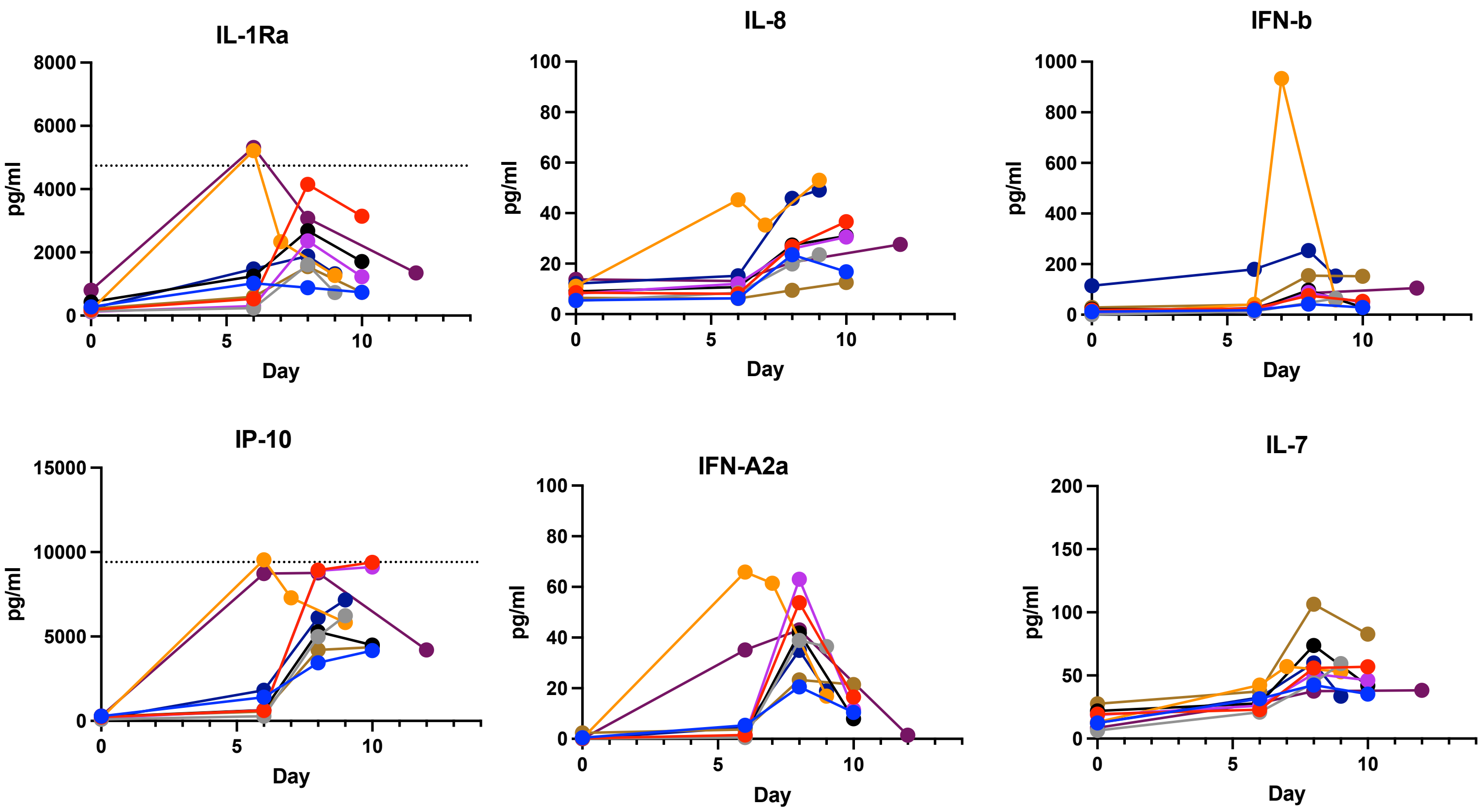


**Supplemental Figure 7. Multiplexed cytokine analysis.** Cytokines which exhibited induction days 6-8 post challenge. Dashed line indicates assay upper limit of quantification (ULOQ) for analytes with samples near or at the ULOQ.

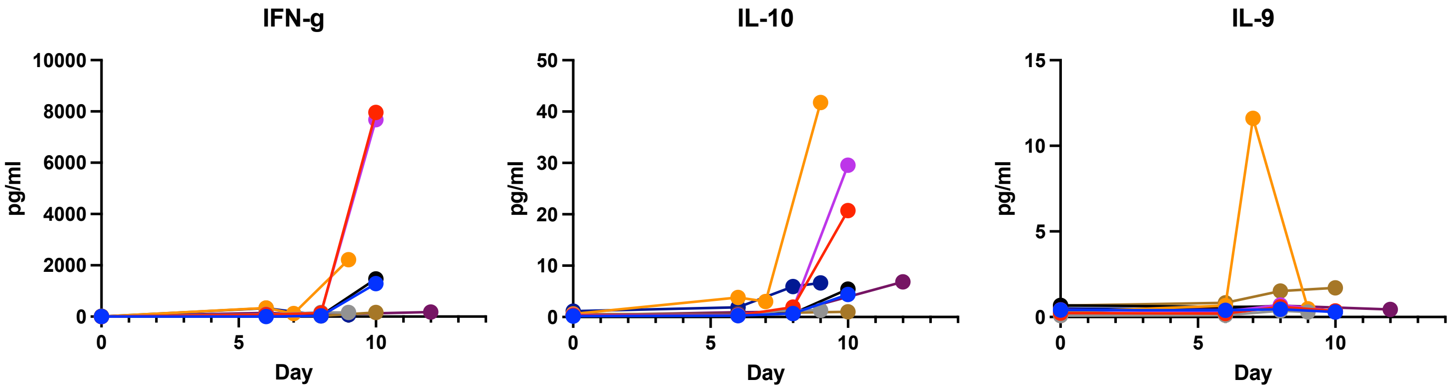


**Supplemental Figure 8. Multiplexed cytokine analysis.** Cytokines which exhibited induction days 8+ post challenge

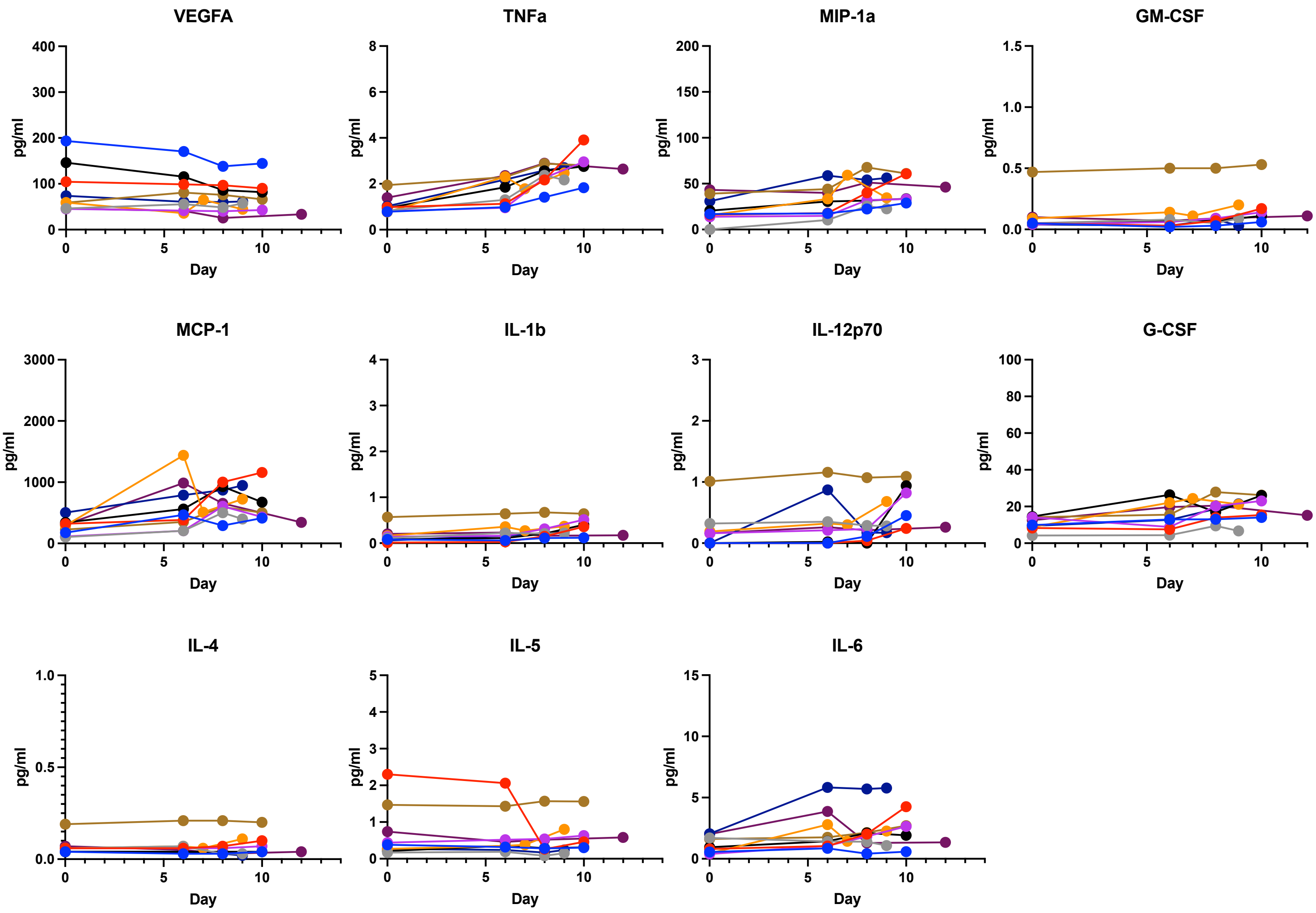


**Supplemental Figure 9. Multiplexed cytokine analysis.** Cytokines which exhibited no change from baseline following challenge

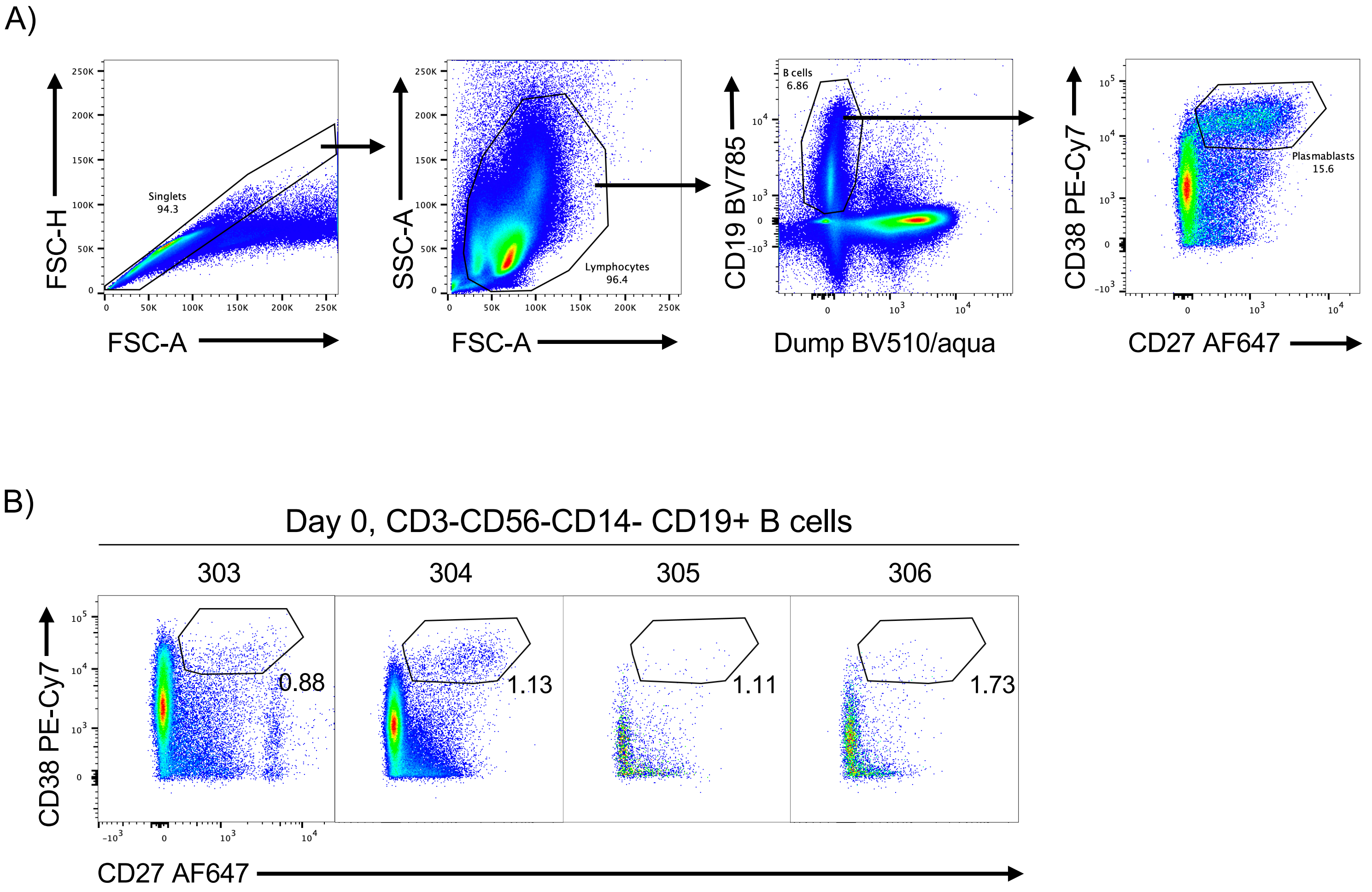


**Supplemental Figure 10. A)** Gating scheme for flow cytometric isolation of DENV-3 elicited plasmablasts**. B)** Plasmablast frequency in all subjects on day 0.
