## Supplemental Figures for "Virologic, clinical, and immunological characteristics of a dengue virus 3 human challenge model"

**Supplemental Table 1.** Study demographics

|  | **All Participants (N=9)** |
| --- | --- |
| Age (years) |  |
| Mean (SD) | 32.7 (5.5) |
| Median | 34 |
| Min, Max | 22, 40 |
| Sex, n (%) |  |
| Male | 4 (44.4) |
| Female | 5 (55.6) |
| Ethnicity, n (%) |  |
| Hispanic or Latino | 0 (0) |
| Non-Hispanic or Latino | 9 (100) |
| Race, n (%) |  |
| White | 8 (88.9) |
| Black or African American | 1 (11.1) |
| American Indian or Alaska Native | 0 |
| Asian | 0 |
| Native Hawaiian or Other Pacific  Islander | 0 |
| Other or Multiple | 0 |

**Supplemental Table 2.** Sample

| **Subject** | **Days analyzed** | **Peak RNAemia** | **Clinical outcome** |
| --- | --- | --- | --- |
| **301** | **0, 6, 8, 10, 14, 28** | **1.37E+05 (D6)** | **Non-hospitalized** |
| **302** | **0, 6, 8, 10, 15, 28** | **7.02E+08 (D8)** | **Hospitalized (D11)** |
| **303** | **0, 6, 8, 9, 16, 28** | **6.89E+07 (D8)** | **Hospitalized (D8)** |
| **304** | **0, 6, 8, 10, 15, 28** | **3.65E+07 (D7)** | **Hospitalized (D7)** |
| **305** | **0, 6, 7, 9, 14, 28** | **3.51E+08 (D6)** | **Hospitalized (D6)** |
| **306** | **0, 6, 8, 10, 14, 28** | **2.94E+05 (D6)** | **Non-hospitalized** |
| **307** | **0, 6, 8, 10, 14, 28** | **3.13E+04 (D8)** | **Non-hospitalized** |
| **308** | **0, 6, 8, 9, 13, 28** | **7.41E+05 (D6)** | **Hospitalized (D8)** |
| **309** | **0, 6, 8, 12, 14, 28** | **4.57E+07 (D6)** | **Hospitalized (D7)** |

**Supplemental Table 4.** Differentially expressed genes day 6 post infection

| **geneID** | **logFC** | **AveExpr** | **t** | **P.Value** | **adj.P.Val** | **B** |
| --- | --- | --- | --- | --- | --- | --- |
| CCL8 | 7.24694862 | -0.582129 | 4.52812693 | 3.49E-05 | 8.86E-04 | 1.64313092 |
| CCL2 | 6.78786833 | 1.13589506 | 4.15947308 | 1.20E-04 | 0.00241416 | 1.01327106 |
| CXCL10 | 6.39526565 | 3.2144798 | 5.33360406 | 2.11E-06 | 8.15E-05 | 4.79034107 |
| USP18 | 5.59715875 | 5.2942684 | 11.6691949 | 3.76E-16 | 4.70E-13 | 26.344708 |
| RSAD2 | 5.44826657 | 8.78506662 | 9.2344631 | 1.48E-12 | 3.17E-10 | 18.4160915 |
| SIGLEC1 | 5.40349977 | 6.35550789 | 8.68415284 | 1.06E-11 | 1.55E-09 | 16.5018671 |
| IFI44L | 5.22576116 | 9.26563843 | 9.83373779 | 1.81E-13 | 5.40E-11 | 20.4632313 |
| USP41 | 5.0671684 | 1.09222199 | 7.18418303 | 2.49E-09 | 1.99E-07 | 10.4101663 |
| ISG15 | 5.03519693 | 0.3381444 | 6.18001737 | 9.93E-08 | 5.39E-06 | 7.10535338 |
| SLC31A1P1 | 5.00323265 | -2.106749 | 6.6240472 | 1.95E-08 | 1.27E-06 | 6.9275423 |
| IFIT1 | 4.91409063 | 8.99783691 | 8.86479883 | 5.53E-12 | 9.28E-10 | 17.1331345 |
| EXOC3L1 | 4.78399636 | 2.25582378 | 8.14120566 | 7.52E-11 | 8.88E-09 | 14.3287761 |
| HESX1 | 4.71632926 | -1.1411462 | 4.14770713 | 1.24E-04 | 0.00249572 | 0.6192598 |
| OAS3 | 4.70287739 | 9.74421226 | 9.52306857 | 5.37E-13 | 1.24E-10 | 19.3933447 |
| CMPK2 | 4.64542572 | 7.47598457 | 9.89258921 | 1.48E-13 | 4.70E-11 | 20.6748976 |
| HES4 | 4.62080875 | 2.18929656 | 6.39214748 | 4.57E-08 | 2.67E-06 | 8.36312393 |
| BATF2 | 4.58294216 | 5.07853875 | 8.65852 | 1.16E-11 | 1.66E-09 | 16.4120321 |
| IFI44 | 4.56125273 | 7.7459889 | 9.9254505 | 1.32E-13 | 4.34E-11 | 20.7865323 |
| LAMP3 | 4.53410893 | 4.21810844 | 11.3339977 | 1.13E-15 | 1.06E-12 | 25.3482868 |
| IFI6 | 4.51175648 | 8.08614509 | 10.5934319 | 1.34E-14 | 7.25E-12 | 23.018309 |
| IFIT3 | 4.48796715 | 9.34813991 | 8.87669214 | 5.30E-12 | 8.97E-10 | 17.173614 |
| HERC5 | 4.45623544 | 7.68061448 | 9.92083206 | 1.34E-13 | 4.34E-11 | 20.7700682 |
| ETV7 | 4.41031227 | 4.46985866 | 7.45330116 | 9.28E-10 | 8.38E-08 | 12.1531407 |
| SERPING1 | 4.39254423 | 5.97008766 | 8.17127912 | 6.74E-11 | 8.12E-09 | 14.6888939 |
| OASL | 4.30691463 | 7.54938765 | 10.8848898 | 5.00E-15 | 3.03E-12 | 23.9831096 |
| DZIP1L | 4.28999938 | 1.33912248 | 5.04747402 | 5.81E-06 | 1.89E-04 | 3.85042703 |
| TMPRSS2 | 4.23015467 | -1.0412947 | 4.83591034 | 1.22E-05 | 3.58E-04 | 2.34318884 |
| ATF3 | 4.19804021 | 2.72288822 | 5.89911689 | 2.76E-07 | 1.33E-05 | 6.70582538 |
| IFIT2 | 4.18745971 | 9.66057711 | 8.98637775 | 3.58E-12 | 6.60E-10 | 17.5471369 |
| OAS1 | 4.09118178 | 8.49494286 | 10.2592462 | 4.17E-14 | 1.74E-11 | 21.9082524 |
| RP4-641G12.3 | 4.02755466 | 1.07856285 | 7.57225036 | 6.00E-10 | 5.72E-08 | 11.8673843 |
| IFITM3 | 4.01967722 | 8.83547974 | 9.043076 | 2.93E-12 | 5.61E-10 | 17.7543667 |
| SLC5A9 | 4.00879539 | 3.30789119 | 8.82980992 | 6.27E-12 | 1.02E-09 | 17.0091939 |
| RP11-466G12.3 | 3.99299924 | 0.55248304 | 7.94948465 | 1.51E-10 | 1.66E-08 | 13.1275201 |
| RMI2 | 3.9388039 | 3.20860803 | 7.60372552 | 5.35E-10 | 5.18E-08 | 12.6732467 |
| GBP1P1 | 3.93646897 | 3.00256366 | 5.37788392 | 1.80E-06 | 7.09E-05 | 4.92240757 |
| MT2A | 3.92847782 | 5.47736325 | 11.1243872 | 2.25E-15 | 1.73E-12 | 24.7498932 |
| SPATS2L | 3.91052822 | 6.06216049 | 9.09275431 | 2.45E-12 | 4.84E-10 | 17.9257932 |
| FAP | 3.8486107 | -0.4422018 | 4.69461037 | 1.98E-05 | 5.51E-04 | 2.44353954 |
| MX1 | 3.83035013 | 9.41242592 | 10.5492811 | 1.55E-14 | 7.87E-12 | 22.8351558 |
| POLR2CP1 | 3.76679199 | -1.5447288 | 4.80224332 | 1.37E-05 | 3.96E-04 | 2.2558468 |
| ZDHHC4P1 | 3.75861056 | 3.94448499 | 8.6653891 | 1.13E-11 | 1.65E-09 | 16.4102331 |
| NEURL3 | 3.7135152 | -0.1059035 | 5.7700402 | 4.41E-07 | 2.05E-05 | 5.74395252 |
| EPSTI1 | 3.67366593 | 7.62773183 | 11.0319657 | 3.06E-15 | 2.05E-12 | 24.4718022 |
| LY6E | 3.55585185 | 9.3559693 | 8.90918712 | 4.72E-12 | 8.13E-10 | 17.2883339 |
| LIPM | 3.53015972 | -0.8572704 | 4.38606178 | 5.64E-05 | 0.0012913 | 1.61351473 |
| IFI27 | 3.49511662 | 6.52857751 | 4.43610004 | 4.77E-05 | 0.00113878 | 1.68889623 |
| OAS2 | 3.49501139 | 9.04882392 | 10.1557524 | 5.95E-14 | 2.24E-11 | 21.5525381 |
| RTP4 | 3.4265898 | 5.4111897 | 10.2226041 | 4.73E-14 | 1.89E-11 | 21.7846972 |
| LAP3 | 3.39744741 | 7.05791179 | 14.4778104 | 6.73E-20 | 3.00E-16 | 34.907231 |
| SMTNL1 | 3.28159997 | 5.65340479 | 9.11420068 | 2.27E-12 | 4.54E-10 | 17.9975846 |
| DHX58 | 3.24397611 | 6.0760597 | 11.0365791 | 3.01E-15 | 2.05E-12 | 24.4913109 |
| DDX60 | 3.22335203 | 7.68361699 | 10.3357336 | 3.21E-14 | 1.43E-11 | 22.1730476 |
| IFI35 | 3.21995924 | 6.29566426 | 12.8681175 | 8.35E-18 | 1.57E-14 | 30.2405377 |
| ANKRD22 | 3.20716177 | 4.05049279 | 5.55380295 | 9.61E-07 | 4.08E-05 | 5.39188834 |
| CTD-2132N18.3 | 3.19608219 | -0.5127289 | 4.64606376 | 2.34E-05 | 6.33E-04 | 2.37814217 |
| SOCS1 | 3.14212409 | 3.48053286 | 6.63030256 | 1.91E-08 | 1.25E-06 | 9.22550176 |
| ADAMTS5 | -3.1419445 | -1.5536131 | -6.2405188 | 7.96E-08 | 4.38E-06 | 7.27089728 |
| IDO1 | 3.10829012 | 3.66040789 | 5.44328039 | 1.43E-06 | 5.83E-05 | 4.99554041 |
| SAMD9L | 3.09383792 | 9.00100623 | 9.94365602 | 1.24E-13 | 4.15E-11 | 20.8371667 |
| PTPRU | 3.09252416 | -0.3957957 | 3.6522206 | 6.04E-04 | 0.00874893 | -0.3858154 |
| IL1RN | 3.08165143 | 7.86787872 | 8.83989451 | 6.05E-12 | 9.96E-10 | 17.0425108 |
| GBP1 | 3.07841798 | 8.28332624 | 9.73633838 | 2.54E-13 | 6.82E-11 | 20.1439334 |
| RP11-477J21.2 | 3.07790744 | 1.66179467 | 6.41706615 | 4.17E-08 | 2.48E-06 | 8.46060818 |
| RP11-576C12.1 | 3.07038841 | 0.09991346 | 6.09509097 | 1.35E-07 | 6.97E-06 | 7.06170618 |
| IFIH1 | 3.05615121 | 5.16152384 | 10.2077611 | 4.98E-14 | 1.91E-11 | 21.7420567 |
| DDX58 | 3.05228273 | 8.76848247 | 9.7648766 | 2.30E-13 | 6.31E-11 | 20.2331575 |
| EIF2AK2 | 3.04531261 | 8.74048854 | 9.02525519 | 3.12E-12 | 5.86E-10 | 17.6923596 |
| RIN2 | 2.97979106 | 5.18601473 | 5.95609191 | 2.25E-07 | 1.09E-05 | 6.72794827 |
| PNPT1 | 2.96200649 | 5.75610442 | 15.7161746 | 2.04E-21 | 3.84E-17 | 38.2945451 |
| LIF | 2.96162412 | 0.1467629 | 3.83557808 | 3.40E-04 | 0.00554097 | 0.12964088 |
| AGRN | 2.96074536 | 4.14073762 | 8.7468866 | 8.44E-12 | 1.30E-09 | 16.7214739 |
| PLSCR1 | 2.93536876 | 7.7488562 | 8.53741393 | 1.79E-11 | 2.45E-09 | 15.9696999 |
| RUFY4 | 2.93362677 | 2.76315936 | 6.60235348 | 2.11E-08 | 1.36E-06 | 9.15381317 |
| RNA5SP39 | 2.92619691 | 1.65110649 | 7.92102272 | 1.68E-10 | 1.83E-08 | 13.6952142 |
| IRF7 | 2.91498532 | 7.85958316 | 9.84790875 | 1.73E-13 | 5.31E-11 | 20.5249112 |
| CD274 | 2.88900383 | 5.19443924 | 7.49777588 | 7.88E-10 | 7.22E-08 | 12.251793 |
| HERC6 | 2.88502488 | 6.78868489 | 14.4793492 | 6.70E-20 | 3.00E-16 | 34.9372389 |
| PARP14 | 2.85820829 | 10.0709856 | 10.144093 | 6.19E-14 | 2.24E-11 | 21.4683706 |
| SAMD4A | 2.85458551 | 4.63716103 | 9.73207227 | 2.58E-13 | 6.83E-11 | 20.1278692 |
| XAF1 | 2.82767522 | 8.87754195 | 10.4731797 | 2.01E-14 | 9.67E-12 | 22.6145676 |
| UBE2L6 | 2.81585917 | 8.04563717 | 12.1290789 | 8.53E-17 | 1.23E-13 | 27.9539733 |
| IFIT5 | 2.8081434 | 7.27915175 | 8.58007019 | 1.54E-11 | 2.12E-09 | 16.1143844 |
| RPL12P42 | 2.80713805 | -1.2840944 | 5.80561357 | 3.88E-07 | 1.82E-05 | 5.52762927 |
| FBXO39 | 2.78660174 | 1.93155647 | 5.78045273 | 4.25E-07 | 1.98E-05 | 6.26924042 |
| TREX1 | 2.78335116 | 5.35628404 | 11.2943685 | 1.28E-15 | 1.09E-12 | 25.3285885 |
| RP11-820K3.2 | 2.78060517 | 1.85978069 | 8.83514841 | 6.15E-12 | 1.00E-09 | 16.8163977 |
| ZBP1 | 2.7590597 | 7.80617961 | 10.5830066 | 1.38E-14 | 7.25E-12 | 22.9932223 |
| AC007899.3 | 2.7590387 | 2.60500859 | 7.70875966 | 3.64E-10 | 3.72E-08 | 13.0527767 |
| MTATP6P26 | 2.75795549 | 1.18368267 | 5.63792846 | 7.10E-07 | 3.11E-05 | 5.79275806 |
| TNFAIP6 | 2.74561128 | 5.67688404 | 6.94939898 | 5.91E-09 | 4.40E-07 | 10.2585333 |
| AC074338.4 | 2.73835156 | 2.56108997 | 8.46640009 | 2.32E-11 | 3.06E-09 | 15.7102685 |
| CARD17 | 2.73004895 | 0.50468438 | 3.99760189 | 2.02E-04 | 0.00369148 | 0.61407234 |
| GBP4 | 2.70490517 | 7.89037438 | 8.8478572 | 5.88E-12 | 9.77E-10 | 17.0641124 |
| RP11-365F18.6 | 2.70402059 | -1.6449969 | 5.03124375 | 6.15E-06 | 1.98E-04 | 3.26860941 |
| MDK | 2.69992275 | 1.48244713 | 5.54441374 | 9.94E-07 | 4.21E-05 | 5.4868867 |
| ACO1 | 2.69133751 | 7.58904119 | 10.0788282 | 7.76E-14 | 2.70E-11 | 21.308234 |
| LIPA | 2.67698151 | 6.62085137 | 9.54150174 | 5.03E-13 | 1.18E-10 | 19.4710116 |
| SF3A3P2 | 2.67073535 | -0.4201798 | 4.66030355 | 2.23E-05 | 6.08E-04 | 2.44147579 |
| PLSCR2 | 2.64557365 | 3.25711476 | 7.89538289 | 1.84E-10 | 1.99E-08 | 13.724632 |
| TFEC | 2.63436714 | 6.2786755 | 7.10575663 | 3.32E-09 | 2.60E-07 | 10.8175555 |
| RPL37P6 | 2.60956117 | 1.39504354 | 6.4196957 | 4.13E-08 | 2.46E-06 | 8.39961012 |
| PML | 2.60674698 | 8.28776 | 11.608425 | 4.58E-16 | 5.37E-13 | 26.3021489 |
| PARP12 | 2.59609495 | 7.68605972 | 12.9697717 | 6.10E-18 | 1.27E-14 | 30.5290199 |
| MTCO3P43 | 2.59185076 | 0.73033391 | 5.19186344 | 3.49E-06 | 1.24E-04 | 4.29492531 |
| STAT2 | 2.580259 | 8.13532584 | 9.78638783 | 2.14E-13 | 6.08E-11 | 20.3140918 |
| RGL1 | 2.57440097 | 3.57931068 | 7.00779967 | 4.77E-09 | 3.65E-07 | 10.5611808 |
| TNFSF10 | 2.57363065 | 8.89749434 | 8.95248475 | 4.04E-12 | 7.23E-10 | 17.4386517 |
| RP1-145M24.1 | 2.54387061 | -0.9120701 | 5.27338299 | 2.62E-06 | 9.66E-05 | 4.39535443 |
| TCN2 | 2.53636103 | 3.439782 | 5.11837344 | 4.53E-06 | 1.54E-04 | 3.96082208 |
| TNFSF13B | 2.53470884 | 7.47993863 | 7.81721946 | 2.45E-10 | 2.55E-08 | 13.4056156 |
| SCO2 | 2.53048376 | 4.07252217 | 6.58516001 | 2.25E-08 | 1.44E-06 | 9.03450028 |
| ANO7L1 | 2.52905119 | 1.62046811 | 5.75325189 | 4.68E-07 | 2.17E-05 | 6.20674191 |
| BST2 | 2.52427124 | 6.26707922 | 11.0573333 | 2.81E-15 | 2.03E-12 | 24.5647691 |
| TRIM6 | 2.51867168 | 3.12165067 | 6.84627921 | 8.63E-09 | 6.09E-07 | 10.0052222 |
| FRMD3 | 2.5081055 | 4.62157375 | 6.01418032 | 1.82E-07 | 9.10E-06 | 6.96340989 |
| KIAA1958 | 2.50482127 | 5.31599908 | 8.69468526 | 1.02E-11 | 1.52E-09 | 16.5183349 |
| HELZ2 | 2.49998114 | 8.42750677 | 9.12414661 | 2.20E-12 | 4.43E-10 | 18.035966 |
| TTC21A | 2.4975464 | 4.70904026 | 8.80827426 | 6.77E-12 | 1.07E-09 | 16.9222338 |
| GPR42 | 2.48010676 | -1.3415914 | 4.51202645 | 3.69E-05 | 9.19E-04 | 1.99658699 |
| PARP9 | 2.47587016 | 8.81203982 | 8.93243729 | 4.34E-12 | 7.55E-10 | 17.3691636 |
| PRM3 | 2.47143039 | -1.3179885 | 6.66296119 | 1.69E-08 | 1.13E-06 | 8.15418007 |
| FAM46A | 2.43797051 | 6.48028836 | 9.67651958 | 3.13E-13 | 7.95E-11 | 19.9348011 |
| MTND3P10 | 2.43726757 | -0.5041549 | 4.38831863 | 5.60E-05 | 0.00128314 | 1.6985341 |
| TTC26 | 2.43466075 | 3.39918635 | 7.83473822 | 2.30E-10 | 2.41E-08 | 13.5020414 |
| FBXO6 | 2.42837581 | 5.06499516 | 8.45194697 | 2.44E-11 | 3.18E-09 | 15.6667951 |
| AC003080.4 | 2.42605129 | 0.734552 | 6.17262005 | 1.02E-07 | 5.50E-06 | 7.60340817 |
| STAT1 | 2.40705993 | 8.44139426 | 10.0002893 | 1.02E-13 | 3.47E-11 | 21.0384309 |
| FFAR3 | 2.40700692 | 1.44359469 | 4.84169479 | 1.19E-05 | 3.52E-04 | 3.1421654 |
| DOCK4 | 2.39996759 | 5.74370591 | 6.72038856 | 1.37E-08 | 9.36E-07 | 9.42062584 |
| DDX60L | 2.3988249 | 9.19382912 | 8.65441448 | 1.18E-11 | 1.67E-09 | 16.398022 |
| TRIM22 | 2.39090655 | 9.56848714 | 10.9418807 | 4.13E-15 | 2.59E-12 | 24.1114065 |
| HEY1 | -2.3833826 | -1.9718792 | -3.8171336 | 3.60E-04 | 0.00577666 | 0.00213206 |
| CETP | 2.38309904 | 4.21550005 | 10.3240231 | 3.34E-14 | 1.43E-11 | 22.1210702 |
| CXCL6 | -2.3797796 | -1.4948266 | -4.133127 | 1.30E-04 | 0.00259009 | 0.86535522 |
| EPHB2 | 2.3759869 | 3.54064975 | 5.49939649 | 1.17E-06 | 4.89E-05 | 5.25891987 |
| TDRD7 | 2.375401 | 6.41302389 | 10.373056 | 2.83E-14 | 1.29E-11 | 22.299757 |
| TOR1B | 2.3694607 | 6.46806573 | 8.61687898 | 1.35E-11 | 1.89E-09 | 16.2340465 |
| SAMD9 | 2.36372748 | 8.68924745 | 7.8421173 | 2.24E-10 | 2.37E-08 | 13.5155023 |
| TRIM5 | 2.35155284 | 6.63407514 | 10.2495241 | 4.31E-14 | 1.76E-11 | 21.8845153 |
| OR52V1P | 2.35045463 | -0.5817499 | 4.13383091 | 1.30E-04 | 0.00258686 | 0.92501891 |
| AIM2 | 2.3363572 | 4.85562615 | 7.51938868 | 7.28E-10 | 6.70E-08 | 12.3354929 |
| CEACAM1 | 2.32703303 | 6.21907874 | 7.5794275 | 5.84E-10 | 5.60E-08 | 12.520452 |
| PRLR | 2.30932169 | 2.8164814 | 6.2992189 | 6.42E-08 | 3.60E-06 | 8.07635943 |
| JUP | 2.3079716 | 5.25224482 | 4.66427253 | 2.20E-05 | 6.00E-04 | 2.25468266 |
| RP11-686D22.7 | 2.30599359 | 2.82891699 | 8.96273969 | 3.90E-12 | 7.05E-10 | 17.432848 |
| APOL6 | 2.30102252 | 9.21812922 | 11.451062 | 7.66E-16 | 7.99E-13 | 25.7614309 |
| DNMT3L | -2.2998967 | -0.3699328 | -3.770575 | 4.17E-04 | 0.00646031 | -0.0285125 |
| MYOF | 2.29892827 | 5.12297532 | 7.65975137 | 4.35E-10 | 4.28E-08 | 12.8348796 |
| AC067945.3 | 2.29845517 | 3.20142525 | 9.79337118 | 2.09E-13 | 6.03E-11 | 20.291685 |
| ACTG2 | 2.29682259 | 1.80775048 | 7.37660566 | 1.23E-09 | 1.08E-07 | 11.8666598 |
| AC007919.18 | 2.29633948 | 1.27230626 | 6.03026294 | 1.71E-07 | 8.65E-06 | 7.1547397 |
| CTD-2521M24.13 | 2.29172441 | 1.7816248 | 6.99392067 | 5.01E-09 | 3.79E-07 | 10.4810232 |
| RP1-228H13.2 | 2.28309153 | -0.9081868 | 3.91836942 | 2.61E-04 | 0.00449533 | 0.31462529 |
| RP1-102G20.2 | 2.27546437 | -0.4298822 | 5.33789443 | 2.08E-06 | 8.06E-05 | 4.57979331 |
| PARP10 | 2.2649929 | 7.17152989 | 9.76306284 | 2.32E-13 | 6.31E-11 | 20.2339916 |
| GBP5 | 2.2592889 | 9.02544093 | 6.72214204 | 1.36E-08 | 9.34E-07 | 9.5057099 |
| MTND4P26 | 2.23696698 | 1.35267628 | 5.39773438 | 1.68E-06 | 6.69E-05 | 5.00707406 |
| RSPH14 | -2.2360512 | -0.9330718 | -4.4225258 | 4.99E-05 | 0.001177 | 1.8258005 |
| CDC42P3 | 2.22500479 | -0.8968121 | 5.4621623 | 1.34E-06 | 5.47E-05 | 4.56329839 |
| ANXA10 | 2.22139365 | -0.112672 | 4.03972969 | 1.77E-04 | 0.0033047 | 0.71347233 |
| LGALS9 | 2.21379071 | 7.9521542 | 9.57749213 | 4.43E-13 | 1.07E-10 | 19.6004637 |
| CXCL8 | -2.2098596 | 2.25224876 | -3.6639635 | 5.82E-04 | 0.00848294 | -0.5101406 |
| ZCCHC2 | 2.20598423 | 8.29266346 | 9.18571169 | 1.76E-12 | 3.68E-10 | 18.2486103 |
| OR52B6 | 2.20181208 | -0.5613698 | 5.01979542 | 6.40E-06 | 2.06E-04 | 3.49974802 |
| 4-Sep | 2.19222032 | 4.45597469 | 5.66119874 | 6.53E-07 | 2.88E-05 | 5.70901725 |
| MAFB | 2.19128598 | 6.02979127 | 6.65099464 | 1.77E-08 | 1.17E-06 | 9.16582592 |
| FCGR1A | 2.1595476 | 6.04678642 | 4.96353253 | 7.80E-06 | 2.44E-04 | 3.215216 |
| RNF213 | 2.15256969 | 11.2863453 | 8.72481204 | 9.13E-12 | 1.39E-09 | 16.6061421 |
| OR52T1P | 2.14768681 | -0.1468784 | 4.51238753 | 3.68E-05 | 9.19E-04 | 2.08604885 |
| NTNG2 | 2.14745141 | 7.72401466 | 6.67302624 | 1.63E-08 | 1.09E-06 | 9.28922192 |
| LGALS9B | 2.14725268 | 2.127297 | 7.3571856 | 1.32E-09 | 1.15E-07 | 11.8069181 |
| OLFM5P | 2.13366094 | 0.67130987 | 4.85568156 | 1.13E-05 | 3.38E-04 | 3.22131806 |
| RBM17P2 | -2.1310639 | -2.1246294 | -4.7221233 | 1.80E-05 | 5.05E-04 | 2.61149272 |
| TGM3 | -2.1240781 | 0.72309211 | -3.889906 | 0.000286 | 0.00481276 | 0.24084302 |
| RGS1 | 2.11607699 | 1.59479175 | 6.26611896 | 7.25E-08 | 4.03E-06 | 7.96956184 |
| KAT2A | 2.11581881 | 5.13830469 | 10.1490053 | 6.09E-14 | 2.24E-11 | 21.5458227 |
| VSIG10L | 2.10757515 | 0.59080645 | 5.89802195 | 2.77E-07 | 1.33E-05 | 6.61813512 |
| LMO2 | 2.09949217 | 6.31714886 | 8.5366687 | 1.80E-11 | 2.45E-09 | 15.9464768 |
| FAM72A | 2.09749846 | 2.15994741 | 8.10401016 | 8.61E-11 | 1.01E-08 | 14.4122325 |
| UBQLNL | 2.09569047 | 0.63955971 | 4.86468658 | 1.10E-05 | 3.31E-04 | 3.24777359 |
| C19orf66 | 2.09198134 | 6.4728434 | 12.6733943 | 1.53E-17 | 2.61E-14 | 29.6723828 |
| IFITM1 | 2.09158535 | 9.31421091 | 9.60367637 | 4.04E-13 | 9.86E-11 | 19.6782665 |
| SDC2 | -2.0870462 | 0.07627288 | -3.7248066 | 4.82E-04 | 0.00727083 | -0.1944593 |
| FCGR1CP | 2.08566691 | 2.96309195 | 4.25759776 | 8.65E-05 | 0.00184401 | 1.1469114 |
| MTND5P28 | 2.08358869 | 2.70273962 | 6.2608481 | 7.39E-08 | 4.09E-06 | 7.93401682 |
| ABTB2 | 2.06647501 | 4.3288127 | 7.252092 | 1.94E-09 | 1.63E-07 | 11.3663335 |
| XXbac-BCX360G3.2 | 2.06244634 | -0.2522815 | 6.57670883 | 2.32E-08 | 1.46E-06 | 8.52038828 |
| XXbac-BPG554J19.2 | 2.06244634 | -0.2522815 | 6.57670883 | 2.32E-08 | 1.46E-06 | 8.52038828 |
| LLpac-136A2.1 | 2.06242573 | -1.0949869 | 4.54034069 | 3.35E-05 | 8.57E-04 | 2.13968389 |
| TMEM51 | 2.0624197 | 0.63669468 | 4.23419526 | 9.35E-05 | 0.00195694 | 1.30847364 |
| TIMM10 | 2.06144631 | 4.01964348 | 5.62287999 | 7.50E-07 | 3.27E-05 | 5.63960138 |
| APOL1 | 2.05918249 | 6.12310666 | 8.63396384 | 1.27E-11 | 1.79E-09 | 16.289337 |
| PPP1R2P1 | 2.05131004 | -0.3497927 | 5.48108436 | 1.25E-06 | 5.17E-05 | 4.96456942 |
| SRGAP2 | 2.04465183 | 7.77075835 | 10.7705536 | 7.35E-15 | 4.31E-12 | 23.6124706 |
| DRAP1 | 2.04455003 | 5.90634601 | 13.2987284 | 2.23E-18 | 5.23E-15 | 31.5575603 |
| PI4K2B | 2.02968903 | 4.83065353 | 11.5094386 | 6.32E-16 | 6.99E-13 | 26.0122929 |
| RP11-686D22.3 | 2.02956083 | 2.31980058 | 7.4080267 | 1.10E-09 | 9.75E-08 | 11.9893092 |
| TMEM123 | 2.01961594 | 9.06342047 | 8.58335708 | 1.52E-11 | 2.11E-09 | 16.147794 |
| ZNF366 | 2.01933888 | 3.00753432 | 5.95474121 | 2.26E-07 | 1.09E-05 | 6.82515388 |
| CASP5 | 2.01878321 | 4.01877905 | 4.84529284 | 1.18E-05 | 3.48E-04 | 2.89959205 |
| HSPA8P5 | 2.00932445 | -0.1839161 | 6.10914785 | 1.29E-07 | 6.71E-06 | 7.00362449 |
| GRAMD1B | 2.00912368 | 5.02339407 | 6.58095553 | 2.29E-08 | 1.45E-06 | 8.93907057 |
| NT5C3A | 2.00351522 | 6.02936891 | 10.4530064 | 2.15E-14 | 1.01E-11 | 22.5672054 |
| CLDN18 | 2.00325671 | 1.57463788 | 6.1571111 | 1.08E-07 | 5.76E-06 | 7.5977271 |

**Supplemental Table 3.** Differentially expressed genes day 8 post infection

| **geneID** | **logFC** | **AveExpr** | **t** | **P.Value** | **adj.P.Val** | **B** |
| --- | --- | --- | --- | --- | --- | --- |
| IFI27 | 7.93571612 | 6.52857751 | 9.31659349 | 1.11E-12 | 2.17E-10 | 18.6147622 |
| CCL2 | 7.54361689 | 1.13589506 | 4.64026024 | 2.38E-05 | 3.72E-04 | 2.36345751 |
| CCL8 | 7.13246682 | -0.582129 | 4.45439268 | 4.48E-05 | 6.20E-04 | 1.40353009 |
| CXCL10 | 6.81109105 | 3.2144798 | 5.6731139 | 6.26E-07 | 1.86E-05 | 5.91396139 |
| PALM2-AKAP2 | 6.60888495 | -2.3304446 | 3.74057062 | 4.58E-04 | 0.00404694 | -0.6197259 |
| OTOF | 6.21560731 | 3.39961896 | 3.58671214 | 7.39E-04 | 0.0058556 | -0.6525768 |
| SIGLEC1 | 6.16027959 | 6.35550789 | 9.65168508 | 3.42E-13 | 8.92E-11 | 19.821639 |
| USP18 | 6.10115899 | 5.2942684 | 12.5769433 | 2.07E-17 | 4.71E-14 | 29.0455775 |
| HESX1 | 5.82448045 | -1.1411462 | 5.22681228 | 3.09E-06 | 7.04E-05 | 3.38205975 |
| IFI44L | 5.74017998 | 9.26563843 | 10.4843608 | 1.93E-14 | 9.56E-12 | 22.6105865 |
| RSAD2 | 5.72575943 | 8.78506662 | 9.56468887 | 4.64E-13 | 1.12E-10 | 19.5408519 |
| C1QC | 5.44618005 | 0.86433275 | 3.87558942 | 2.99E-04 | 0.00291731 | 0.24038188 |
| USP41 | 5.27905779 | 1.09222199 | 7.50804214 | 7.59E-10 | 5.99E-08 | 11.3547651 |
| ADAMTS5 | -5.2298488 | -1.5536131 | -6.6198587 | 1.98E-08 | 1.01E-06 | 6.55577692 |
| CXCL11 | 5.21935843 | -1.12088 | 3.988552 | 2.08E-04 | 0.00218201 | 0.10677166 |
| SPATS2L | 5.14681026 | 6.06216049 | 11.4429455 | 7.86E-16 | 8.20E-13 | 25.7416084 |
| IFIT1 | 5.10209124 | 8.99783691 | 9.12207256 | 2.21E-12 | 3.78E-10 | 18.0215636 |
| POLR2CP1 | 4.85498487 | -1.5447288 | 6.4092121 | 4.29E-08 | 1.91E-06 | 6.52523008 |
| PI3 | -4.8480812 | 2.47364729 | -4.6939591 | 1.98E-05 | 3.17E-04 | 2.71532537 |
| KCTD14 | 4.79437387 | 0.65016798 | 5.63123501 | 7.28E-07 | 2.11E-05 | 5.40020623 |
| IFIT3 | 4.78077263 | 9.34813991 | 9.31264902 | 1.13E-12 | 2.17E-10 | 18.6716309 |
| CMPK2 | 4.77588583 | 7.47598457 | 10.1227035 | 6.67E-14 | 2.61E-11 | 21.4467531 |
| IFI44 | 4.76815237 | 7.7459889 | 10.28217 | 3.86E-14 | 1.72E-11 | 21.9800376 |
| OAS3 | 4.76177853 | 9.74421226 | 9.62935628 | 3.70E-13 | 9.51E-11 | 19.7444241 |
| SLC31A1P1 | 4.75528449 | -2.106749 | 6.26599388 | 7.25E-08 | 2.98E-06 | 5.85832467 |
| HES4 | 4.67870658 | 2.18929656 | 6.47628403 | 3.36E-08 | 1.57E-06 | 8.63537378 |
| SERPING1 | 4.66865848 | 5.97008766 | 8.61733938 | 1.34E-11 | 1.93E-09 | 16.2677382 |
| NAT8 | 4.65322902 | 1.16331174 | 4.17438398 | 1.14E-04 | 0.00132705 | 1.13117387 |
| TMPRSS2 | 4.5888233 | -1.0412947 | 5.2974385 | 2.40E-06 | 5.69E-05 | 3.49926257 |
| EXOC3L1 | 4.53034066 | 2.25582378 | 7.67029681 | 4.19E-10 | 3.56E-08 | 12.7024431 |
| C1QB | 4.5112678 | 1.39849469 | 4.67269934 | 2.13E-05 | 3.36E-04 | 2.63839384 |
| ISG15 | 4.50149643 | 0.3381444 | 5.47471009 | 1.28E-06 | 3.34E-05 | 4.87280466 |
| IFI6 | 4.49096606 | 8.08614509 | 10.5878582 | 1.36E-14 | 7.30E-12 | 22.9931174 |
| IFITM3 | 4.46533649 | 8.83547974 | 9.81164204 | 1.96E-13 | 5.93E-11 | 20.3853709 |
| OAS1 | 4.46129887 | 8.49494286 | 10.9702589 | 3.76E-15 | 2.80E-12 | 24.2344583 |
| MT2A | 4.39187421 | 5.47736325 | 12.2753117 | 5.36E-17 | 8.39E-14 | 28.3462733 |
| OASL | 4.33984711 | 7.54938765 | 10.9722552 | 3.74E-15 | 2.80E-12 | 24.2594641 |
| ALMS1P1 | 4.32401628 | 1.07374967 | 4.2050606 | 1.03E-04 | 0.00121698 | 1.2221753 |
| ETV7 | 4.31916073 | 4.46985866 | 7.30410932 | 1.60E-09 | 1.12E-07 | 11.6254383 |
| CH17-296N19.1 | -4.3136851 | 2.01758359 | -6.8210037 | 9.47E-09 | 5.35E-07 | 9.84472441 |
| CCNA1 | 4.2890589 | -0.0846615 | 4.35666648 | 6.22E-05 | 8.09E-04 | 1.56829402 |
| IFIT2 | 4.27025992 | 9.66057711 | 9.1404658 | 2.07E-12 | 3.57E-10 | 18.0704527 |
| ATF3 | 4.25747354 | 2.72288822 | 5.98816045 | 2.00E-07 | 7.09E-06 | 7.01423781 |
| HERC5 | 4.24866831 | 7.68061448 | 9.59296957 | 4.20E-13 | 1.02E-10 | 19.6525117 |
| BATF2 | 4.24744752 | 5.07853875 | 8.08222039 | 9.32E-11 | 1.02E-08 | 14.3840129 |
| OVOL1 | 4.2137612 | -2.2541658 | 6.12764051 | 1.20E-07 | 4.60E-06 | 5.58648336 |
| NKAIN1 | 4.1245452 | -3.17844 | 4.30455894 | 7.40E-05 | 9.36E-04 | 0.57323746 |
| NEURL3 | 4.11134893 | -0.1059035 | 6.45725204 | 3.60E-08 | 1.65E-06 | 7.80169861 |
| RIN2 | 4.09823023 | 5.18601473 | 7.93158372 | 1.61E-10 | 1.56E-08 | 13.8335768 |
| LY6E | 4.04377039 | 9.3559693 | 9.85978979 | 1.66E-13 | 5.18E-11 | 20.5412053 |
| EPSTI1 | 4.035266 | 7.62773183 | 11.9147795 | 1.70E-16 | 2.26E-13 | 27.2736712 |
| RP4-641G12.3 | 4.01186187 | 1.07856285 | 7.53859812 | 6.79E-10 | 5.46E-08 | 11.6929314 |
| ZDHHC4P1 | 4.01165523 | 3.94448499 | 9.2766791 | 1.28E-12 | 2.35E-10 | 18.4943174 |
| GBP1P1 | 3.97070675 | 3.00256366 | 5.42782685 | 1.51E-06 | 3.82E-05 | 5.09690912 |
| RTP4 | 3.96791805 | 5.4111897 | 11.6951284 | 3.45E-16 | 3.81E-13 | 26.5504367 |
| RBM17P2 | -3.9110239 | -2.1246294 | -6.1096823 | 1.28E-07 | 4.87E-06 | 5.56980103 |
| PTPRU | 3.90080694 | -0.3957957 | 4.75724586 | 1.60E-05 | 2.69E-04 | 2.64189994 |
| LAMP3 | 3.89667337 | 4.21810844 | 9.80537368 | 2.00E-13 | 5.94E-11 | 20.3392392 |
| FAP | 3.88703538 | -0.4422018 | 4.75681148 | 1.60E-05 | 2.69E-04 | 2.5902719 |
| MX1 | 3.85466434 | 9.41242592 | 10.6274228 | 1.19E-14 | 6.58E-12 | 23.0767824 |
| PHF24 | -3.8427842 | -1.2092825 | -4.0497834 | 1.71E-04 | 0.00184887 | 0.57225195 |
| OAS2 | 3.84124811 | 9.04882392 | 10.9614804 | 3.87E-15 | 2.80E-12 | 24.1851625 |
| RMI2 | 3.83961797 | 3.20860803 | 7.3907031 | 1.17E-09 | 8.53E-08 | 11.9187585 |
| RP11-477J21.2 | 3.8066012 | 1.66179467 | 8.12858809 | 7.87E-11 | 8.85E-09 | 14.2972499 |
| LIF | 3.80298081 | 0.1467629 | 5.07193344 | 5.33E-06 | 1.10E-04 | 3.67364343 |
| TCN2 | 3.79969255 | 3.439782 | 7.93252235 | 1.61E-10 | 1.56E-08 | 13.8548535 |
| OTX1 | -3.7774953 | 0.45074125 | -5.6818601 | 6.06E-07 | 1.81E-05 | 5.67402169 |
| CARD17 | 3.72308239 | 0.50468438 | 5.65828774 | 6.60E-07 | 1.94E-05 | 5.687002 |
| NMRAL1P1 | 3.69884765 | -0.3926125 | 3.93386645 | 2.49E-04 | 0.00252 | 0.37220065 |
| CTD-2132N18.3 | 3.62221295 | -0.5127289 | 5.35665836 | 1.95E-06 | 4.73E-05 | 4.46256863 |
| DZIP1L | 3.6191717 | 1.33912248 | 4.17677928 | 1.13E-04 | 0.0013191 | 1.11406503 |
| RP11-466G12.3 | 3.59776017 | 0.55248304 | 7.08665657 | 3.57E-09 | 2.33E-07 | 10.2801465 |
| C1QA | 3.58284536 | 2.17143186 | 5.19412865 | 3.47E-06 | 7.75E-05 | 4.32004262 |
| RUFY4 | 3.57767493 | 2.76315936 | 8.2292082 | 5.46E-11 | 6.58E-09 | 14.8470317 |
| RP11-290L7.3 | -3.5705918 | -3.3051161 | -3.8228468 | 3.54E-04 | 0.00330302 | -0.5148554 |
| FBXO39 | 3.54973501 | 1.93155647 | 7.55741684 | 6.33E-10 | 5.17E-08 | 12.2733784 |
| DDX60 | 3.53151448 | 7.68361699 | 11.1647747 | 1.97E-15 | 1.75E-12 | 24.8930272 |
| HEY1 | -3.5279267 | -1.9718792 | -4.4659694 | 4.31E-05 | 6.01E-04 | 1.29411567 |
| MDK | 3.48178013 | 1.48244713 | 7.360218 | 1.31E-09 | 9.41E-08 | 11.6341093 |
| METTL7B | 3.45347066 | -1.417302 | 4.32869645 | 6.83E-05 | 8.75E-04 | 1.24440553 |
| SMTNL1 | 3.4513138 | 5.65340479 | 9.54482267 | 4.97E-13 | 1.15E-10 | 19.4882201 |
| RBM17P1 | -3.4392139 | -0.2524074 | -5.4393783 | 1.45E-06 | 3.70E-05 | 4.92829328 |
| SLC5A9 | 3.39434581 | 3.30789119 | 7.47722059 | 8.50E-10 | 6.62E-08 | 12.2360591 |
| CTSL | 3.3837642 | 2.9198531 | 8.0479092 | 1.06E-10 | 1.13E-08 | 14.2539811 |
| TFEC | 3.35120738 | 6.2786755 | 8.77715172 | 7.57E-12 | 1.16E-09 | 16.8160277 |
| TEKT1 | 3.34419743 | -1.6906615 | 3.90989262 | 2.68E-04 | 0.0026861 | -0.0220245 |
| CLDN23 | 3.32737605 | -0.8399016 | 4.01771665 | 1.90E-04 | 0.00201414 | 0.60142935 |
| PAX3 | 3.29938296 | -1.0489676 | 4.22828625 | 9.53E-05 | 0.0011467 | 1.09931244 |
| SAMD4A | 3.28408518 | 4.63716103 | 11.1523603 | 2.05E-15 | 1.75E-12 | 24.8294437 |
| EPHB2 | 3.26394438 | 3.54064975 | 7.78402662 | 2.76E-10 | 2.45E-08 | 13.3305536 |
| ARPP21 | -3.2591741 | -2.2976918 | -3.536658 | 8.61E-04 | 0.00660385 | -0.9008702 |
| CXCR2P1 | 3.25006171 | 6.08080437 | 9.19862948 | 1.69E-12 | 3.01E-10 | 18.2896357 |
| FRMD3 | 3.23338927 | 4.62157375 | 7.75145603 | 3.11E-10 | 2.72E-08 | 13.1949151 |
| LAP3 | 3.22132264 | 7.05791179 | 13.882758 | 3.85E-19 | 2.41E-15 | 33.2068856 |
| MS4A4A | 3.19953526 | 2.9589409 | 7.47540988 | 8.56E-10 | 6.64E-08 | 12.2376613 |
| PNMA6A | -3.1982332 | -1.2319706 | -5.5431315 | 9.99E-07 | 2.73E-05 | 4.68330866 |
| CDKN1C | 3.19546352 | 3.41914583 | 7.28502206 | 1.72E-09 | 1.19E-07 | 11.538292 |
| PLSCR1 | 3.19300464 | 7.7488562 | 9.17905992 | 1.81E-12 | 3.17E-10 | 18.225406 |
| DHX58 | 3.1600681 | 6.0760597 | 10.8020111 | 6.61E-15 | 4.00E-12 | 23.7190856 |
| C15orf48 | 3.15693707 | -1.2056301 | 4.61980411 | 2.56E-05 | 3.96E-04 | 2.20183435 |
| EIF2AK2 | 3.15357052 | 8.74048854 | 9.30934624 | 1.14E-12 | 2.17E-10 | 18.6746643 |
| IRF7 | 3.14196968 | 7.85958316 | 10.5060045 | 1.80E-14 | 9.37E-12 | 22.7316206 |
| PKD2L1 | 3.133964 | -1.8891684 | 3.37497395 | 0.00140088 | 0.00959116 | -1.1577921 |
| JUP | 3.12582176 | 5.25224482 | 6.21112262 | 8.86E-08 | 3.53E-06 | 7.64311024 |
| TMEM255A | 3.10885745 | 2.31940815 | 8.62459922 | 1.31E-11 | 1.89E-09 | 16.2105988 |
| HERC6 | 3.1055033 | 6.78868489 | 15.4525216 | 4.24E-21 | 7.96E-17 | 37.5588611 |
| RPL37P6 | 3.10384634 | 1.39504354 | 7.78766964 | 2.73E-10 | 2.43E-08 | 12.9868963 |
| RP11-576C12.1 | 3.1031831 | 0.09991346 | 6.165972 | 1.05E-07 | 4.05E-06 | 7.25004934 |
| IFI35 | 3.09100003 | 6.29566426 | 12.4408764 | 3.17E-17 | 5.42E-14 | 28.9304295 |
| SAMD9L | 3.08189784 | 9.00100623 | 9.93658208 | 1.27E-13 | 4.33E-11 | 20.8071248 |
| IL1RN | 3.05811252 | 7.86787872 | 8.81000334 | 6.73E-12 | 1.04E-09 | 16.9400602 |
| UBE2L6 | 3.02239814 | 8.04563717 | 12.9052041 | 7.45E-18 | 2.00E-14 | 30.2938208 |
| GBP1 | 3.01641459 | 8.28332624 | 9.59611461 | 4.15E-13 | 1.02E-10 | 19.663648 |
| RP1-228H13.2 | 2.99395442 | -0.9081868 | 5.35568116 | 1.95E-06 | 4.74E-05 | 4.42188335 |
| ANXA10 | 2.99265127 | -0.112672 | 5.68311232 | 6.03E-07 | 1.80E-05 | 5.65943603 |
| SCO2 | 2.990905 | 4.07252217 | 7.8541137 | 2.14E-10 | 1.94E-08 | 13.5736444 |
| XAF1 | 2.9585729 | 8.87754195 | 10.9018877 | 4.73E-15 | 3.29E-12 | 24.0084864 |
| ALPL | -2.9582056 | 5.98271276 | -5.1655237 | 3.83E-06 | 8.40E-05 | 3.95505137 |
| RGL1 | 2.95429048 | 3.57931068 | 8.12964421 | 7.84E-11 | 8.85E-09 | 14.552283 |
| TMEM51 | 2.9470934 | 0.63669468 | 6.31548829 | 6.05E-08 | 2.56E-06 | 7.94660862 |
| DDX58 | 2.9414674 | 8.76848247 | 9.49269469 | 5.97E-13 | 1.34E-10 | 19.3025424 |
| ANO5 | 2.93625963 | 0.30995233 | 4.157002 | 1.21E-04 | 0.00139079 | 1.07939423 |
| RSPH14 | -2.9314778 | -0.9330718 | -4.9588324 | 7.93E-06 | 1.52E-04 | 3.23186684 |
| GBP4 | 2.92950461 | 7.89037438 | 9.49209225 | 5.98E-13 | 1.34E-10 | 19.3071423 |
| NAT8B | 2.91861139 | 0.97581383 | 4.13595131 | 1.29E-04 | 0.00146798 | 1.00897191 |
| TRIM6 | 2.91439833 | 3.12165067 | 8.04328307 | 1.07E-10 | 1.15E-08 | 14.2401827 |
| MTND4P22 | -2.9077502 | -0.4789305 | -4.6031097 | 2.71E-05 | 4.13E-04 | 2.3434092 |
| RNASE1 | 2.90625328 | -0.0541436 | 3.63602218 | 6.35E-04 | 0.00520666 | -0.3867013 |
| IFIH1 | 2.90591356 | 5.16152384 | 9.73993593 | 2.51E-13 | 6.94E-11 | 20.1562295 |
| CCRL2 | 2.90088962 | 3.41405055 | 7.09505585 | 3.46E-09 | 2.28E-07 | 10.8836491 |
| TIMM10 | 2.86907944 | 4.01964348 | 8.02425075 | 1.15E-10 | 1.21E-08 | 14.1780215 |
| SOAT2 | -2.8654549 | -1.5411279 | -4.2069468 | 1.02E-04 | 0.0012115 | 0.78619203 |
| KISS1R | -2.8482282 | -0.7066817 | -4.1345708 | 1.30E-04 | 0.00147285 | 0.85628136 |
| AC213203.1 | -2.8346706 | -1.4613119 | -3.896284 | 2.80E-04 | 0.0027846 | 0.08534554 |
| HNRNPCP4 | 2.83064283 | -2.0770287 | 3.44450855 | 0.00113779 | 0.00819002 | -1.1180737 |
| RP11-320N7.1 | 2.82835265 | -1.5519437 | 4.64215654 | 2.37E-05 | 3.69E-04 | 2.03084281 |
| MCRIP2P1 | 2.82487654 | -1.6447443 | 4.73103414 | 1.75E-05 | 2.88E-04 | 2.33727652 |
| RP5-892F13.2 | -2.8173095 | -1.6083111 | -4.6040388 | 2.70E-05 | 4.13E-04 | 1.94763311 |
| 4-Sep | 2.81193035 | 4.45597469 | 7.29018423 | 1.69E-09 | 1.17E-07 | 11.5361385 |
| OR52V1P | 2.80178535 | -0.5817499 | 5.04229517 | 5.92E-06 | 1.19E-04 | 3.52956079 |
| MYOF | 2.80033137 | 5.12297532 | 9.27194469 | 1.30E-12 | 2.37E-10 | 18.5467472 |
| ALOX15 | -2.7911284 | 2.11540222 | -4.9736653 | 7.52E-06 | 1.46E-04 | 3.59701462 |
| SF3A3P2 | 2.77048973 | -0.4201798 | 4.87009648 | 1.08E-05 | 1.96E-04 | 3.03567067 |
| XXbac-BPG541D20.6 | 2.76295533 | 0.06119171 | 6.45229882 | 3.67E-08 | 1.67E-06 | 8.26033025 |
| NECAB2 | -2.7534824 | 1.14246833 | -5.6849963 | 5.99E-07 | 1.79E-05 | 5.95592311 |
| SOCS1 | 2.75014769 | 3.48053286 | 5.7286637 | 5.12E-07 | 1.56E-05 | 6.05927539 |
| IFIT5 | 2.74721748 | 7.27915175 | 8.43837824 | 2.56E-11 | 3.41E-09 | 15.6178986 |
| MTCYBP11 | -2.7435391 | -0.2887552 | -4.5173111 | 3.62E-05 | 5.24E-04 | 2.12087564 |
| MAFB | 2.74232006 | 6.02979127 | 8.15396362 | 7.18E-11 | 8.27E-09 | 14.5993888 |
| ANKRD22 | 2.73002213 | 4.05049279 | 4.68139284 | 2.07E-05 | 3.27E-04 | 2.45412183 |
| SAMD9 | 2.72031177 | 8.68924745 | 8.87267886 | 5.38E-12 | 8.42E-10 | 17.1608559 |
| CDC42P3 | 2.71638992 | -0.8968121 | 6.8756291 | 7.75E-09 | 4.49E-07 | 8.69629335 |
| ANO7L1 | 2.71499966 | 1.62046811 | 6.23534557 | 8.11E-08 | 3.28E-06 | 7.85024119 |
| KIAA1958 | 2.71275634 | 5.31599908 | 9.3732959 | 9.09E-13 | 1.88E-10 | 18.8958829 |
| CDKN1A | 2.71233707 | 5.43584719 | 9.77455135 | 2.23E-13 | 6.43E-11 | 20.2745214 |
| OR52B6 | 2.70766442 | -0.5613698 | 6.35299315 | 5.27E-08 | 2.27E-06 | 7.55952628 |
| PARP14 | 2.70468947 | 10.0709856 | 9.70506322 | 2.84E-13 | 7.72E-11 | 19.9918869 |
| SDC2 | -2.7031342 | 0.07627288 | -4.0727718 | 1.59E-04 | 0.00173654 | 0.83290179 |
| MTATP6P26 | 2.70134453 | 1.18368267 | 5.50676266 | 1.14E-06 | 3.05E-05 | 5.34819916 |
| TGM3 | -2.6505293 | 0.72309211 | -4.373459 | 5.88E-05 | 7.76E-04 | 1.72443659 |
| ELOVL3 | 2.63361207 | -0.5896168 | 4.27297098 | 8.22E-05 | 0.00101905 | 1.36721314 |
| AIM2 | 2.63042907 | 4.85562615 | 8.44921855 | 2.46E-11 | 3.31E-09 | 15.6649924 |
| FAM26F | 2.62147121 | 3.37183012 | 4.15422117 | 1.22E-04 | 0.00140048 | 0.79432952 |
| LIPA | 2.6206748 | 6.62085137 | 9.38237859 | 8.80E-13 | 1.86E-10 | 18.9240058 |
| SCRT2 | -2.6198771 | 2.32830976 | -5.6894458 | 5.90E-07 | 1.77E-05 | 5.96377351 |
| CTD-2521M24.13 | 2.59181724 | 1.7816248 | 8.01206774 | 1.20E-10 | 1.25E-08 | 13.9706144 |
| PARP9 | 2.58190369 | 8.81203982 | 9.28155946 | 1.26E-12 | 2.34E-10 | 18.5783678 |
| ARHGAP23 | 2.57524689 | 1.27417879 | 6.05124687 | 1.59E-07 | 5.87E-06 | 7.1972416 |
| VSIG10L | 2.5655852 | 0.59080645 | 7.35120348 | 1.35E-09 | 9.60E-08 | 11.4635812 |
| PLSCR2 | 2.56350779 | 3.25711476 | 7.61925013 | 5.05E-10 | 4.18E-08 | 12.7472121 |
| RPL12P42 | 2.55711218 | -1.2840944 | 5.22672347 | 3.09E-06 | 7.04E-05 | 3.77858398 |
| OLFM5P | 2.55679253 | 0.67130987 | 5.94063709 | 2.37E-07 | 8.17E-06 | 6.73850607 |
| ZBP1 | 2.55536937 | 7.80617961 | 9.92947819 | 1.30E-13 | 4.36E-11 | 20.801104 |
| MTCO3P43 | 2.54509931 | 0.73033391 | 5.08651868 | 5.07E-06 | 1.06E-04 | 3.94685011 |
| PXT1 | 2.53973065 | -0.139246 | 3.55140014 | 8.23E-04 | 0.00638651 | -0.6118604 |
| ENTPD2 | -2.5396507 | -1.63185 | -4.9593014 | 7.91E-06 | 1.52E-04 | 3.04018163 |
| MTCO3P5 | -2.5382002 | -0.3881875 | -4.1943615 | 1.07E-04 | 0.00125432 | 1.17584611 |
| RP11-290L7.5 | -2.5380013 | 1.32771578 | -6.804467 | 1.01E-08 | 5.56E-07 | 9.76941017 |
| RNA5SP237 | 2.53741272 | -1.8292897 | 3.64248169 | 6.22E-04 | 0.00513743 | -0.4969738 |
| HSPA8P5 | 2.53362275 | -0.1839161 | 7.94024483 | 1.56E-10 | 1.53E-08 | 12.7562943 |
| AC007899.3 | 2.52681854 | 2.60500859 | 6.98284999 | 5.22E-09 | 3.27E-07 | 10.4928059 |
| RNA5SP39 | 2.52583965 | 1.65110649 | 6.71943662 | 1.38E-08 | 7.38E-07 | 9.52427234 |
| TNFAIP6 | 2.51800536 | 5.67688404 | 6.42945899 | 3.99E-08 | 1.81E-06 | 8.39568369 |
| PNPT1 | 2.51484099 | 5.75610442 | 13.5728219 | 9.72E-19 | 3.65E-15 | 32.3301907 |
| MARCO | 2.51373144 | 1.98768601 | 4.09572312 | 1.47E-04 | 0.0016346 | 0.76618646 |
| AGRN | 2.51221256 | 4.14073762 | 7.35400988 | 1.34E-09 | 9.54E-08 | 11.7869005 |
| TRIM5 | 2.50679647 | 6.63407514 | 10.865287 | 5.34E-15 | 3.58E-12 | 23.9317989 |
| CES1 | 2.505195 | 3.95809228 | 4.3864534 | 5.63E-05 | 7.51E-04 | 1.44572485 |
| OR52T1P | 2.49967994 | -0.1468784 | 5.36439493 | 1.89E-06 | 4.64E-05 | 4.66989618 |
| PARP12 | 2.49204299 | 7.68605972 | 12.543977 | 2.29E-17 | 4.71E-14 | 29.2302759 |
| TREX1 | 2.4914651 | 5.35628404 | 10.1905401 | 5.28E-14 | 2.16E-11 | 21.6854742 |
| UBQLNL | 2.48205082 | 0.63955971 | 5.86435657 | 3.13E-07 | 1.03E-05 | 6.47750346 |
| ACO1 | 2.48100296 | 7.58904119 | 9.41452261 | 7.86E-13 | 1.70E-10 | 19.0392613 |
| LGALS3BP | 2.46850276 | 5.59434398 | 8.39727809 | 2.97E-11 | 3.85E-09 | 15.4659678 |
| INHBB | -2.4642083 | -1.3086028 | -3.9309669 | 2.51E-04 | 0.00254005 | 0.34864919 |
| TNFSF10 | 2.46036128 | 8.89749434 | 8.63421961 | 1.26E-11 | 1.86E-09 | 16.3272519 |
| RAB3C | -2.4592325 | -0.3896796 | -3.4563064 | 0.0010981 | 0.00795309 | -0.8604946 |
| BST2 | 2.45804294 | 6.26707922 | 10.8085415 | 6.46E-15 | 4.00E-12 | 23.7461128 |
| XXbac-BCX360G3.2 | 2.43992339 | -0.2522815 | 7.96619696 | 1.42E-10 | 1.41E-08 | 12.8774498 |
| XXbac-BPG554J19.2 | 2.43992339 | -0.2522815 | 7.96619696 | 1.42E-10 | 1.41E-08 | 12.8774498 |
| FAM46A | 2.42672103 | 6.48028836 | 9.65346492 | 3.40E-13 | 8.92E-11 | 19.8583391 |
| MTCO2P5 | -2.4255069 | -1.078138 | -3.9308866 | 2.51E-04 | 0.00254005 | 0.38287285 |
| CCR1 | 2.42491266 | 8.0636848 | 6.58805175 | 2.23E-08 | 1.12E-06 | 9.01432257 |
| TLR7 | 2.41417832 | 5.27772077 | 9.19401038 | 1.71E-12 | 3.04E-10 | 18.2724614 |
| RP11-365F18.6 | 2.40900946 | -1.6449969 | 4.40024274 | 5.38E-05 | 7.23E-04 | 1.45994587 |
| IL4I1 | 2.40438228 | 1.9925996 | 6.46665385 | 3.48E-08 | 1.61E-06 | 8.67792137 |
| CLC | -2.4004144 | 4.68286129 | -4.7088091 | 1.88E-05 | 3.05E-04 | 2.4683092 |
| MTND5P24 | -2.3929536 | -0.0506884 | -4.021816 | 1.87E-04 | 0.00199214 | 0.68441877 |
| KLHDC7B | 2.39039005 | 3.61523963 | 8.39445279 | 3.00E-11 | 3.86E-09 | 15.4817221 |
| AC074338.4 | 2.39011776 | 2.56108997 | 7.27351925 | 1.79E-09 | 1.23E-07 | 11.5202369 |
| RP11-435F17.3 | -2.3852795 | 0.03481579 | -4.1047281 | 1.43E-04 | 0.00159862 | 0.91499581 |
| DDX60L | 2.38508459 | 9.19382912 | 8.63182527 | 1.28E-11 | 1.86E-09 | 16.3184119 |
| STAT1 | 2.36897278 | 8.44139426 | 9.88334843 | 1.53E-13 | 4.86E-11 | 20.6397456 |
| RP1-145M24.1 | 2.36099204 | -0.9120701 | 4.85011221 | 1.16E-05 | 2.08E-04 | 3.0694691 |
| PGAP1 | 2.36052524 | 4.87105809 | 9.53994006 | 5.06E-13 | 1.16E-10 | 19.4713423 |
| TSPEAR | -2.3534662 | -0.086084 | -4.2000053 | 1.05E-04 | 0.00123432 | 1.19578115 |
| CDK2AP2P2 | 2.33466987 | -1.6310867 | 3.68715218 | 5.42E-04 | 0.00461137 | -0.3487318 |
| SMIM10L2A | -2.3308097 | 0.03798606 | -6.2023412 | 9.15E-08 | 3.62E-06 | 7.31151641 |
| RP1-102G20.2 | 2.33058248 | -0.4298822 | 5.47569157 | 1.27E-06 | 3.34E-05 | 4.97556657 |
| RP11-820K3.2 | 2.33038524 | 1.85978069 | 7.2538919 | 1.93E-09 | 1.32E-07 | 11.3779377 |
| FBXO6 | 2.33003758 | 5.06499516 | 8.10584431 | 8.55E-11 | 9.50E-09 | 14.4433106 |
| CA4 | -2.3218886 | 2.62805651 | -5.3141249 | 2.26E-06 | 5.39E-05 | 4.67973096 |
| CD300E | 2.32110372 | 6.9186064 | 7.65799223 | 4.38E-10 | 3.69E-08 | 12.8277159 |
| ANGPT1 | -2.3201929 | 1.45513506 | -6.1034455 | 1.31E-07 | 4.96E-06 | 7.39640418 |
| TRIM22 | 2.31910964 | 9.56848714 | 10.6783008 | 1.00E-14 | 5.71E-12 | 23.2434209 |
| RP11-686D22.7 | 2.31218552 | 2.82891699 | 8.98430396 | 3.61E-12 | 6.11E-10 | 17.4937381 |
| C3AR1 | 2.3118645 | 5.33853286 | 5.43918709 | 1.45E-06 | 3.70E-05 | 4.88373536 |
| PRLR | 2.30973537 | 2.8164814 | 6.29770649 | 6.46E-08 | 2.71E-06 | 8.07659825 |
| CNTNAP3B | -2.3046638 | 3.82455668 | -7.0728091 | 3.75E-09 | 2.41E-07 | 10.7753977 |
| PML | 2.30216804 | 8.28776 | 10.4411764 | 2.24E-14 | 1.08E-11 | 22.5125183 |
| MTND3P10 | 2.29921729 | -0.5041549 | 4.11336146 | 1.39E-04 | 0.00155883 | 0.89931244 |
| GCH1 | 2.27776594 | 6.13728168 | 8.51006556 | 1.98E-11 | 2.71E-09 | 15.8607395 |
| TOR1B | 2.27534217 | 6.46806573 | 8.32241686 | 3.90E-11 | 4.85E-09 | 15.1946875 |
| TMEM123 | 2.27506727 | 9.06342047 | 9.55950096 | 4.72E-13 | 1.12E-10 | 19.5260845 |
| CEACAM1 | 2.27271235 | 6.21907874 | 7.42938492 | 1.01E-09 | 7.58E-08 | 11.9900333 |
| AXL | 2.26668996 | 2.90081738 | 6.41467508 | 4.21E-08 | 1.89E-06 | 8.47589606 |
| RAB36 | -2.2552527 | 2.87329365 | -5.5710889 | 9.03E-07 | 2.54E-05 | 5.51401894 |
| TDRD7 | 2.25219773 | 6.41302389 | 9.90492699 | 1.42E-13 | 4.66E-11 | 20.7178397 |
| CMKLR1 | 2.25004122 | 5.04936467 | 8.19714098 | 6.14E-11 | 7.21E-09 | 14.7554049 |
| DNMT3L | -2.2474545 | -0.3699328 | -3.7883304 | 3.95E-04 | 0.00360262 | 0.01989202 |
| MTND4P26 | 2.23325629 | 1.35267628 | 5.38469755 | 1.76E-06 | 4.35E-05 | 4.96241907 |
| GRAMD1B | 2.22731701 | 5.02339407 | 7.283271 | 1.73E-09 | 1.19E-07 | 11.4836299 |
| TTC21A | 2.22483347 | 4.70904026 | 7.86101196 | 2.09E-10 | 1.90E-08 | 13.5637205 |
| PPP1R2P1 | 2.21598114 | -0.3497927 | 5.98448648 | 2.02E-07 | 7.16E-06 | 6.4937234 |
| CLDN18 | 2.21467427 | 1.57463788 | 6.8715449 | 7.86E-09 | 4.54E-07 | 10.0747122 |
| RNASE2 | 2.21143269 | 4.45181758 | 5.0438121 | 5.89E-06 | 1.19E-04 | 3.57808047 |
| IDO1 | 2.21133109 | 3.66040789 | 3.7874576 | 3.96E-04 | 0.00360826 | -0.3613712 |
| STAT2 | 2.20837887 | 8.13532584 | 8.55982592 | 1.65E-11 | 2.32E-09 | 16.0580795 |
| CD274 | 2.20601853 | 5.19443924 | 5.78652407 | 4.15E-07 | 1.31E-05 | 6.12412097 |
| CXCL6 | -2.1956761 | -1.4948266 | -3.8852196 | 2.90E-04 | 0.00285588 | 0.20009032 |
| CD1C | -2.1949283 | 1.87759811 | -5.8614909 | 3.17E-07 | 1.04E-05 | 6.57457639 |
| CASP5 | 2.19172841 | 4.01877905 | 5.2734773 | 2.62E-06 | 6.10E-05 | 4.37093047 |
| FAM101B | -2.1871057 | 5.3505249 | -8.8369863 | 6.11E-12 | 9.48E-10 | 17.0232759 |
| PTPRO | 2.18670716 | 3.18523138 | 6.58761974 | 2.23E-08 | 1.12E-06 | 9.07219086 |
| OR52H2P | 2.18438151 | -0.3869706 | 3.94266631 | 2.42E-04 | 0.0024632 | 0.44774705 |
| RP1-102G20.4 | 2.17603342 | -0.3993263 | 3.75457881 | 4.39E-04 | 0.00389965 | -0.068019 |
| SUCNR1 | 2.16780629 | 1.10120604 | 4.84450405 | 1.18E-05 | 2.11E-04 | 3.19559735 |
| TNFSF13B | 2.16563076 | 7.47993863 | 6.82211841 | 9.43E-09 | 5.35E-07 | 9.82793076 |
| LGALS9 | 2.16263851 | 7.9521542 | 9.39787436 | 8.33E-13 | 1.78E-10 | 18.9827128 |
| JPH4 | 2.15920378 | 1.01365576 | 4.49091899 | 3.96E-05 | 5.65E-04 | 2.07255707 |
| DUSP5 | 2.15002857 | 4.71135078 | 10.0344152 | 9.04E-14 | 3.46E-11 | 21.1534595 |
| RP11-686D22.3 | 2.14860068 | 2.31980058 | 7.89675841 | 1.83E-10 | 1.72E-08 | 13.6939837 |
| LILRB4 | 2.1482113 | 5.10794998 | 7.3926182 | 1.16E-09 | 8.50E-08 | 11.8702005 |
| ADAMTS7P1 | -2.1456806 | 1.15230327 | -3.4570535 | 0.00109563 | 0.00793828 | -0.9696876 |
| NRXN1 | -2.1455433 | 0.60187682 | -3.5962568 | 7.17E-04 | 0.00571786 | -0.6024387 |
| SDHCP2 | 2.14413963 | -1.5847047 | 3.50285858 | 9.54E-04 | 0.00716625 | -0.794646 |
| WNK2 | -2.1386804 | -0.0872533 | -4.1936195 | 1.07E-04 | 0.00125661 | 1.15734615 |
| TRIM74 | -2.1361965 | -0.0775645 | -3.4151599 | 0.00124253 | 0.00872654 | -0.9701319 |
| PI4K2B | 2.13185467 | 4.83065353 | 12.083675 | 9.87E-17 | 1.43E-13 | 27.8102286 |
| MMP28 | -2.1312933 | 0.17960488 | -5.5837444 | 8.63E-07 | 2.44E-05 | 5.4557004 |
| PRM3 | 2.13116346 | -1.3179885 | 5.61221265 | 7.79E-07 | 2.25E-05 | 4.94298385 |
| ZCCHC2 | 2.12038707 | 8.29266346 | 8.88644007 | 5.12E-12 | 8.22E-10 | 17.2072487 |
| SDC3 | 2.1165728 | 3.88583406 | 6.12889019 | 1.20E-07 | 4.59E-06 | 7.40433754 |
| BLVRA | 2.11394641 | 4.55221884 | 8.51096175 | 1.97E-11 | 2.71E-09 | 15.8848635 |
| ZNF684 | 2.1131102 | 2.82691188 | 7.479204 | 8.44E-10 | 6.60E-08 | 12.2511719 |
| P2RY6 | 2.11174621 | 2.66739987 | 7.97343011 | 1.38E-10 | 1.40E-08 | 13.9876466 |
| HELZ2 | 2.10606631 | 8.42750677 | 7.86347451 | 2.07E-10 | 1.89E-08 | 13.5918847 |
| MTND6P11 | -2.1046201 | -1.9226973 | -3.4057096 | 0.00127816 | 0.00894001 | -1.0368706 |
| PDZD3 | -2.0912464 | -0.4477927 | -3.5764017 | 7.62E-04 | 0.00600817 | -0.5455794 |
| KAT2A | 2.08933556 | 5.13830469 | 10.0224255 | 9.42E-14 | 3.54E-11 | 21.1179151 |
| FABP6 | -2.0863055 | -0.7169177 | -3.832899 | 3.43E-04 | 0.00322871 | 0.14889829 |
| AC003080.4 | 2.08607492 | 0.734552 | 5.21294501 | 3.24E-06 | 7.36E-05 | 4.38639736 |
| RP11-24B13.2 | -2.0832369 | 0.60925809 | -5.9701126 | 2.13E-07 | 7.45E-06 | 6.88642294 |
| PTP4A1 | 2.07608125 | 6.74575131 | 8.76788812 | 7.83E-12 | 1.19E-09 | 16.7757356 |
| AC007919.18 | 2.06707413 | 1.27230626 | 5.35699721 | 1.94E-06 | 4.73E-05 | 4.8710709 |
| NRCAM | -2.0590791 | 0.62861666 | -3.5030592 | 9.53E-04 | 0.00716476 | -0.749001 |
| TTC26 | 2.05855 | 3.39918635 | 6.52134603 | 2.85E-08 | 1.38E-06 | 8.82245031 |
| NOG | -2.0580758 | 1.55823635 | -4.8184655 | 1.29E-05 | 2.27E-04 | 3.10341202 |
| GPBAR1 | 2.05360718 | 5.14607517 | 6.89413611 | 7.24E-09 | 4.25E-07 | 10.0685662 |
| ADPRH | 2.04949449 | 4.19635974 | 7.89967041 | 1.81E-10 | 1.71E-08 | 13.7196622 |
| DOCK4 | 2.04883796 | 5.74370591 | 5.8147681 | 3.75E-07 | 1.21E-05 | 6.18651871 |
| CNTNAP3 | -2.0359098 | 4.40483926 | -7.5099475 | 7.54E-10 | 5.97E-08 | 12.3099189 |
| MERTK | 2.0325357 | 2.69087111 | 4.72693941 | 1.77E-05 | 2.90E-04 | 2.65180052 |
| LGALS9B | 2.02779636 | 2.127297 | 6.89670979 | 7.17E-09 | 4.22E-07 | 10.1845376 |
| FNDC11 | 2.02667146 | 2.05500811 | 6.29631492 | 6.49E-08 | 2.71E-06 | 8.08279277 |
| APOL6 | 2.02300822 | 9.21812922 | 10.2409279 | 4.44E-14 | 1.94E-11 | 21.8239913 |
| RNF213 | 2.01460871 | 11.2863453 | 8.24202493 | 5.22E-11 | 6.32E-09 | 14.9259141 |
| PDCD1LG2 | 2.01412303 | 1.71111776 | 4.2068123 | 1.02E-04 | 0.0012115 | 1.16861307 |
| GPD2 | 2.00963844 | 5.64714105 | 7.73880925 | 3.26E-10 | 2.84E-08 | 13.1018043 |
| NTNG2 | 2.00764609 | 7.72401466 | 6.2948338 | 6.53E-08 | 2.72E-06 | 7.94197752 |
| EPB41L5 | 2.00461729 | 4.41131646 | 6.20459585 | 9.08E-08 | 3.60E-06 | 7.63924529 |
| RRAS | 2.00433232 | 3.91639112 | 9.9462753 | 1.23E-13 | 4.26E-11 | 20.847727 |

**Supplemental Table 5.** Differentially expressed genes day 10 post infection

| **geneID** | **logFC** | **AveExpr** | **t** | **P.Value** | **adj.P.Val** | **B** |
| --- | --- | --- | --- | --- | --- | --- |
| IFI27 | 7.82755146 | 6.52857751 | 9.16414958 | 1.90E-12 | 1.49E-09 | 17.9274876 |
| CXCL10 | 5.67562505 | 3.2144798 | 4.72900825 | 1.76E-05 | 4.56E-04 | 2.81068971 |
| ADAMTS5 | -5.3467268 | -1.5536131 | -6.8241722 | 9.36E-09 | 1.05E-06 | 5.98542247 |
| SIGLEC1 | 5.17895214 | 6.35550789 | 8.34932019 | 3.54E-11 | 1.27E-08 | 15.2924464 |
| CH17-296N19.1 | -5.1760918 | 2.01758359 | -6.819802 | 9.51E-09 | 1.06E-06 | 9.37990833 |
| IFI44L | 5.05287834 | 9.26563843 | 9.53409407 | 5.16E-13 | 5.70E-10 | 19.3581719 |
| PI3 | -4.9472024 | 2.47364729 | -4.7379769 | 1.70E-05 | 4.45E-04 | 2.83667906 |
| KCTD14 | 4.91432621 | 0.65016798 | 5.80237477 | 3.92E-07 | 2.10E-05 | 5.55391521 |
| CXCL11 | 4.85336527 | -1.12088 | 3.69774734 | 5.24E-04 | 0.00644501 | -0.8478936 |
| USP18 | 4.80613434 | 5.2942684 | 10.1602086 | 5.86E-14 | 1.50E-10 | 21.3148885 |
| RSAD2 | 4.68563761 | 8.78506662 | 8.25694485 | 4.94E-11 | 1.66E-08 | 14.9913015 |
| RBM17P1 | -4.6327491 | -0.2524074 | -5.7715444 | 4.38E-07 | 2.26E-05 | 5.01618703 |
| RBM17P2 | -4.4741296 | -2.1246294 | -6.4309124 | 3.96E-08 | 3.32E-06 | 4.95425831 |
| SERPING1 | 4.44961466 | 5.97008766 | 8.22958639 | 5.46E-11 | 1.77E-08 | 14.8925653 |
| MTND5P24 | -4.3418296 | -0.0506884 | -4.9217618 | 9.02E-06 | 2.63E-04 | 2.74849831 |
| BATF2 | 4.32773462 | 5.07853875 | 8.19916875 | 6.09E-11 | 1.91E-08 | 14.7685016 |
| GBP1P1 | 4.3238933 | 3.00256366 | 5.97503421 | 2.10E-07 | 1.26E-05 | 6.94506869 |
| PHF24 | -4.3021965 | -1.2092825 | -4.2098252 | 1.01E-04 | 0.00185063 | 0.60758845 |
| MTND6P11 | -4.105259 | -1.9226973 | -4.7754459 | 1.50E-05 | 4.02E-04 | 1.22518579 |
| NAT8 | 4.09390253 | 1.16331174 | 3.64875031 | 6.10E-04 | 0.00726581 | -0.3557456 |
| MTND4P22 | -4.0843374 | -0.4789305 | -5.2160827 | 3.21E-06 | 1.15E-04 | 3.47517481 |
| HESX1 | 4.0646517 | -1.1411462 | 3.54228978 | 8.46E-04 | 0.00930935 | -1.0912149 |
| MTCO2P5 | -4.041057 | -1.078138 | -4.8851722 | 1.02E-05 | 2.94E-04 | 2.1552439 |
| ETV7 | 4.03833038 | 4.46985866 | 6.83821107 | 8.89E-09 | 1.01E-06 | 9.98306563 |
| LIF | 4.02088819 | 0.1467629 | 5.41888923 | 1.56E-06 | 6.43E-05 | 4.40961858 |
| MTCYBP11 | -3.9832065 | -0.2887552 | -5.1949951 | 3.46E-06 | 1.22E-04 | 3.52183653 |
| IFI44 | 3.97403104 | 7.7459889 | 8.85776556 | 5.67E-12 | 3.23E-09 | 17.0957617 |
| CMPK2 | 3.9727182 | 7.47598457 | 8.69775044 | 1.01E-11 | 4.51E-09 | 16.5408549 |
| USP41 | 3.93390557 | 1.09222199 | 5.48292893 | 1.24E-06 | 5.36E-05 | 4.6802576 |
| SDC2 | -3.9293003 | 0.07627288 | -4.6563381 | 2.26E-05 | 5.61E-04 | 2.16978773 |
| ATF3 | 3.90570407 | 2.72288822 | 5.4769204 | 1.27E-06 | 5.45E-05 | 5.26598464 |
| MTCO3P5 | -3.8604916 | -0.3881875 | -4.93563 | 8.59E-06 | 2.53E-04 | 2.79009311 |
| IFITM3 | 3.85512711 | 8.83547974 | 8.69723202 | 1.01E-11 | 4.51E-09 | 16.5228388 |
| OAS3 | 3.85419081 | 9.74421226 | 8.15611883 | 7.12E-11 | 2.19E-08 | 14.6245842 |
| IFIT3 | 3.80923933 | 9.34813991 | 7.79437331 | 2.66E-10 | 6.41E-08 | 13.3613841 |
| RMI2 | 3.80154708 | 3.20860803 | 7.34476694 | 1.38E-09 | 2.36E-07 | 11.6696546 |
| TMPRSS2 | 3.7786269 | -1.0412947 | 4.27422877 | 8.19E-05 | 0.00157524 | 0.46023908 |
| SPATS2L | 3.77234662 | 6.06216049 | 8.77683188 | 7.58E-12 | 3.75E-09 | 16.8082715 |
| IFIT1 | 3.7593796 | 8.99783691 | 7.22135086 | 2.17E-09 | 3.31E-07 | 11.3361329 |
| C1QB | 3.74907073 | 1.39849469 | 3.83870378 | 3.37E-04 | 0.00461673 | 0.1593435 |
| HNRNPCP4 | 3.73898936 | -2.0770287 | 4.7993622 | 1.38E-05 | 3.74E-04 | 1.75943058 |
| NEURL3 | 3.72660277 | -0.1059035 | 5.82489377 | 3.61E-07 | 1.96E-05 | 5.42298994 |
| ALMS1P1 | 3.71602586 | 1.07374967 | 3.58180417 | 7.50E-04 | 0.00850558 | -0.5313488 |
| OAS1 | 3.71178263 | 8.49494286 | 9.44543725 | 7.05E-13 | 7.35E-10 | 19.1021597 |
| LY6E | 3.66107237 | 9.3559693 | 9.04336061 | 2.93E-12 | 2.11E-09 | 17.6970965 |
| MTND3P9 | -3.6576089 | -2.087085 | -4.0948653 | 1.48E-04 | 0.00247854 | -0.0873206 |
| NMRAL1P1 | 3.62533144 | -0.3926125 | 3.87599997 | 2.99E-04 | 0.00419605 | 0.09709126 |
| MTND4LP12 | -3.5868001 | -1.973906 | -4.0998409 | 1.45E-04 | 0.00245842 | 0.09003632 |
| IFI6 | 3.50088246 | 8.08614509 | 8.63599476 | 1.26E-11 | 5.13E-09 | 16.327262 |
| EPSTI1 | 3.49919446 | 7.62773183 | 10.5326098 | 1.64E-14 | 7.71E-11 | 22.7507206 |
| RTP4 | 3.49009339 | 5.4111897 | 10.3692963 | 2.86E-14 | 1.08E-10 | 22.1664796 |
| FBXO39 | 3.47415088 | 1.93155647 | 7.41765018 | 1.06E-09 | 1.87E-07 | 11.5250411 |
| OVOL1 | 3.46574079 | -2.2541658 | 4.94390089 | 8.35E-06 | 2.48E-04 | 1.91156403 |
| CDH2 | -3.4592473 | -1.7842818 | -3.9068092 | 2.71E-04 | 0.00391763 | -0.2256706 |
| TSPEAR | -3.4446136 | -0.086084 | -5.1093288 | 4.67E-06 | 1.53E-04 | 3.45423827 |
| METTL7B | 3.44438497 | -1.417302 | 4.36094768 | 6.13E-05 | 0.00125265 | 1.04013601 |
| RP4-641G12.3 | 3.38466192 | 1.07856285 | 6.28471932 | 6.77E-08 | 5.11E-06 | 7.29447571 |
| OASL | 3.37235012 | 7.54938765 | 8.89133878 | 5.03E-12 | 3.15E-09 | 17.2178614 |
| HEY1 | -3.3653886 | -1.9718792 | -4.5612145 | 3.12E-05 | 7.22E-04 | 1.2583561 |
| ANKRD22 | 3.33639642 | 4.05049279 | 5.78915474 | 4.11E-07 | 2.17E-05 | 6.300012 |
| EXOC3L1 | 3.33531336 | 2.25582378 | 5.50866606 | 1.13E-06 | 5.03E-05 | 5.28737412 |
| SDC1 | 3.33442827 | -1.0985613 | 3.66799746 | 5.75E-04 | 0.00691982 | -0.5426037 |
| CDC45 | 3.32116138 | -0.0135068 | 6.43569591 | 3.90E-08 | 3.31E-06 | 7.49834626 |
| PTPRU | 3.28434792 | -0.3957957 | 3.95184229 | 2.35E-04 | 0.00349926 | 0.23589927 |
| TEKT1 | 3.28337214 | -1.6906615 | 3.86730931 | 3.07E-04 | 0.00428803 | -0.4028373 |
| ZDHHC4P1 | 3.2796598 | 3.94448499 | 7.50870755 | 7.57E-10 | 1.51E-07 | 12.3013506 |
| ISG15 | 3.2749732 | 0.3381444 | 3.87736169 | 2.98E-04 | 0.00418102 | 0.10963189 |
| MT2A | 3.24947131 | 5.47736325 | 9.33305345 | 1.05E-12 | 9.84E-10 | 18.7157585 |
| RP11-290L7.3 | -3.2389886 | -3.3051161 | -3.6817997 | 5.51E-04 | 0.00669391 | -1.0300679 |
| SMTNL1 | 3.2348584 | 5.65340479 | 8.96307562 | 3.89E-12 | 2.61E-09 | 17.4599039 |
| CARD17 | 3.22574977 | 0.50468438 | 4.85818322 | 1.13E-05 | 3.18E-04 | 3.01766805 |
| IGLV1-47 | 3.20577621 | 3.55912851 | 5.25741401 | 2.77E-06 | 1.02E-04 | 4.48623177 |
| GBP1 | 3.17944675 | 8.28332624 | 9.928965 | 1.30E-13 | 2.72E-10 | 20.724647 |
| RRM2 | 3.16930129 | 3.3856448 | 8.30266315 | 4.19E-11 | 1.46E-08 | 15.0862634 |
| FABP6 | -3.1602345 | -0.7169177 | -4.8137269 | 1.31E-05 | 3.62E-04 | 2.54792832 |
| SCRT2 | -3.1374844 | 2.32830976 | -6.3575409 | 5.19E-08 | 4.13E-06 | 8.26242655 |
| NRXN1 | -3.1321552 | 0.60187682 | -4.5972001 | 2.76E-05 | 6.64E-04 | 2.38432277 |
| IFIT2 | 3.13128681 | 9.66057711 | 7.10356551 | 3.35E-09 | 4.77E-07 | 10.9211999 |
| HES4 | 3.11515474 | 2.18929656 | 4.16807704 | 1.16E-04 | 0.00205328 | 1.11263345 |
| OAS2 | 3.11257688 | 9.04882392 | 9.17869722 | 1.81E-12 | 1.48E-09 | 18.1756098 |
| SOCS1 | 3.1008503 | 3.48053286 | 6.56384217 | 2.43E-08 | 2.24E-06 | 9.01842006 |
| LIPM | 3.08533865 | -0.8572704 | 3.80048357 | 3.80E-04 | 0.00509799 | -0.1354597 |
| IGLC1 | 3.07919353 | 5.14650159 | 6.42314821 | 4.08E-08 | 3.39E-06 | 8.48881887 |
| IGLV4-69 | 3.07581126 | 1.84852262 | 3.75018458 | 4.45E-04 | 0.00575933 | -0.125174 |
| ENTPD2 | -3.0622576 | -1.63185 | -5.5939495 | 8.32E-07 | 3.88E-05 | 4.06233169 |
| RUFY4 | 3.05105461 | 2.76315936 | 6.94747879 | 5.95E-09 | 7.30E-07 | 10.3051023 |
| PBK | 3.04831124 | -1.0518386 | 4.87271468 | 1.07E-05 | 3.05E-04 | 2.51247284 |
| PXT1 | 3.03168802 | -0.139246 | 4.36829668 | 5.98E-05 | 0.00122967 | 1.54910467 |
| IGLV1-44 | 3.02926826 | 3.2475498 | 5.70994546 | 5.48E-07 | 2.76E-05 | 6.05078216 |
| DDX60 | 3.00828577 | 7.68361699 | 9.69019425 | 2.99E-13 | 4.68E-10 | 19.9538689 |
| ALPL | -2.9865341 | 5.98271276 | -5.2503776 | 2.84E-06 | 1.04E-04 | 4.3585014 |
| HIST1H3G | 2.94304087 | 3.67375764 | 8.11117581 | 8.39E-11 | 2.50E-08 | 14.4389439 |
| RSPH14 | -2.9420509 | -0.9330718 | -5.1532944 | 4.00E-06 | 1.36E-04 | 3.53057494 |
| HERC5 | 2.92032913 | 7.68061448 | 7.01216618 | 4.69E-09 | 6.03E-07 | 10.5741233 |
| OTX1 | -2.9173386 | 0.45074125 | -5.4726279 | 1.29E-06 | 5.53E-05 | 5.06578971 |
| CCRL2 | 2.9053168 | 3.41405055 | 7.14582638 | 2.87E-09 | 4.21E-07 | 11.0630251 |
| PNMA6A | -2.8877657 | -1.2319706 | -5.5567655 | 9.51E-07 | 4.35E-05 | 4.52188222 |
| POLR2CP1 | 2.88420876 | -1.5447288 | 3.57915174 | 7.56E-04 | 0.00856291 | -1.0620246 |
| UCHL1 | 2.88169661 | -3.0329491 | 4.18331246 | 1.11E-04 | 0.00198263 | -0.2385459 |
| CACNA2D3 | -2.8644468 | 1.3775838 | -7.1325601 | 3.01E-09 | 4.32E-07 | 10.4589132 |
| IGLV2-23 | 2.86260049 | 2.85540372 | 4.49532682 | 3.90E-05 | 8.68E-04 | 2.03823524 |
| LAP3 | 2.86031047 | 7.05791179 | 12.4354869 | 3.23E-17 | 3.03E-13 | 28.7664209 |
| JCHAIN | 2.86027032 | 6.99845032 | 6.24104481 | 7.94E-08 | 5.80E-06 | 7.82478562 |
| NECAB2 | -2.8402407 | 1.14246833 | -5.8269223 | 3.59E-07 | 1.96E-05 | 6.32024304 |
| LAMP3 | 2.82622079 | 4.21810844 | 7.05411215 | 4.02E-09 | 5.35E-07 | 10.7443229 |
| IGLC7 | 2.81056044 | -1.8091036 | 3.57501664 | 7.66E-04 | 0.00865124 | -0.955853 |
| INHBB | -2.8031057 | -1.3086028 | -4.3117849 | 7.23E-05 | 0.00142996 | 1.12804423 |
| SUCNR1 | 2.78843238 | 1.10120604 | 6.46350634 | 3.52E-08 | 3.06E-06 | 8.44564116 |
| IRF7 | 2.77607172 | 7.85958316 | 9.3858052 | 8.70E-13 | 8.60E-10 | 18.9119499 |
| MX1 | 2.76545477 | 9.41242592 | 8.04349824 | 1.07E-10 | 3.10E-08 | 14.2388877 |
| HIST1H3C | 2.75571121 | 2.48037715 | 5.06582527 | 5.45E-06 | 1.74E-04 | 3.90789052 |
| FRMD3 | 2.70087281 | 4.62157375 | 6.49260854 | 3.16E-08 | 2.81E-06 | 8.74735963 |
| MDK | 2.68933537 | 1.48244713 | 5.56782338 | 9.14E-07 | 4.22E-05 | 5.46951769 |
| CDCA5 | 2.68630657 | 0.87812563 | 6.63056465 | 1.91E-08 | 1.85E-06 | 8.82981287 |
| TCN2 | 2.68193588 | 3.439782 | 5.48442144 | 1.23E-06 | 5.35E-05 | 5.28005065 |
| HIST1H2BM | 2.68073323 | 2.04863383 | 6.46328407 | 3.52E-08 | 3.06E-06 | 8.57076633 |
| TIMM10 | 2.67541582 | 4.01964348 | 7.48726387 | 8.19E-10 | 1.59E-07 | 12.2763156 |
| RP5-892F13.2 | -2.6684554 | -1.6083111 | -4.5831179 | 2.90E-05 | 6.85E-04 | 1.63941852 |
| SCO2 | 2.66730345 | 4.07252217 | 6.99231177 | 5.04E-09 | 6.44E-07 | 10.5278422 |
| GBP4 | 2.66424759 | 7.89037438 | 8.67951496 | 1.07E-11 | 4.59E-09 | 16.4791799 |
| IGKV2-28 | 2.65330334 | 2.38090744 | 3.86936236 | 3.05E-04 | 0.00426965 | 0.17193089 |
| IGLC2 | 2.64224492 | 6.58554216 | 5.91533546 | 2.60E-07 | 1.52E-05 | 6.6650905 |
| HIST1H3J | 2.62678032 | 3.03059472 | 6.75512365 | 1.21E-08 | 1.29E-06 | 9.67595845 |
| IGLV3-1 | 2.62016562 | 3.64601343 | 5.07506382 | 5.27E-06 | 1.70E-04 | 3.8885532 |
| HIST1H2AJ | 2.60584932 | 3.43481868 | 6.58227525 | 2.28E-08 | 2.16E-06 | 9.083145 |
| ANGPT1 | -2.594438 | 1.45513506 | -6.7195253 | 1.37E-08 | 1.42E-06 | 9.35299981 |
| RP11-24B13.2 | -2.5937182 | 0.60925809 | -6.9260929 | 6.43E-09 | 7.70E-07 | 9.70844987 |
| IGLC3 | 2.58694635 | 5.19149196 | 4.42375939 | 4.97E-05 | 0.00106047 | 1.62493972 |
| IGLV6-57 | 2.56931034 | 1.76606675 | 4.39453548 | 5.48E-05 | 0.0011391 | 1.78920129 |
| IGKV1D-39 | 2.56790257 | 3.86533446 | 6.19017802 | 9.57E-08 | 6.75E-06 | 7.69978974 |
| BHLHA15 | 2.56714297 | -0.6193916 | 3.67072435 | 5.70E-04 | 0.00687033 | -0.4099031 |
| HIST1H2BB | 2.55529958 | 0.15142088 | 4.8066563 | 1.35E-05 | 3.68E-04 | 2.84204441 |
| XAF1 | 2.55511453 | 8.87754195 | 9.53665292 | 5.12E-13 | 5.70E-10 | 19.3951219 |
| IGKV3-20 | 2.55131465 | 4.84230282 | 4.8743675 | 1.06E-05 | 3.04E-04 | 3.11977484 |
| GBP5 | 2.54374699 | 9.02544093 | 7.40040557 | 1.13E-09 | 1.98E-07 | 11.9712547 |
| C1QA | 2.53949997 | 2.17143186 | 3.5760403 | 7.63E-04 | 0.00863454 | -0.6130083 |
| IGLV2-8 | 2.52880843 | 2.72865308 | 4.6080929 | 2.66E-05 | 6.43E-04 | 2.39672989 |
| TGM3 | -2.51978 | 0.72309211 | -4.4340702 | 4.80E-05 | 0.00103351 | 1.90625477 |
| MZB1 | 2.51815896 | 4.32432781 | 6.30848134 | 6.21E-08 | 4.76E-06 | 8.09771915 |
| IFI35 | 2.51539188 | 6.29566426 | 10.2889039 | 3.77E-14 | 1.18E-10 | 21.9735183 |
| FBXO6 | 2.51166893 | 5.06499516 | 8.72304833 | 9.19E-12 | 4.32E-09 | 16.6311666 |
| KIFC1 | 2.5093442 | 1.43844572 | 6.32771808 | 5.79E-08 | 4.49E-06 | 8.0257595 |
| IGHV4-59 | 2.46801226 | 2.73953349 | 3.73932167 | 4.60E-04 | 0.00588792 | -0.2548258 |
| RP11-290L7.5 | -2.4657928 | 1.32771578 | -6.9315021 | 6.31E-09 | 7.59E-07 | 10.0728072 |
| PLSCR1 | 2.44792449 | 7.7488562 | 7.25581665 | 1.92E-09 | 3.05E-07 | 11.446721 |
| HERC6 | 2.44388621 | 6.78868489 | 12.4368508 | 3.21E-17 | 3.03E-13 | 28.7926244 |
| HIST1H2AB | 2.4433665 | 1.9982149 | 5.3953205 | 1.70E-06 | 6.89E-05 | 4.99288173 |
| CNTNAP3 | -2.4401526 | 4.40483926 | -8.7855107 | 7.35E-12 | 3.75E-09 | 16.8431831 |
| IGHV4-39 | 2.43258261 | 2.64502212 | 3.644394 | 6.18E-04 | 0.00734575 | -0.5314383 |
| CD274 | 2.43126692 | 5.19443924 | 6.35537766 | 5.23E-08 | 4.14E-06 | 8.23576817 |
| UBE2L6 | 2.42603773 | 8.04563717 | 10.5969511 | 1.32E-14 | 7.71E-11 | 22.9546091 |
| RAB3C | -2.4219061 | -0.3896796 | -3.5546526 | 8.15E-04 | 0.00904258 | -0.6168372 |
| IGKV2-30 | 2.41783345 | 1.8804678 | 4.32043615 | 7.02E-05 | 0.00140564 | 1.55077924 |
| IGLV3-21 | 2.4175827 | 3.35661179 | 4.1280579 | 1.33E-04 | 0.00227748 | 0.86444088 |
| HIST1H2AL | 2.41498518 | 2.96639264 | 7.25910733 | 1.89E-09 | 3.04E-07 | 11.4361414 |
| CNTNAP3B | -2.4089166 | 3.82455668 | -7.4303064 | 1.01E-09 | 1.86E-07 | 12.0778483 |
| IGKC | 2.40630375 | 8.62410501 | 5.86163286 | 3.16E-07 | 1.78E-05 | 6.5299302 |
| HIST1H2BO | 2.39174759 | 3.59352657 | 7.61416946 | 5.15E-10 | 1.10E-07 | 12.7117234 |
| HIST1H2AH | 2.39170301 | 3.07116528 | 6.5664857 | 2.41E-08 | 2.24E-06 | 9.02403479 |
| PDCD1LG2 | 2.38705431 | 1.71111776 | 5.12953691 | 4.35E-06 | 1.46E-04 | 4.11288345 |
| ARHGEF40 | -2.3870417 | 6.31367677 | -7.8360487 | 2.29E-10 | 5.58E-08 | 13.507481 |
| SAMD9L | 2.37972669 | 9.00100623 | 7.90404512 | 1.78E-10 | 4.41E-08 | 13.7524449 |
| BCAN | 2.37570593 | -1.2353288 | 3.70552555 | 5.11E-04 | 0.006328 | -0.425107 |
| IGHV5-51 | 2.369955 | 2.66250532 | 3.73290163 | 4.70E-04 | 0.00595985 | -0.2717251 |
| RPL37P6 | 2.36934244 | 1.39504354 | 5.8134693 | 3.77E-07 | 2.04E-05 | 6.20341234 |
| FCGR1A | 2.36877986 | 6.04678642 | 5.3753668 | 1.82E-06 | 7.34E-05 | 4.77423809 |
| EIF2AK2 | 2.36669673 | 8.74048854 | 7.23076237 | 2.10E-09 | 3.26E-07 | 11.3679352 |
| DHX58 | 2.36427928 | 6.0760597 | 8.27142845 | 4.69E-11 | 1.60E-08 | 15.0519015 |
| TAS2R40 | -2.3594755 | 0.76525918 | -4.1731109 | 1.14E-04 | 0.00202915 | 1.12539326 |
| THBD | -2.3545434 | 4.24872703 | -8.0039893 | 1.24E-10 | 3.42E-08 | 14.1060953 |
| RP11-477J21.2 | 2.35319215 | 1.66179467 | 4.80317919 | 1.36E-05 | 3.71E-04 | 3.04750802 |
| HIST1H3B | 2.35064856 | 3.98356728 | 6.53140563 | 2.74E-08 | 2.50E-06 | 8.8961859 |
| IGKV1-16 | 2.34852639 | 1.59807531 | 4.09720818 | 1.47E-04 | 0.00247206 | 0.88606511 |
| IGLV10-54 | 2.3334717 | -0.5748286 | 3.56143133 | 7.98E-04 | 0.0089113 | -0.626554 |
| SAMD4A | 2.32625777 | 4.63716103 | 7.91485737 | 1.71E-10 | 4.35E-08 | 13.7917981 |
| HIST1H1B | 2.32489363 | 4.79850015 | 7.07085823 | 3.78E-09 | 5.21E-07 | 10.7923225 |
| IGKV1-5 | 2.32338491 | 4.10348926 | 4.98624212 | 7.20E-06 | 2.20E-04 | 3.53310286 |
| CTSL | 2.31825609 | 2.9198531 | 5.34346864 | 2.04E-06 | 8.10E-05 | 4.81548884 |
| RP11-320N7.1 | 2.31540192 | -1.5519437 | 3.72989502 | 4.74E-04 | 0.00599071 | -0.5435652 |
| CXCR2P1 | 2.31115633 | 6.08080437 | 6.75648843 | 1.20E-08 | 1.29E-06 | 9.6495621 |
| MYOF | 2.30654507 | 5.12297532 | 7.67777357 | 4.08E-10 | 9.20E-08 | 12.9518362 |
| ZNF219 | -2.3044293 | 3.58057228 | -8.8369085 | 6.11E-12 | 3.38E-09 | 16.9679525 |
| CXCL6 | -2.3017708 | -1.4948266 | -4.1017272 | 1.44E-04 | 0.00244775 | 0.58989948 |
| CDK1 | 2.29736516 | 1.02024771 | 4.58337563 | 2.89E-05 | 6.85E-04 | 2.35125263 |
| IGLV1-40 | 2.29062125 | 3.48599448 | 6.23828978 | 8.02E-08 | 5.81E-06 | 7.87689996 |
| EPHB2 | 2.29011517 | 3.54064975 | 5.31590311 | 2.25E-06 | 8.77E-05 | 4.7087989 |
| HIST1H3F | 2.28492412 | 3.23323918 | 5.86196723 | 3.16E-07 | 1.78E-05 | 6.56888366 |
| FAM26F | 2.26388881 | 3.37183012 | 3.56612406 | 7.87E-04 | 0.00882628 | -0.8303509 |
| FCGR1CP | 2.25906882 | 2.96309195 | 4.68771381 | 2.03E-05 | 5.12E-04 | 2.62703357 |
| JUP | 2.24367328 | 5.25224482 | 4.5312746 | 3.46E-05 | 7.86E-04 | 1.96843973 |
| IGLV1-51 | 2.23511494 | 3.29989492 | 5.424842 | 1.53E-06 | 6.34E-05 | 5.06196211 |
| OR52V1P | 2.22910056 | -0.5817499 | 3.93585728 | 2.47E-04 | 0.00365722 | 0.2356821 |
| 4-Sep | 2.2242856 | 4.45597469 | 5.76171443 | 4.54E-07 | 2.33E-05 | 6.16825172 |
| TMEM255A | 2.22348147 | 2.31940815 | 5.984613 | 2.02E-07 | 1.23E-05 | 6.9907678 |
| IGKV4-1 | 2.21840621 | 4.34767543 | 4.96165612 | 7.85E-06 | 2.36E-04 | 3.42980606 |
| RP11-34P1.2 | -2.2112799 | -1.3692495 | -4.2945386 | 7.65E-05 | 0.00149143 | 1.07903506 |
| MTATP6P26 | 2.21104858 | 1.18368267 | 4.4493475 | 4.56E-05 | 9.90E-04 | 1.94306952 |
| CDC25A | 2.19243373 | 0.29445638 | 4.49336386 | 3.93E-05 | 8.73E-04 | 2.02799608 |
| IFIT5 | 2.18815584 | 7.27915175 | 6.85132256 | 8.47E-09 | 9.70E-07 | 9.99287441 |
| SF3A3P2 | 2.18139704 | -0.4201798 | 3.73630068 | 4.65E-04 | 0.00592369 | -0.2616495 |
| IGHV2-5 | 2.17699082 | 1.56947571 | 3.86561169 | 3.09E-04 | 0.00430165 | 0.20099454 |
| KLHDC7B | 2.17647565 | 3.61523963 | 7.63173817 | 4.82E-10 | 1.04E-07 | 12.7698182 |
| RP11-34P1.1 | -2.1721503 | 2.08489632 | -6.1099596 | 1.28E-07 | 8.60E-06 | 7.40849504 |
| TYMS | 2.16485003 | 2.67559134 | 6.96465811 | 5.58E-09 | 6.99E-07 | 10.3865184 |
| CEP55 | 2.16457586 | 0.64420332 | 4.88883549 | 1.01E-05 | 2.91E-04 | 3.23708284 |
| IGLV2-11 | 2.16452561 | 3.00546009 | 3.92214747 | 2.58E-04 | 0.00378553 | 0.24622385 |
| IGHA1 | 2.16353289 | 6.92576824 | 4.64495133 | 2.35E-05 | 5.79E-04 | 2.3167307 |
| IGLL5 | 2.16242541 | 4.60854851 | 5.04027852 | 5.96E-06 | 1.88E-04 | 3.66975684 |
| IGLV2-14 | 2.16188976 | 4.30051558 | 4.30046053 | 7.50E-05 | 0.00146938 | 1.27227427 |
| GPR84 | 2.16027588 | 1.28519451 | 5.31061267 | 2.29E-06 | 8.87E-05 | 4.68219082 |
| AIM2 | 2.14532527 | 4.85562615 | 6.90494572 | 6.95E-09 | 8.11E-07 | 10.2000613 |
| CDCA2 | 2.14349993 | 0.72505424 | 4.31145093 | 7.23E-05 | 0.00143004 | 1.51675868 |
| CTD-3093B17.2 | -2.1345932 | 0.4212716 | -5.5941581 | 8.31E-07 | 3.88E-05 | 5.38754475 |
| TOP2A | 2.12787858 | 2.78394874 | 5.85600722 | 3.23E-07 | 1.79E-05 | 6.55984306 |
| TIGD3 | -2.1217665 | 2.90097818 | -6.415154 | 4.20E-08 | 3.46E-06 | 8.49770542 |
| ANXA10 | 2.11989263 | -0.112672 | 3.8862974 | 2.89E-04 | 0.0040997 | 0.20072331 |
| STAT1 | 2.11711879 | 8.44139426 | 8.86913478 | 5.45E-12 | 3.23E-09 | 17.1277628 |
| IGKV3-11 | 2.11678506 | 3.91017576 | 4.72665201 | 1.77E-05 | 4.59E-04 | 2.68419403 |
| ISL2 | -2.1116328 | 1.36065377 | -4.4236983 | 4.97E-05 | 0.00106047 | 1.87962601 |
| PARP9 | 2.1102333 | 8.81203982 | 7.71136707 | 3.61E-10 | 8.36E-08 | 13.0727049 |
| LGALS3BP | 2.10965002 | 5.59434398 | 7.22017778 | 2.18E-09 | 3.31E-07 | 11.3101339 |
| CTD-2521M24.13 | 2.10210066 | 1.7816248 | 6.40701601 | 4.33E-08 | 3.53E-06 | 8.33949413 |
| CD38 | 2.09619948 | 6.35698295 | 7.52506286 | 7.13E-10 | 1.46E-07 | 12.3971501 |
| IGHA2 | 2.09415661 | 5.53451566 | 3.69077786 | 5.36E-04 | 0.00656058 | -0.6695205 |
| CCNJL | -2.0925346 | 5.31839834 | -5.9919739 | 1.97E-07 | 1.21E-05 | 6.92974004 |
| TPX2 | 2.08867745 | 2.78672005 | 6.46304854 | 3.52E-08 | 3.06E-06 | 8.65595164 |
| TRIM6 | 2.0874742 | 3.12165067 | 5.60494288 | 8.00E-07 | 3.77E-05 | 5.69894641 |
| TMEM51 | 2.08702108 | 0.63669468 | 4.33210258 | 6.75E-05 | 0.00135927 | 1.54864 |
| PARP14 | 2.08691995 | 10.0709856 | 7.67516397 | 4.12E-10 | 9.20E-08 | 12.9304465 |
| CDC42P3 | 2.0780495 | -0.8968121 | 5.10938784 | 4.67E-06 | 1.53E-04 | 3.08686988 |
| ADGRE3 | -2.0735476 | 6.17986173 | -6.8872191 | 7.42E-09 | 8.60E-07 | 10.1124465 |
| AXL | 2.06988458 | 2.90081738 | 5.86096932 | 3.17E-07 | 1.78E-05 | 6.5727675 |
| IGHM | 2.0690847 | 8.63063996 | 4.31235717 | 7.21E-05 | 0.00142996 | 1.34667436 |
| HIST2H3A | 2.05998215 | 4.36759885 | 7.44502691 | 9.56E-10 | 1.78E-07 | 12.1292231 |
| HIST2H3C | 2.05998215 | 4.36759885 | 7.44502691 | 9.56E-10 | 1.78E-07 | 12.1292231 |
| IGKV3-15 | 2.0561001 | 3.46282213 | 4.59892385 | 2.75E-05 | 6.61E-04 | 2.29702108 |
| HIST1H2BI | 2.0535202 | 3.3921485 | 6.94011995 | 6.11E-09 | 7.40E-07 | 10.3419439 |
| TXNDC5 | 2.04529411 | 6.35591419 | 5.85816385 | 3.20E-07 | 1.79E-05 | 6.45079853 |
| CES1 | 2.03646806 | 3.95809228 | 3.54311841 | 8.44E-04 | 0.0092913 | -0.9777235 |
| ZWINT | 2.03032271 | 1.22462144 | 5.25020294 | 2.84E-06 | 1.04E-04 | 4.47837075 |
| IGLV3-19 | 2.03000223 | 2.60219877 | 4.0291319 | 1.83E-04 | 0.00291717 | 0.59997405 |
| VAMP5 | 2.02164807 | 4.50184279 | 7.02095143 | 4.54E-09 | 5.88E-07 | 10.6165854 |
| HIST1H2BL | 2.01038643 | 2.4435397 | 6.08712336 | 1.39E-07 | 9.08E-06 | 7.34501698 |
| RGL1 | 2.00932098 | 3.57931068 | 5.38114807 | 1.78E-06 | 7.22E-05 | 4.91620729 |
| TNFAIP6 | 2.00069877 | 5.67688404 | 5.16101121 | 3.90E-06 | 1.33E-04 | 4.0377132 |

**Supplemental Table 6.** Differentially expressed genes day 10 post infection

| **geneID** | **logFC** | **AveExpr** | **t** | **P.Value** | **adj.P.Val** | **B** |
| --- | --- | --- | --- | --- | --- | --- |
| IFI27 | 6.38312492 | 6.52857751 | 7.7690393 | 2.92E-10 | 2.90E-08 | 13.2686858 |
| IGLV3-1 | 6.32205981 | 3.64601343 | 11.8197316 | 2.31E-16 | 2.16E-13 | 26.9233201 |
| IGKV1-39 | 5.81353786 | -2.3323798 | 4.32352674 | 6.95E-05 | 9.40E-04 | 1.32207567 |
| IGHG1 | 5.75999438 | 5.20630833 | 7.84746478 | 2.19E-10 | 2.25E-08 | 13.5117656 |
| SDC1 | 5.75300457 | -1.0985613 | 6.64980311 | 1.78E-08 | 9.42E-07 | 8.56624828 |
| GLDC | 5.53110257 | -3.0538121 | 4.95107542 | 8.14E-06 | 1.66E-04 | 2.70085858 |
| IGLV4-69 | 5.52213543 | 1.84852262 | 6.99388893 | 5.02E-09 | 3.23E-07 | 10.5330076 |
| IGHG3 | 5.16428194 | 1.63487172 | 6.27884929 | 6.92E-08 | 3.01E-06 | 8.0219514 |
| JCHAIN | 5.09968206 | 6.99845032 | 9.68811729 | 3.01E-13 | 9.42E-11 | 19.9782771 |
| IGLV3-25 | 5.04098024 | 2.68747646 | 7.65054548 | 4.50E-10 | 4.17E-08 | 12.8513625 |
| HIST1H3G | 5.02970621 | 3.67375764 | 13.7061093 | 6.52E-19 | 8.34E-15 | 32.6090322 |
| IGLV1-47 | 5.02686973 | 3.55912851 | 8.05738579 | 1.02E-10 | 1.16E-08 | 14.2809829 |
| BHLHA15 | 4.86220094 | -0.6193916 | 7.42319103 | 1.04E-09 | 8.61E-08 | 11.3909544 |
| IGLV6-57 | 4.82492834 | 1.76606675 | 8.76468846 | 7.92E-12 | 1.40E-09 | 16.7011023 |
| IGLC1 | 4.8137688 | 5.14650159 | 9.36866817 | 9.24E-13 | 2.55E-10 | 18.8742373 |
| HIST1H2AJ | 4.7722249 | 3.43481868 | 12.1206671 | 8.77E-17 | 1.03E-13 | 27.8872066 |
| IGLV3-21 | 4.7710939 | 3.35661179 | 8.32854067 | 3.81E-11 | 5.23E-09 | 15.2497865 |
| IGLV1-40 | 4.74307332 | 3.48599448 | 12.991606 | 5.70E-18 | 2.14E-14 | 30.5480265 |
| IGLC2 | 4.74148965 | 6.58554216 | 9.40342457 | 8.17E-13 | 2.31E-10 | 18.9925372 |
| IGLV3-10 | 4.71859851 | 0.63892604 | 5.73712481 | 4.97E-07 | 1.59E-05 | 6.14701363 |
| HIST1H3J | 4.7184777 | 3.03059472 | 12.4730581 | 2.87E-17 | 5.39E-14 | 28.9090363 |
| HIST1H3C | 4.66829636 | 2.48037715 | 8.94498523 | 4.15E-12 | 8.14E-10 | 17.389629 |
| IGKV1D-39 | 4.64593527 | 3.86533446 | 11.0084067 | 3.31E-15 | 2.07E-12 | 24.3868137 |
| IGKV3-20 | 4.6421159 | 4.84230282 | 8.2844135 | 4.47E-11 | 5.91E-09 | 15.0579467 |
| IGLC3 | 4.62224347 | 5.19149196 | 7.2839585 | 1.73E-09 | 1.30E-07 | 11.4569285 |
| IGLV2-23 | 4.62203607 | 2.85540372 | 7.4296821 | 1.01E-09 | 8.50E-08 | 12.0627417 |
| IGHV4-59 | 4.61462638 | 2.73953349 | 7.25218843 | 1.94E-09 | 1.42E-07 | 11.4283221 |
| RRM2 | 4.58939731 | 3.3856448 | 12.0330629 | 1.16E-16 | 1.21E-13 | 27.6044504 |
| IGHV3-66 | 4.56036281 | -0.7793606 | 4.91853447 | 9.12E-06 | 1.81E-04 | 3.32820701 |
| HIST1H3B | 4.55452232 | 3.98356728 | 12.3931028 | 3.69E-17 | 6.30E-14 | 28.7777379 |
| UCHL1 | 4.55034964 | -3.0329491 | 7.06854216 | 3.81E-09 | 2.52E-07 | 8.19077426 |
| PBK | 4.47899285 | -1.0518386 | 7.49560046 | 7.95E-10 | 6.88E-08 | 11.2282992 |
| IGKV1-9 | 4.459992 | 1.82161954 | 7.31367302 | 1.55E-09 | 1.19E-07 | 11.6641668 |
| HIST1H2BB | 4.44660111 | 0.15142088 | 8.8558598 | 5.71E-12 | 1.06E-09 | 16.4011609 |
| DHFRP1 | 4.43943073 | -3.9535354 | 4.27283254 | 8.22E-05 | 0.00108087 | 0.80995517 |
| IGLV1-44 | 4.42812683 | 3.2475498 | 8.41185849 | 2.82E-11 | 4.15E-09 | 15.5439106 |
| IGHV5-51 | 4.42538573 | 2.66250532 | 7.28219588 | 1.74E-09 | 1.31E-07 | 11.5345514 |
| CAV1 | 4.4044619 | -0.3923474 | 6.84149065 | 8.78E-09 | 5.20E-07 | 9.72592966 |
| HIST1H2AB | 4.40432876 | 1.9982149 | 10.3096243 | 3.51E-14 | 1.40E-11 | 21.9415717 |
| HIST1H1B | 4.39878089 | 4.79850015 | 12.5545543 | 2.22E-17 | 4.68E-14 | 29.304572 |
| IGKV6-21 | 4.38198283 | -0.9277962 | 5.27879837 | 2.57E-06 | 6.32E-05 | 4.3434171 |
| IGKV2-28 | 4.37573481 | 2.38090744 | 6.66912373 | 1.65E-08 | 8.90E-07 | 9.37007963 |
| IGKV1-5 | 4.36423023 | 4.10348926 | 9.11446635 | 2.27E-12 | 4.96E-10 | 17.9944934 |
| HIST1H2AH | 4.35645883 | 3.07116528 | 12.3592667 | 4.11E-17 | 6.43E-14 | 28.5990162 |
| IGHV4-39 | 4.35146275 | 2.64502212 | 6.78203306 | 1.09E-08 | 6.24E-07 | 9.7438942 |
| IGLV2-8 | 4.3506902 | 2.72865308 | 8.21187579 | 5.82E-11 | 7.19E-09 | 14.8413395 |
| MZB1 | 4.33963814 | 4.32432781 | 10.4840207 | 1.94E-14 | 8.96E-12 | 22.6698596 |
| HIST1H2BM | 4.33549545 | 2.04863383 | 10.9822725 | 3.61E-15 | 2.19E-12 | 24.0647624 |
| IGLV3-27 | 4.27895762 | -0.2352673 | 5.32094411 | 2.21E-06 | 5.59E-05 | 4.6782186 |
| AC136616.2 | 4.27504154 | -3.0281087 | 4.58795727 | 2.85E-05 | 4.57E-04 | 1.70732227 |
| IGLV2-11 | 4.25932324 | 3.00546009 | 8.03659812 | 1.10E-10 | 1.24E-08 | 14.2129565 |
| IGHV3-21 | 4.24281015 | 2.96203594 | 6.98347165 | 5.21E-09 | 3.33E-07 | 10.4372316 |
| SH3RF2 | 4.23321479 | 0.19799817 | 6.87066523 | 7.89E-09 | 4.75E-07 | 9.97508935 |
| CDC45 | 4.22823504 | -0.0135068 | 8.40334675 | 2.91E-11 | 4.23E-09 | 14.6505381 |
| IGLV8-61 | 4.19281237 | 1.63143491 | 5.5655462 | 9.21E-07 | 2.65E-05 | 5.52473079 |
| IGHA1 | 4.17904678 | 6.92576824 | 7.96326157 | 1.44E-10 | 1.55E-08 | 13.9029144 |
| IGLC7 | 4.16481027 | -1.8091036 | 5.61825754 | 7.62E-07 | 2.26E-05 | 5.07170655 |
| IGKC | 4.13226016 | 8.62410501 | 8.98689469 | 3.58E-12 | 7.38E-10 | 17.5579964 |
| IGLV9-49 | 4.12737237 | -0.1138147 | 6.15207193 | 1.10E-07 | 4.51E-06 | 7.47322839 |
| IGHV3-48 | 4.10284603 | 0.68988075 | 4.4306869 | 4.85E-05 | 7.11E-04 | 1.85462653 |
| IGHV4-4 | 4.09411382 | -0.3331091 | 4.14069962 | 1.27E-04 | 0.00154543 | 1.02771243 |
| IGKV1-37 | 4.08401501 | -2.1332425 | 5.70671658 | 5.54E-07 | 1.73E-05 | 5.2956703 |
| IGLV4-60 | 4.07595612 | -0.5858216 | 3.73848277 | 4.61E-04 | 0.00425648 | -0.1105403 |
| IGKV1-12 | 4.06239249 | 1.92940559 | 8.23620718 | 5.33E-11 | 6.67E-09 | 14.9185204 |
| STXBP5L | 4.01970577 | -2.4707557 | 5.67346435 | 6.25E-07 | 1.92E-05 | 5.03834904 |
| HIST1H3F | 4.01323959 | 3.23323918 | 10.551567 | 1.54E-14 | 7.41E-12 | 22.8806675 |
| IGKV6D-21 | 4.00750525 | -2.0730657 | 3.69318415 | 5.32E-04 | 0.00477167 | -0.3529731 |
| IGHV3-53 | 4.00748379 | 0.52583044 | 4.94816082 | 8.23E-06 | 1.67E-04 | 3.51948075 |
| TXNDC5 | 3.99898946 | 6.35591419 | 10.2805782 | 3.88E-14 | 1.52E-11 | 21.9869975 |
| HIST1H2BL | 3.91357402 | 2.4435397 | 12.6739635 | 1.53E-17 | 4.68E-14 | 29.4508775 |
| RP11-290L7.3 | -3.9108817 | -3.3051161 | -4.1742917 | 1.14E-04 | 0.00141684 | 0.56709507 |
| IGHV1-24 | 3.91050981 | -0.3408272 | 4.21812282 | 9.86E-05 | 0.0012644 | 1.26009302 |
| IGHV3-30 | 3.90746368 | 2.20049391 | 4.34041737 | 6.57E-05 | 9.01E-04 | 1.3550456 |
| IGLV1-36 | 3.89940325 | -0.2368439 | 4.75957232 | 1.58E-05 | 2.83E-04 | 2.92494174 |
| IGLV5-37 | 3.89789957 | -2.5713574 | 4.3481063 | 6.40E-05 | 8.84E-04 | 1.41815708 |
| HIST1H2AL | 3.89385794 | 2.96639264 | 12.1557391 | 7.84E-17 | 9.81E-14 | 27.9641924 |
| IGKV1-16 | 3.87723161 | 1.59807531 | 7.13977435 | 2.93E-09 | 2.03E-07 | 11.0483607 |
| HIST1H2BO | 3.87665133 | 3.59352657 | 12.5554371 | 2.21E-17 | 4.68E-14 | 29.2512476 |
| IGHV2-5 | 3.86729003 | 1.56947571 | 7.31339381 | 1.55E-09 | 1.19E-07 | 11.663057 |
| IGKV1-33 | 3.86351427 | 3.06362565 | 8.23677931 | 5.32E-11 | 6.67E-09 | 14.9197783 |
| IGHV6-1 | 3.8589349 | 0.14197451 | 7.35929313 | 1.31E-09 | 1.03E-07 | 11.6146403 |
| IGHV4-61 | 3.85200683 | 0.2486952 | 5.53135379 | 1.04E-06 | 2.96E-05 | 5.45499438 |
| IGHV3-13 | 3.84845687 | -0.0277646 | 5.79716799 | 4.00E-07 | 1.32E-05 | 6.31601378 |
| IGKV1-6 | 3.84083956 | -0.5517574 | 4.98686163 | 7.19E-06 | 1.50E-04 | 3.61630536 |
| IGHV3-74 | 3.84058097 | 1.46382789 | 5.83859218 | 3.44E-07 | 1.17E-05 | 6.48554062 |
| CDCA5 | 3.81652262 | 0.87812563 | 9.8027049 | 2.02E-13 | 6.54E-11 | 19.8807763 |
| HIST1H2BI | 3.8115772 | 3.3921485 | 13.2677303 | 2.45E-18 | 1.15E-14 | 31.3883707 |
| IGLL5 | 3.81004943 | 4.60854851 | 8.51390927 | 1.95E-11 | 3.03E-09 | 15.861932 |
| CDK1 | 3.80743268 | 1.02024771 | 8.01836776 | 1.18E-10 | 1.30E-08 | 14.0299576 |
| IGLV1-51 | 3.80294583 | 3.29989492 | 9.45362417 | 6.85E-13 | 1.98E-10 | 19.1747808 |
| IGHV1-2 | 3.7939725 | 1.64108715 | 5.20526412 | 3.33E-06 | 7.91E-05 | 4.27212857 |
| IGKV4-1 | 3.77486255 | 4.34767543 | 8.2441729 | 5.18E-11 | 6.61E-09 | 14.9098084 |
| IGHV1-18 | 3.75950412 | 2.3874645 | 7.5521768 | 6.46E-10 | 5.75E-08 | 12.4984421 |
| IGLV3-19 | 3.75795773 | 2.60219877 | 7.89975228 | 1.81E-10 | 1.91E-08 | 13.7337281 |
| IGHV1-69-2 | 3.75772011 | 1.51360155 | 4.67657682 | 2.11E-05 | 3.57E-04 | 2.48886 |
| IGHV3-15 | 3.74197391 | 1.99915113 | 6.82846851 | 9.21E-09 | 5.42E-07 | 9.93608677 |
| CEP55 | 3.72238282 | 0.64420332 | 8.93106739 | 4.37E-12 | 8.37E-10 | 16.9877408 |
| IGLV2-14 | 3.71978559 | 4.30051558 | 7.24256597 | 2.01E-09 | 1.46E-07 | 11.30678 |
| IGHV3-33 | 3.70793144 | 3.39467328 | 6.6418722 | 1.83E-08 | 9.65E-07 | 9.17174623 |
| IGKV2D-29 | 3.68355382 | -0.0713615 | 5.73439054 | 5.01E-07 | 1.59E-05 | 6.05966724 |
| IGHV2-26 | 3.67375247 | -1.3074046 | 4.82756507 | 1.25E-05 | 2.33E-04 | 3.03658753 |
| CCNB2 | 3.66215514 | 1.50997416 | 10.2714262 | 4.00E-14 | 1.53E-11 | 21.7375609 |
| IGHV3-23 | 3.64585195 | 3.33241739 | 7.18167745 | 2.52E-09 | 1.81E-07 | 11.1212799 |
| IGHV1-69 | 3.64576034 | -0.6317906 | 3.46477134 | 0.00107044 | 0.0083449 | -0.8882441 |
| IGKV2D-30 | 3.63785117 | -2.7447393 | 4.90853993 | 9.44E-06 | 1.87E-04 | 3.02249434 |
| CDCA2 | 3.63208771 | 0.72505424 | 7.74618565 | 3.17E-10 | 3.09E-08 | 13.0365763 |
| CH17-212P11.4 | 3.63068456 | -2.6696926 | 4.23404297 | 9.35E-05 | 0.00121038 | 0.97836636 |
| ZWINT | 3.62251808 | 1.22462144 | 9.99735128 | 1.03E-13 | 3.39E-11 | 20.7712511 |
| TOP2A | 3.61375777 | 2.78394874 | 10.4696048 | 2.03E-14 | 8.96E-12 | 22.5801497 |
| TYMS | 3.61135827 | 2.67559134 | 12.2551708 | 5.71E-17 | 8.25E-14 | 28.2229864 |
| IGHV1-46 | 3.60555299 | 1.67322902 | 7.69775721 | 3.79E-10 | 3.58E-08 | 13.0252432 |
| IGKV3-11 | 3.60380752 | 3.91017576 | 8.08048239 | 9.38E-11 | 1.08E-08 | 14.336402 |
| IGHV3-43 | 3.59249738 | 0.24369062 | 6.04623658 | 1.62E-07 | 6.25E-06 | 7.18532053 |
| IGKV1-27 | 3.58649565 | 1.06369302 | 6.05429695 | 1.57E-07 | 6.14E-06 | 7.24097868 |
| IGKV2D-28 | 3.58255386 | -0.2691291 | 4.11545864 | 1.38E-04 | 0.00165489 | 0.95156943 |
| KIFC1 | 3.55971697 | 1.43844572 | 9.35639019 | 9.65E-13 | 2.59E-10 | 18.6421252 |
| DERL3 | 3.54083211 | 2.35441094 | 8.31382541 | 4.02E-11 | 5.47E-09 | 15.2008689 |
| IGKV3-15 | 3.5387231 | 3.46282213 | 8.09994037 | 8.74E-11 | 1.02E-08 | 14.4170964 |
| IGHV3-7 | 3.51237486 | 2.30005838 | 7.27841165 | 1.76E-09 | 1.32E-07 | 11.5276183 |
| IGLV7-43 | 3.50762706 | 1.10905251 | 6.20708721 | 8.99E-08 | 3.77E-06 | 7.77145596 |
| IGKV2-30 | 3.50079744 | 1.8804678 | 6.52950781 | 2.76E-08 | 1.37E-06 | 8.8911476 |
| DEPDC1 | 3.49654229 | 0.52899406 | 7.41982484 | 1.05E-09 | 8.68E-08 | 11.8483728 |
| IGHV3-11 | 3.49615757 | 1.31651774 | 6.59489508 | 2.17E-08 | 1.11E-06 | 9.12583507 |
| CDC25A | 3.49399137 | 0.29445638 | 7.5524274 | 6.45E-10 | 5.75E-08 | 12.3148886 |
| HIST1H2AI | 3.46477152 | 3.93962279 | 11.4632516 | 7.36E-16 | 5.53E-13 | 25.8790847 |
| IGLV4-3 | 3.45731777 | -2.7774833 | 5.76191381 | 4.54E-07 | 1.46E-05 | 5.22771684 |
| KIF4A | 3.44198187 | 0.78251322 | 7.43223144 | 1.00E-09 | 8.48E-08 | 11.9635632 |
| IGLV3-16 | 3.40343759 | -1.5877071 | 4.87216874 | 1.07E-05 | 2.07E-04 | 3.17802137 |
| HIST1H2BE | 3.35927833 | 5.28185819 | 12.0983124 | 9.42E-17 | 1.04E-13 | 27.9090888 |
| MKI67 | 3.35835104 | 4.63253904 | 8.11361993 | 8.31E-11 | 9.76E-09 | 14.429896 |
| SIGLEC1 | 3.35324773 | 6.35550789 | 5.64244394 | 6.99E-07 | 2.10E-05 | 5.59137575 |
| IGLV10-54 | 3.35027408 | -0.5748286 | 5.34995823 | 1.99E-06 | 5.10E-05 | 4.7744661 |
| E2F7 | 3.34519188 | 0.26506285 | 8.1997375 | 6.08E-11 | 7.41E-09 | 14.5582638 |
| HIST1H2AM | 3.33751414 | 4.14408399 | 13.3526559 | 1.89E-18 | 1.15E-14 | 31.7040705 |
| CDCA7 | 3.32851859 | 1.96972485 | 10.8619546 | 5.40E-15 | 3.05E-12 | 23.7518194 |
| LYPD2 | -3.3215982 | -0.8883201 | -3.9683795 | 2.22E-04 | 0.00240643 | 0.41728074 |
| BIRC5 | 3.30268172 | 1.65275431 | 8.62815725 | 1.29E-11 | 2.15E-09 | 16.2622844 |
| IGKV1-17 | 3.30111706 | 0.93588413 | 5.43328981 | 1.48E-06 | 3.94E-05 | 5.11868633 |
| IGLV1-41 | 3.30035434 | -2.714945 | 5.9947324 | 1.95E-07 | 7.31E-06 | 6.0930961 |
| CENPA | 3.29297325 | -0.0367356 | 10.6493655 | 1.11E-14 | 5.61E-12 | 21.8758032 |
| NRXN1 | -3.2909347 | 0.60187682 | -4.7517179 | 1.63E-05 | 2.88E-04 | 2.89063351 |
| IGKV5-2 | 3.28647529 | -0.7224096 | 6.82560676 | 9.31E-09 | 5.45E-07 | 9.6022257 |
| CCNA2 | 3.27754857 | 2.55561192 | 10.4781739 | 1.98E-14 | 8.96E-12 | 22.5808652 |
| HIST2H3A | 3.25966129 | 4.36759885 | 11.6888749 | 3.52E-16 | 2.88E-13 | 26.6047235 |
| HIST2H3C | 3.25966129 | 4.36759885 | 11.6888749 | 3.52E-16 | 2.88E-13 | 26.6047235 |
| HRASLS2 | 3.22690817 | -1.6573674 | 6.19372065 | 9.44E-08 | 3.93E-06 | 7.17559601 |
| IGLV7-46 | 3.20919959 | 1.1620273 | 5.96638321 | 2.16E-07 | 7.86E-06 | 6.93560656 |
| ASPM | 3.16853446 | 2.97679305 | 8.96899538 | 3.81E-12 | 7.70E-10 | 17.4966417 |
| E2F8 | 3.16301322 | 1.01655749 | 8.28263133 | 4.50E-11 | 5.91E-09 | 15.0026196 |
| TPX2 | 3.15750567 | 2.78672005 | 10.2318259 | 4.58E-14 | 1.65E-11 | 21.7892653 |
| CICP19 | -3.1176447 | -0.7072186 | -3.3883765 | 0.00134605 | 0.00996017 | -1.0337389 |
| IGLV5-45 | 3.11642718 | -0.1079216 | 5.07283004 | 5.32E-06 | 1.17E-04 | 3.92940733 |
| MTND4LP12 | -3.0993789 | -1.973906 | -4.0016187 | 2.00E-04 | 0.00221823 | 0.51097089 |
| HIST1H2BH | 3.08555625 | 4.11742223 | 13.6027212 | 8.88E-19 | 8.34E-15 | 32.444071 |
| HIST1H4A | 3.0855357 | 2.53563859 | 10.3472043 | 3.09E-14 | 1.26E-11 | 22.1929318 |
| AC233755.2 | 3.08551405 | -3.1866988 | 3.72301239 | 4.84E-04 | 0.00442616 | -0.3222203 |
| DLGAP5 | 3.06541368 | 2.28198991 | 9.08393698 | 2.53E-12 | 5.47E-10 | 17.8737465 |
| IGKV2-24 | 3.05484313 | 0.97021596 | 5.51079767 | 1.12E-06 | 3.13E-05 | 5.37966219 |
| IGHM | 3.04205169 | 8.63063996 | 5.93072667 | 2.46E-07 | 8.71E-06 | 6.65456691 |
| MYBL2 | 3.03646696 | 4.06950454 | 7.31235393 | 1.56E-09 | 1.19E-07 | 11.560552 |
| IGHV3-49 | 3.02722021 | 0.3535845 | 3.71850344 | 4.91E-04 | 0.00447369 | -0.2913452 |
| NCAPG | 3.01827397 | 2.46085441 | 9.27782134 | 1.27E-12 | 3.19E-10 | 18.5545615 |
| BCAN | 3.01792479 | -1.2353288 | 4.9034279 | 9.61E-06 | 1.89E-04 | 3.2151018 |
| IGKV3OR2-268 | 3.01428485 | -1.7094586 | 3.90273985 | 2.75E-04 | 0.00284681 | 0.28976476 |
| IGHV2-70 | 2.99502925 | -0.0590726 | 4.66409946 | 2.20E-05 | 3.70E-04 | 2.61778528 |
| CCNB1 | 2.99388924 | 1.964688 | 9.48176339 | 6.20E-13 | 1.82E-10 | 19.2159732 |
| KCTD14 | 2.99106558 | 0.65016798 | 3.39212199 | 0.0013311 | 0.0098848 | -1.0308763 |
| IGHJ3 | 2.97979408 | -0.8211757 | 4.42334235 | 4.98E-05 | 7.24E-04 | 1.87471641 |
| HIST2H4A | 2.96655643 | 5.74143261 | 11.5987733 | 4.72E-16 | 3.70E-13 | 26.3253366 |
| IGKV3D-20 | 2.94066755 | -0.5788167 | 4.52939822 | 3.48E-05 | 5.35E-04 | 2.1982835 |
| PTTG1 | 2.93498622 | 1.83890267 | 10.6501361 | 1.10E-14 | 5.61E-12 | 23.0762567 |
| PYCR1 | 2.92640378 | 0.007831 | 7.49251892 | 8.04E-10 | 6.92E-08 | 12.1525487 |
| HIST1H2AK | 2.91723261 | 2.19417492 | 9.72703632 | 2.63E-13 | 8.36E-11 | 20.093571 |
| IGKV1D-16 | 2.89999299 | -1.5975645 | 4.17281752 | 1.14E-04 | 0.0014203 | 1.06230751 |
| CFAP46 | -2.8993539 | -2.2433815 | -3.6620255 | 5.86E-04 | 0.00511711 | -0.4244094 |
| IGHV1-58 | 2.89171737 | -1.5157219 | 4.47127033 | 4.23E-05 | 6.36E-04 | 2.00453095 |
| HIST1H4L | 2.86161703 | 2.08904132 | 8.13460553 | 7.70E-11 | 9.16E-09 | 14.5683726 |
| IGLV2-18 | 2.85626652 | -0.5416247 | 4.11081498 | 1.40E-04 | 0.00167377 | 0.93469905 |
| IGHV3-73 | 2.8091942 | -0.5238404 | 5.28269974 | 2.53E-06 | 6.26E-05 | 4.58886592 |
| HMMR | 2.8066658 | 0.96168847 | 7.25764559 | 1.90E-09 | 1.41E-07 | 11.4403313 |
| GSG2 | 2.79168919 | 0.58411391 | 8.02518146 | 1.15E-10 | 1.27E-08 | 14.0330273 |
| HIST1H2AG | 2.78882516 | 5.28925792 | 11.4307731 | 8.18E-16 | 5.91E-13 | 25.7837837 |
| TSHR | 2.75639641 | 1.13567572 | 7.71457958 | 3.56E-10 | 3.41E-08 | 13.0475569 |
| MTND5P24 | -2.7471218 | -0.0506884 | -4.5595351 | 3.14E-05 | 4.93E-04 | 2.29404491 |
| CTD-2116N17.1 | 2.74651018 | 2.05053384 | 10.4670958 | 2.05E-14 | 8.96E-12 | 22.5432711 |
| TNFRSF17 | 2.73929932 | 2.50952914 | 10.0653486 | 8.12E-14 | 2.83E-11 | 21.22966 |
| MTND4P22 | -2.7340483 | -0.4789305 | -4.827005 | 1.25E-05 | 2.34E-04 | 3.13399091 |
| IGHA2 | 2.72831479 | 5.53451566 | 4.6710346 | 2.15E-05 | 3.62E-04 | 2.15990048 |
| MTCYBP11 | -2.7187548 | -0.2887552 | -4.8334738 | 1.23E-05 | 2.30E-04 | 3.15779431 |
| PHF24 | -2.7177968 | -1.2092825 | -3.7651321 | 4.24E-04 | 0.00399975 | -0.0408635 |
| BUB1 | 2.70431859 | 2.8262914 | 9.40053716 | 8.26E-13 | 2.31E-10 | 18.9854887 |
| RP11-49K24.6 | 2.70270778 | -1.808946 | 3.46165002 | 0.00108056 | 0.00840636 | -0.8462955 |
| IFI44L | 2.69964959 | 9.26563843 | 5.77332902 | 4.36E-07 | 1.42E-05 | 5.97479496 |
| HIST1H2BJ | 2.69629202 | 4.04422032 | 10.0966188 | 7.29E-14 | 2.58E-11 | 21.3667342 |
| MTCO3P5 | -2.6954876 | -0.3881875 | -4.5598758 | 3.14E-05 | 4.93E-04 | 2.29773555 |
| RBM17P2 | -2.6941516 | -2.1246294 | -5.5788485 | 8.78E-07 | 2.54E-05 | 5.26943037 |
| KLHL14 | 2.68704889 | 3.10271361 | 6.10865158 | 1.29E-07 | 5.13E-06 | 7.2503533 |
| RAD51 | 2.68069398 | 1.0832504 | 5.02700614 | 6.24E-06 | 1.34E-04 | 3.75293249 |
| MT1E | 2.68042691 | 0.67864287 | 5.67322494 | 6.25E-07 | 1.92E-05 | 5.93249527 |
| TRIP13 | 2.67354679 | 0.40554582 | 7.16082761 | 2.72E-09 | 1.92E-07 | 11.07448 |
| HIST1H2BF | 2.66421611 | 4.38446476 | 11.3807207 | 9.65E-16 | 6.71E-13 | 25.6206379 |
| IGHV3-72 | 2.65674453 | -0.6225928 | 3.71909363 | 4.90E-04 | 0.00446979 | -0.1865863 |
| MCM10 | 2.65662542 | 2.29064464 | 9.15115697 | 1.99E-12 | 4.57E-10 | 18.116563 |
| HMGB3 | 2.64846008 | 1.71827967 | 9.06457957 | 2.71E-12 | 5.79E-10 | 17.7964456 |
| GPRC5D | 2.64650991 | -0.0986747 | 6.44487515 | 3.77E-08 | 1.80E-06 | 8.53329205 |
| IGHV3-20 | 2.64375494 | -1.5229043 | 3.69236454 | 5.33E-04 | 0.00478157 | -0.2349549 |
| CDC6 | 2.63859364 | 2.04542278 | 9.24477307 | 1.43E-12 | 3.45E-10 | 18.3998936 |
| HIST1H3A | 2.63492436 | 2.32258838 | 9.22859104 | 1.52E-12 | 3.56E-10 | 18.396538 |
| HIST1H1A | 2.62934477 | -1.2568166 | 5.87754528 | 2.99E-07 | 1.04E-05 | 6.46050279 |
| IGLV2-5 | 2.624271 | -1.9000598 | 4.8501151 | 1.16E-05 | 2.20E-04 | 3.11788395 |
| IGKV1-8 | 2.61193188 | -0.1759499 | 4.14717563 | 1.24E-04 | 0.00151987 | 1.03862665 |
| USP18 | 2.59670959 | 5.2942684 | 5.44317579 | 1.43E-06 | 3.82E-05 | 4.97999036 |
| CCNE1 | 2.57957343 | 0.50340415 | 8.09636031 | 8.85E-11 | 1.03E-08 | 14.2482798 |
| CENPW | 2.57915417 | -0.5461235 | 5.3923223 | 1.71E-06 | 4.48E-05 | 4.94777618 |
| FAM72C | 2.57772954 | -2.1569904 | 3.45362899 | 0.00110699 | 0.00857646 | -0.8661754 |
| HIST1H2BN | 2.57580393 | 4.24538672 | 11.7110543 | 3.28E-16 | 2.88E-13 | 26.6798477 |
| BUB1B | 2.56832613 | 1.7744837 | 7.40701066 | 1.10E-09 | 9.06E-08 | 11.9951536 |
| CD38 | 2.56207501 | 6.35698295 | 8.94468618 | 4.16E-12 | 8.14E-10 | 17.3529003 |
| MND1 | 2.55917231 | -1.5383241 | 5.83657804 | 3.46E-07 | 1.18E-05 | 6.26998086 |
| HIST1H2AE | 2.55310213 | 4.65718637 | 10.5835619 | 1.38E-14 | 6.83E-12 | 22.9973001 |
| HIST1H3I | 2.5454351 | 3.93314492 | 10.3584634 | 2.97E-14 | 1.24E-11 | 22.2485198 |
| PDZK1IP1 | -2.5367407 | 5.77630399 | -4.8237666 | 1.27E-05 | 2.36E-04 | 2.68289558 |
| TSPEAR | -2.5259744 | -0.086084 | -4.758391 | 1.59E-05 | 2.84E-04 | 2.92057191 |
| STMN1 | 2.51481248 | 4.31863767 | 10.4598176 | 2.10E-14 | 8.97E-12 | 22.5851497 |
| IGHJ2 | 2.51400913 | -1.9347148 | 3.57465617 | 7.66E-04 | 0.00640884 | -0.5541674 |
| CENPU | 2.50299025 | 1.83085381 | 9.13507092 | 2.11E-12 | 4.72E-10 | 18.0177434 |
| IGHGP | 2.49208593 | -1.5945826 | 4.6820458 | 2.07E-05 | 3.52E-04 | 2.6403766 |
| RBM17P1 | -2.4862701 | -0.2524074 | -5.1411492 | 4.18E-06 | 9.54E-05 | 4.15490411 |
| HIST2H3D | 2.4806886 | 3.77308984 | 8.52728278 | 1.86E-11 | 2.93E-09 | 15.9119841 |
| HIST1H2BG | 2.46757178 | 3.97665457 | 11.2152728 | 1.67E-15 | 1.12E-12 | 25.0828504 |
| MTND3P9 | -2.4668603 | -2.087085 | -3.5662389 | 7.86E-04 | 0.00654119 | -0.5941747 |
| PKMYT1 | 2.4412849 | 1.09822514 | 8.33179609 | 3.77E-11 | 5.20E-09 | 15.1564657 |
| IGLJ2 | 2.44039018 | -1.5699475 | 5.16583161 | 3.83E-06 | 8.87E-05 | 4.07015264 |
| IGHG2 | 2.43943386 | 4.29847679 | 3.60252296 | 7.04E-04 | 0.00597932 | -1.1430112 |
| DTL | 2.43843266 | 2.32687192 | 6.41570439 | 4.19E-08 | 1.98E-06 | 8.43791854 |
| TAS2R40 | -2.4342009 | 0.76525918 | -4.3561436 | 6.23E-05 | 8.67E-04 | 1.62602708 |
| FABP6 | -2.4266667 | -0.7169177 | -4.4455746 | 4.62E-05 | 6.81E-04 | 1.94334015 |
| RP11-19J5.1 | -2.4130056 | -1.243562 | -3.6687756 | 5.73E-04 | 0.00503219 | -0.3074078 |
| IGLJ3 | 2.40729622 | -1.3390985 | 4.61089338 | 2.64E-05 | 4.29E-04 | 2.4511228 |
| CH17-296N19.1 | -2.3975204 | 2.01758359 | -5.8212468 | 3.66E-07 | 1.23E-05 | 6.27627732 |
| E2F1 | 2.38462267 | 2.46578772 | 8.1293553 | 7.85E-11 | 9.27E-09 | 14.5512766 |
| ITM2C | 2.38444559 | 5.20503934 | 8.62147853 | 1.32E-11 | 2.18E-09 | 16.2043369 |
| NUSAP1 | 2.37712331 | 2.08809381 | 4.48891614 | 3.99E-05 | 6.03E-04 | 1.8900654 |
| HIST1H4D | 2.37707308 | 4.3399836 | 9.26330339 | 1.34E-12 | 3.31E-10 | 18.4843867 |
| IGKV1D-8 | 2.37515435 | -1.2483747 | 4.1742852 | 1.14E-04 | 0.00141684 | 1.12505988 |
| CENPE | 2.37434067 | 2.45209193 | 6.90918978 | 6.85E-09 | 4.24E-07 | 10.2015912 |
| HIST1H3D | 2.37288472 | 5.29458101 | 10.7157989 | 8.83E-15 | 4.74E-12 | 23.434403 |
| HIST1H2AD | 2.36862451 | 4.35040211 | 9.14342604 | 2.05E-12 | 4.64E-10 | 18.0658036 |
| ADAMTS14 | 2.35450672 | -0.4074917 | 4.32499383 | 6.91E-05 | 9.40E-04 | 1.57825832 |
| CDCA8 | 2.34897288 | 1.4672952 | 8.81809836 | 6.54E-12 | 1.20E-09 | 16.9209539 |
| DSCC1 | 2.34099893 | 0.91341021 | 7.77239799 | 2.88E-10 | 2.88E-08 | 13.218642 |
| KIF11 | 2.32859596 | 3.16578095 | 8.79037759 | 7.22E-12 | 1.30E-09 | 16.8644237 |
| C17orf96 | 2.32003351 | -0.1212967 | 7.21041155 | 2.26E-09 | 1.64E-07 | 11.0370764 |
| MTCO2P5 | -2.3087578 | -1.078138 | -4.0286678 | 1.83E-04 | 0.00207668 | 0.70651102 |
| ARHGAP11A | 2.30431407 | 2.69649736 | 7.48875435 | 8.15E-10 | 6.95E-08 | 12.2581868 |
| CHPF | 2.28901302 | 2.05748813 | 7.65707378 | 4.40E-10 | 4.09E-08 | 12.8746008 |
| IGHV4-34 | 2.28064089 | 1.93311898 | 4.48058897 | 4.10E-05 | 6.18E-04 | 1.80862311 |
| GGH | 2.272471 | 0.65043958 | 8.24673365 | 5.13E-11 | 6.60E-09 | 14.8796246 |
| NT5DC2 | 2.2696783 | 2.31561805 | 8.47732047 | 2.23E-11 | 3.43E-09 | 15.7766438 |
| ENTPD2 | -2.2446374 | -1.63185 | -4.9676834 | 7.68E-06 | 1.58E-04 | 3.4449237 |
| KCNN3 | 2.24073848 | 1.27221877 | 8.0761708 | 9.53E-11 | 1.09E-08 | 14.3393722 |
| HIST1H4F | 2.20382387 | 4.58201066 | 6.60124145 | 2.12E-08 | 1.09E-06 | 8.91740795 |
| ARHGEF40 | -2.183299 | 6.31367677 | -7.2424351 | 2.01E-09 | 1.46E-07 | 11.2247675 |
| IGLV3-9 | 2.1721212 | 1.32900873 | 6.25426044 | 7.57E-08 | 3.25E-06 | 7.90142089 |
| RAB3C | -2.1671294 | -0.3896796 | -3.4183415 | 0.00123075 | 0.0093087 | -1.0314143 |
| CABLES1 | 2.16449615 | 0.71797393 | 6.02382163 | 1.75E-07 | 6.70E-06 | 7.13571741 |
| PLK1 | 2.15159624 | 2.74631213 | 8.37008149 | 3.28E-11 | 4.63E-09 | 15.3886754 |
| HIST2H2AB | 2.15109754 | 3.42913951 | 9.52379853 | 5.35E-13 | 1.60E-10 | 19.4096837 |
| FBXO39 | 2.14125716 | 1.93155647 | 4.38121572 | 5.73E-05 | 8.10E-04 | 1.70953275 |
| NUF2 | 2.13477235 | 1.07344 | 6.83046596 | 9.14E-09 | 5.40E-07 | 9.95359853 |
| IGHJ4 | 2.13357534 | 2.3283847 | 5.9025535 | 2.73E-07 | 9.56E-06 | 6.57824811 |
| APOBEC3H | 2.12914645 | 1.52744202 | 7.11709598 | 3.19E-09 | 2.17E-07 | 10.9612274 |
| MCM2 | 2.12791913 | 3.33459092 | 8.30726174 | 4.12E-11 | 5.52E-09 | 15.1438767 |
| OXCT2 | 2.1271415 | -1.0764663 | 4.65197926 | 2.29E-05 | 3.83E-04 | 2.54438708 |
| LMOD1 | -2.1229884 | -0.6258565 | -3.4350361 | 0.00117065 | 0.00895873 | -0.9144088 |
| IFI44 | 2.10642684 | 7.7459889 | 5.07547911 | 5.27E-06 | 1.16E-04 | 3.50126042 |
| MTND6P11 | -2.1028616 | -1.9226973 | -3.6010672 | 7.07E-04 | 0.00600147 | -0.4854871 |
| PSAT1 | 2.08937821 | 1.91429576 | 7.37775171 | 1.22E-09 | 9.83E-08 | 11.8897555 |
| FEN1 | 2.08809922 | 2.90364531 | 8.97538636 | 3.73E-12 | 7.61E-10 | 17.5161108 |
| UBE2C | 2.08695129 | 1.27094373 | 8.26778841 | 4.75E-11 | 6.15E-09 | 15.0128381 |
| ESPL1 | 2.08003232 | 2.05256655 | 7.63544165 | 4.76E-10 | 4.36E-08 | 12.8013051 |
| ALPL | -2.0781419 | 5.98271276 | -3.7751559 | 4.11E-04 | 0.00390314 | -0.7097773 |
| INHBB | -2.0750413 | -1.3086028 | -3.7595955 | 4.32E-04 | 0.00405694 | -0.0531322 |
| HIST1H3H | 2.07343007 | 5.6992742 | 7.77778508 | 2.83E-10 | 2.86E-08 | 13.1695645 |
| LAG3 | 2.07264365 | 2.86273634 | 5.99375966 | 1.96E-07 | 7.32E-06 | 6.93088463 |
| SPATA3 | 2.06988251 | -0.9276438 | 5.48422204 | 1.23E-06 | 3.37E-05 | 5.22331586 |
| IGKJ4 | 2.06901206 | -0.6495209 | 5.20539337 | 3.33E-06 | 7.91E-05 | 4.34670759 |
| PCNA | 2.06461349 | 4.63756007 | 10.0157274 | 9.64E-14 | 3.23E-11 | 21.078808 |
| HIST1H4I | 2.06316064 | 4.44536617 | 9.1150066 | 2.27E-12 | 4.96E-10 | 17.9614398 |
| SCG5 | 2.04278452 | -1.4370596 | 3.91705104 | 2.62E-04 | 0.00274993 | 0.37542124 |
| H2AFX | 2.03949765 | 4.05968015 | 8.54579855 | 1.74E-11 | 2.77E-09 | 15.9609255 |
| LY6E | 2.03355123 | 9.3559693 | 5.45092361 | 1.39E-06 | 3.73E-05 | 4.87116043 |
| IGF1 | 2.02768746 | 1.87364029 | 6.31870589 | 5.98E-08 | 2.69E-06 | 8.12183799 |
| MAD2L1 | 2.02438548 | 2.69007284 | 7.39186065 | 1.16E-09 | 9.45E-08 | 11.8999691 |
| SERPING1 | 2.01393422 | 5.97008766 | 3.86296071 | 3.12E-04 | 0.00313614 | -0.3413551 |

**Supplemental Table 7. Complete IPA analysis, day 0 vs day 6**

| **Ingenuity Canonical Pathways** | **-log(p-value)** | **Ratio** | **z-score** |
| --- | --- | --- | --- |
| Role of Hypercytokinemia/hyperchemokinemia in the Pathogenesis of Influenza | 1.98E+01 | 1.98E-01 | 3.638 |
| Interferon Signaling | 1.71E+01 | 3.33E-01 | 2.887 |
| Activation of IRF by Cytosolic Pattern Recognition Receptors | 9.22E+00 | 1.38E-01 | 1.667 |
| Role of Pattern Recognition Receptors in Recognition of Bacteria and Viruses | 7.97E+00 | 7.05E-02 | 2 |
| Coronavirus Pathogenesis Pathway | 6.80E+00 | 5.42E-02 | -2.111 |
| Role of PKR in Interferon Induction and Antiviral Response | 5.35E+00 | 5.88E-02 | 2.646 |
| Granulocyte Adhesion and Diapedesis | 4.31E+00 | 4.23E-02 | NaN |
| Death Receptor Signaling | 4.29E+00 | 6.25E-02 | 2.449 |
| Retinoic acid Mediated Apoptosis Signaling | 4.25E+00 | 8.33E-02 | 2.236 |
| Agranulocyte Adhesion and Diapedesis | 3.94E+00 | 3.74E-02 | NaN |
| Role of RIG1-like Receptors in Antiviral Innate Immunity | 3.56E+00 | 8.70E-02 | 1 |
| Role of IL-17F in Allergic Inflammatory Airway Diseases | 3.53E+00 | 8.51E-02 | NaN |
| Pyroptosis Signaling Pathway | 3.35E+00 | 5.38E-02 | 2.236 |
| UVA-Induced MAPK Signaling | 3.24E+00 | 5.10E-02 | NaN |
| Role of JAK1, JAK2 and TYK2 in Interferon Signaling | 3.14E+00 | 1.15E-01 | NaN |
| Necroptosis Signaling Pathway | 3.12E+00 | 3.82E-02 | 2.449 |
| HMGB1 Signaling | 2.99E+00 | 3.59E-02 | 1 |
| Airway Pathology in Chronic Obstructive Pulmonary Disease | 2.88E+00 | 4.24E-02 | NaN |
| Role of JAK2 in Hormone-like Cytokine Signaling | 2.80E+00 | 8.82E-02 | NaN |
| Role of MAPK Signaling in Inhibiting the Pathogenesis of Influenza | 2.71E+00 | 5.19E-02 | 1 |
| Hepatic Cholestasis | 2.69E+00 | 3.14E-02 | NaN |
| Coronavirus Replication Pathway | 2.44E+00 | 6.67E-02 | NaN |
| Role of IL-17A in Psoriasis | 2.40E+00 | 1.43E-01 | NaN |
| Role of IL-17A in Arthritis | 2.16E+00 | 5.26E-02 | NaN |
| Tumor Microenvironment Pathway | 2.11E+00 | 2.79E-02 | 1.342 |
| Wound Healing Signaling Pathway | 2.11E+00 | 2.38E-02 | 1.633 |
| Inflammasome pathway | 2.09E+00 | 1.00E-01 | NaN |
| IL-17 Signaling | 2.03E+00 | 2.67E-02 | 1.342 |
| Th1 Pathway | 2.00E+00 | 3.28E-02 | NaN |
| LXR/RXR Activation | 1.99E+00 | 3.25E-02 | 0 |
| Systemic Lupus Erythematosus In B Cell Signaling Pathway | 1.95E+00 | 1.52E-02 | 2.714 |
| IL-6 Signaling | 1.94E+00 | 3.12E-02 | 0 |
| Atherosclerosis Signaling | 1.90E+00 | 3.05E-02 | NaN |
| Role of JAK family kinases in IL-6-type Cytokine Signaling | 1.90E+00 | 8.00E-02 | NaN |
| IL-17A Signaling in Gastric Cells | 1.87E+00 | 7.69E-02 | NaN |
| TREM1 Signaling | 1.80E+00 | 3.90E-02 | NaN |
| JAK/STAT Signaling | 1.73E+00 | 3.66E-02 | NaN |
| NAD Signaling Pathway | 1.69E+00 | 2.65E-02 | 2 |
| Prolactin Signaling | 1.67E+00 | 3.49E-02 | NaN |
| Airway Inflammation in Asthma | 1.67E+00 | 6.06E-02 | NaN |
| Role of Macrophages, Fibroblasts and Endothelial Cells in Rheumatoid Arthritis | 1.59E+00 | 1.83E-02 | NaN |
| Th1 and Th2 Activation Pathway | 1.51E+00 | 2.33E-02 | NaN |
| Erythropoietin Signaling Pathway | 1.47E+00 | 2.26E-02 | -1 |
| Oncostatin M Signaling | 1.46E+00 | 4.65E-02 | NaN |
| Tryptophan Degradation to 2-amino-3-carboxymuconate Semialdehyde | 1.39E+00 | 1.67E-01 | NaN |
| IL-13 Signaling Pathway | 1.34E+00 | 2.59E-02 | NaN |
| Role of Cytokines in Mediating Communication between Immune Cells | 1.28E+00 | 3.70E-02 | NaN |
| FXR/RXR Activation | 1.26E+00 | 2.38E-02 | NaN |
| Pathogenesis of Multiple Sclerosis | 1.22E+00 | 1.11E-01 | NaN |
| MSP-RON Signaling Pathway | 1.22E+00 | 3.45E-02 | NaN |
| Neuroinflammation Signaling Pathway | 1.17E+00 | 1.58E-02 | 1.342 |
| Role of JAK1 and JAK3 in Œ≥c Cytokine Signaling | 1.09E+00 | 2.90E-02 | NaN |
| Agrin Interactions at Neuromuscular Junction | 1.08E+00 | 2.86E-02 | NaN |
| Growth Hormone Signaling | 1.07E+00 | 2.82E-02 | NaN |
| NAD biosynthesis II (from tryptophan) | 1.07E+00 | 7.69E-02 | NaN |
| Guanosine Nucleotides Degradation III | 1.07E+00 | 7.69E-02 | NaN |
| Urate Biosynthesis/Inosine 5'-phosphate Degradation | 1.04E+00 | 7.14E-02 | NaN |
| Toll-like Receptor Signaling | 1.00E+00 | 2.56E-02 | NaN |
| IL-7 Signaling Pathway | 1.00E+00 | 2.56E-02 | NaN |
| Maturity Onset Diabetes of Young (MODY) Signaling | 9.91E-01 | 2.53E-02 | NaN |
| Adenosine Nucleotides Degradation II | 9.83E-01 | 6.25E-02 | NaN |
| FLT3 Signaling in Hematopoietic Progenitor Cells | 9.67E-01 | 2.44E-02 | NaN |
| Role of MAPK Signaling in the Pathogenesis of Influenza | 9.67E-01 | 2.44E-02 | NaN |
| Differential Regulation of Cytokine Production in Macrophages and T Helper Cells by IL-17A and IL-17F | 9.36E-01 | 5.56E-02 | NaN |
| PDGF Signaling | 9.32E-01 | 2.33E-02 | NaN |
| Purine Nucleotides Degradation II (Aerobic) | 9.14E-01 | 5.26E-02 | NaN |
| Acute Myeloid Leukemia Signaling | 8.93E-01 | 2.20E-02 | NaN |
| Crosstalk between Dendritic Cells and Natural Killer Cells | 8.93E-01 | 2.20E-02 | NaN |
| Acute Phase Response Signaling | 8.76E-01 | 1.62E-02 | NaN |
| FcŒ≥ Receptor-mediated Phagocytosis in Macrophages and Monocytes | 8.70E-01 | 2.13E-02 | NaN |
| p53 Signaling | 8.39E-01 | 2.04E-02 | NaN |
| Salvage Pathways of Pyrimidine Ribonucleotides | 8.39E-01 | 2.04E-02 | NaN |
| Clathrin-mediated Endocytosis Signaling | 8.36E-01 | 1.55E-02 | NaN |
| Role of Lipids/Lipid Rafts in the Pathogenesis of Influenza | 8.36E-01 | 4.35E-02 | NaN |
| Differential Regulation of Cytokine Production in Intestinal Epithelial Cells by IL-17A and IL-17F | 8.36E-01 | 4.35E-02 | NaN |
| Pyrimidine Deoxyribonucleotides De Novo Biosynthesis I | 8.36E-01 | 4.35E-02 | NaN |
| Hepatic Fibrosis / Hepatic Stellate Cell Activation | 8.30E-01 | 1.55E-02 | NaN |
| IL-22 Signaling | 8.18E-01 | 4.17E-02 | NaN |
| TCA Cycle II (Eukaryotic) | 8.18E-01 | 4.17E-02 | NaN |
| D-myo-inositol (1,4,5)-Trisphosphate Biosynthesis | 7.88E-01 | 3.85E-02 | NaN |
| Sertoli Cell-Sertoli Cell Junction Signaling | 7.77E-01 | 1.46E-02 | NaN |
| NAD Salvage Pathway II | 7.72E-01 | 3.70E-02 | NaN |
| Tryptophan Degradation III (Eukaryotic) | 7.72E-01 | 3.70E-02 | NaN |
| MSP-RON Signaling In Macrophages Pathway | 7.10E-01 | 1.68E-02 | NaN |
| EIF2 Signaling | 7.03E-01 | 1.34E-02 | NaN |
| Neuroprotective Role of THOP1 in Alzheimer's Disease | 7.03E-01 | 1.67E-02 | NaN |
| p38 MAPK Signaling | 7.03E-01 | 1.67E-02 | NaN |
| Renin-Angiotensin Signaling | 6.93E-01 | 1.64E-02 | NaN |
| IL-15 Production | 6.88E-01 | 1.63E-02 | NaN |
| RHOA Signaling | 6.82E-01 | 1.61E-02 | NaN |
| Inhibition of Angiogenesis by TSP1 | 6.82E-01 | 2.94E-02 | NaN |
| IL-9 Signaling | 6.72E-01 | 2.86E-02 | NaN |
| Pyrimidine Ribonucleotides Interconversion | 6.72E-01 | 2.86E-02 | NaN |
| Osteoarthritis Pathway | 6.60E-01 | 1.27E-02 | NaN |
| Complement System | 6.50E-01 | 2.70E-02 | NaN |
| IL-17A Signaling in Fibroblasts | 6.38E-01 | 2.63E-02 | NaN |
| Pyrimidine Ribonucleotides De Novo Biosynthesis | 6.38E-01 | 2.63E-02 | NaN |
| Notch Signaling | 6.38E-01 | 2.63E-02 | NaN |
| Inhibition of Matrix Metalloproteases | 6.29E-01 | 2.56E-02 | NaN |
| IL-12 Signaling and Production in Macrophages | 6.23E-01 | 1.47E-02 | NaN |
| Mechanisms of Viral Exit from Host Cells | 6.11E-01 | 2.44E-02 | NaN |
| B Cell Activating Factor Signaling | 5.92E-01 | 2.33E-02 | NaN |
| Endocannabinoid Cancer Inhibition Pathway | 5.75E-01 | 1.36E-02 | NaN |
| Axonal Guidance Signaling | 5.70E-01 | 9.82E-03 | NaN |
| Role of OCT4 in Mammalian Embryonic Stem Cell Pluripotency | 5.67E-01 | 2.17E-02 | NaN |
| iNOS Signaling | 5.59E-01 | 2.13E-02 | NaN |
| Insulin Secretion Signaling Pathway | 5.42E-01 | 1.10E-02 | NaN |
| Protein Ubiquitination Pathway | 5.38E-01 | 1.09E-02 | NaN |
| Inhibition of ARE-Mediated mRNA Degradation Pathway | 5.21E-01 | 1.24E-02 | NaN |
| Transcriptional Regulatory Network in Embryonic Stem Cells | 5.09E-01 | 1.85E-02 | NaN |
| EGF Signaling | 5.03E-01 | 1.82E-02 | NaN |
| CSDE1 Signaling Pathway | 4.96E-01 | 1.79E-02 | NaN |
| Sirtuin Signaling Pathway | 4.92E-01 | 1.03E-02 | NaN |
| CNTF Signaling | 4.89E-01 | 1.75E-02 | NaN |
| Germ Cell-Sertoli Cell Junction Signaling | 4.87E-01 | 1.17E-02 | NaN |
| Triacylglycerol Degradation | 4.78E-01 | 1.69E-02 | NaN |
| Senescence Pathway | 4.78E-01 | 1.01E-02 | NaN |
| IL-2 Signaling | 4.66E-01 | 1.64E-02 | NaN |
| Tight Junction Signaling | 4.63E-01 | 1.12E-02 | NaN |
| Thrombopoietin Signaling | 4.55E-01 | 1.59E-02 | NaN |
| Pyridoxal 5'-phosphate Salvage Pathway | 4.39E-01 | 1.52E-02 | NaN |
| Glucocorticoid Receptor Signaling | 4.38E-01 | 8.61E-03 | NaN |
| IL-17A Signaling in Airway Cells | 4.33E-01 | 1.49E-02 | NaN |
| Remodeling of Epithelial Adherens Junctions | 4.28E-01 | 1.47E-02 | NaN |
| Production of Nitric Oxide and Reactive Oxygen Species in Macrophages | 4.25E-01 | 1.05E-02 | NaN |
| SPINK1 General Cancer Pathway | 4.24E-01 | 1.45E-02 | NaN |
| Regulation Of The Epithelial Mesenchymal Transition By Growth Factors Pathway | 4.23E-01 | 1.04E-02 | NaN |
| Leukocyte Extravasation Signaling | 4.19E-01 | 1.04E-02 | NaN |
| GM-CSF Signaling | 4.18E-01 | 1.43E-02 | NaN |
| IL-10 Signaling | 4.09E-01 | 1.39E-02 | NaN |
| Ephrin B Signaling | 4.09E-01 | 1.39E-02 | NaN |
| ID1 Signaling Pathway | 4.00E-01 | 1.00E-02 | NaN |
| ERK5 Signaling | 4.00E-01 | 1.35E-02 | NaN |
| Caveolar-mediated Endocytosis Signaling | 3.96E-01 | 1.33E-02 | NaN |
| Gustation Pathway | 3.93E-01 | 9.85E-03 | NaN |
| Ephrin Receptor Signaling | 3.93E-01 | 9.85E-03 | NaN |
| Hypoxia Signaling in the Cardiovascular System | 3.90E-01 | 1.32E-02 | NaN |
| VDR/RXR Activation | 3.82E-01 | 1.28E-02 | NaN |
| NF-Œ∫B Activation by Viruses | 3.82E-01 | 1.28E-02 | NaN |
| IL-3 Signaling | 3.78E-01 | 1.27E-02 | NaN |
| Thyroid Cancer Signaling | 3.78E-01 | 1.27E-02 | NaN |
| Role of BRCA1 in DNA Damage Response | 3.74E-01 | 1.25E-02 | NaN |
| Chemokine Signaling | 3.74E-01 | 1.25E-02 | NaN |
| Heparan Sulfate Biosynthesis (Late Stages) | 3.70E-01 | 1.23E-02 | NaN |
| BEX2 Signaling Pathway | 3.70E-01 | 1.23E-02 | NaN |
| Role of Osteoblasts, Osteoclasts and Chondrocytes in Rheumatoid Arthritis | 3.43E-01 | 8.93E-03 | NaN |
| Heparan Sulfate Biosynthesis | 3.43E-01 | 1.14E-02 | NaN |
| IL-4 Signaling | 3.26E-01 | 1.08E-02 | NaN |
| TGF-Œ≤ Signaling | 3.16E-01 | 1.04E-02 | NaN |
| VEGF Signaling | 3.07E-01 | 1.01E-02 | NaN |
| Sumoylation Pathway | 2.95E-01 | 9.71E-03 | NaN |
| Cardiac Hypertrophy Signaling (Enhanced) | 2.92E-01 | 7.37E-03 | NaN |
| Virus Entry via Endocytic Pathways | 2.92E-01 | 9.62E-03 | NaN |
| Mouse Embryonic Stem Cell Pluripotency | 2.92E-01 | 9.62E-03 | NaN |
| IGF-1 Signaling | 2.92E-01 | 9.62E-03 | NaN |
| PD-1, PD-L1 cancer immunotherapy pathway | 2.87E-01 | 9.43E-03 | NaN |
| LPS/IL-1 Mediated Inhibition of RXR Function | 2.84E-01 | 7.87E-03 | NaN |
| PPAR Signaling | 2.83E-01 | 9.35E-03 | NaN |
| Paxillin Signaling | 2.81E-01 | 9.26E-03 | NaN |
| T Cell Exhaustion Signaling Pathway | 2.65E-01 | 7.07E-03 | 2 |
| Regulation of Actin-based Motility by Rho | 2.60E-01 | 8.62E-03 | NaN |
| Bladder Cancer Signaling | 2.60E-01 | 8.62E-03 | NaN |
| Role of Tissue Factor in Cancer | 2.60E-01 | 8.62E-03 | NaN |
| Signaling by Rho Family GTPases | 2.60E-01 | 7.46E-03 | NaN |
| Hepatic Fibrosis Signaling Pathway | 2.57E-01 | 7.11E-03 | NaN |
| GPCR-Mediated Nutrient Sensing in Enteroendocrine Cells | 2.55E-01 | 8.47E-03 | NaN |
| Cholecystokinin/Gastrin-mediated Signaling | 2.53E-01 | 8.40E-03 | NaN |
| TEC Kinase Signaling | 2.51E-01 | 6.91E-03 | NaN |
| Role of NANOG in Mammalian Embryonic Stem Cell Pluripotency | 2.50E-01 | 8.33E-03 | NaN |
| Sphingosine-1-phosphate Signaling | 2.50E-01 | 8.33E-03 | NaN |
| Pancreatic Adenocarcinoma Signaling | 2.37E-01 | 7.94E-03 | NaN |
| STAT3 Pathway | 2.18E-01 | 7.41E-03 | NaN |
| Adipogenesis pathway | 2.18E-01 | 7.41E-03 | NaN |
| Iron homeostasis signaling pathway | 2.11E-01 | 7.19E-03 | NaN |
| GŒ±i Signaling | 2.09E-01 | 7.14E-03 | NaN |
| Apelin Endothelial Signaling Pathway | 2.07E-01 | 7.09E-03 | NaN |
| PI3K Signaling in B Lymphocytes | 2.03E-01 | 6.99E-03 | NaN |
| Dilated Cardiomyopathy Signaling Pathway | 0.00E+00 | 6.76E-03 | NaN |
| Oxytocin In Brain Signaling Pathway | 0.00E+00 | 5.08E-03 | NaN |
| Oxytocin Signaling Pathway | 0.00E+00 | 3.55E-03 | NaN |
| Pulmonary Fibrosis Idiopathic Signaling Pathway | 0.00E+00 | 3.07E-03 | NaN |
| CLEAR Signaling Pathway | 0.00E+00 | 3.51E-03 | NaN |
| Natural Killer Cell Signaling | 0.00E+00 | 5.03E-03 | NaN |
| Actin Cytoskeleton Signaling | 0.00E+00 | 4.08E-03 | NaN |
| Huntington's Disease Signaling | 0.00E+00 | 3.53E-03 | NaN |
| NRF2-mediated Oxidative Stress Response | 0.00E+00 | 4.22E-03 | NaN |
| PPARŒ±/RXRŒ± Activation | 0.00E+00 | 5.13E-03 | NaN |
| Mitochondrial Dysfunction | 0.00E+00 | 5.85E-03 | NaN |
| RAR Activation | 0.00E+00 | 4.90E-03 | NaN |
| IL-8 Signaling | 0.00E+00 | 4.76E-03 | NaN |
| Role of NFAT in Regulation of the Immune Response | 0.00E+00 | 1.70E-03 | NaN |
| T Helper Cell Differentiation | 0.00E+00 | 2.12E-03 | NaN |
| IL-15 Signaling | 0.00E+00 | 1.89E-03 | NaN |
| Dendritic Cell Maturation | 0.00E+00 | 6.76E-03 | 1 |
| Cellular Effects of Sildenafil (Viagra) | 0.00E+00 | 6.67E-03 | NaN |
| Endothelin-1 Signaling | 0.00E+00 | 5.21E-03 | NaN |
| CREB Signaling in Neurons | 0.00E+00 | 1.65E-03 | NaN |
| Type I Diabetes Mellitus Signaling | 0.00E+00 | 3.93E-03 | NaN |
| Graft-versus-Host Disease Signaling | 0.00E+00 | 2.24E-03 | NaN |
| Type II Diabetes Mellitus Signaling | 0.00E+00 | 6.54E-03 | NaN |
| Colorectal Cancer Metastasis Signaling | 0.00E+00 | 3.69E-03 | NaN |
| Communication between Innate and Adaptive Immune Cells | 0.00E+00 | 4.31E-03 | NaN |
| Systemic Lupus Erythematosus Signaling | 0.00E+00 | 4.82E-03 | NaN |
| ILK Signaling | 0.00E+00 | 5.00E-03 | NaN |
| FAK Signaling | 0.00E+00 | 1.92E-03 | NaN |
| Altered T Cell and B Cell Signaling in Rheumatoid Arthritis | 0.00E+00 | 4.07E-03 | NaN |
| Protein Kinase A Signaling | 0.00E+00 | 2.44E-03 | NaN |
| Role of NFAT in Cardiac Hypertrophy | 0.00E+00 | 4.50E-03 | NaN |
| Breast Cancer Regulation by Stathmin1 | 0.00E+00 | 1.68E-03 | NaN |
| NUR77 Signaling in T Lymphocytes | 0.00E+00 | 1.95E-03 | NaN |
| RHOGDI Signaling | 0.00E+00 | 4.65E-03 | NaN |
| Gap Junction Signaling | 0.00E+00 | 5.05E-03 | NaN |
| Hematopoiesis from Pluripotent Stem Cells | 0.00E+00 | 4.52E-03 | NaN |
| Superpathway of Inositol Phosphate Compounds | 0.00E+00 | 4.22E-03 | NaN |
| 3-phosphoinositide Biosynthesis | 0.00E+00 | 4.81E-03 | NaN |
| Sperm Motility | 0.00E+00 | 3.92E-03 | NaN |
| Estrogen Receptor Signaling | 0.00E+00 | 2.44E-03 | NaN |
| ERK/MAPK Signaling | 0.00E+00 | 4.63E-03 | NaN |
| Integrin Signaling | 0.00E+00 | 4.69E-03 | NaN |
| cAMP-mediated signaling | 0.00E+00 | 4.26E-03 | NaN |
| NF-Œ∫B Signaling | 0.00E+00 | 5.26E-03 | NaN |
| G-Protein Coupled Receptor Signaling | 0.00E+00 | 1.42E-03 | NaN |
| Phagosome Formation | 0.00E+00 | 2.89E-03 | NaN |
| Opioid Signaling Pathway | 0.00E+00 | 3.57E-03 | NaN |
| Adrenomedullin signaling pathway | 0.00E+00 | 5.03E-03 | NaN |
| Synaptogenesis Signaling Pathway | 0.00E+00 | 3.15E-03 | NaN |
| Systemic Lupus Erythematosus In T Cell Signaling Pathway | 0.00E+00 | 1.56E-03 | NaN |

**Supplemental Table 8. Complete IPA analysis, day 0 vs day 8**

| **Ingenuity Canonical Pathways** | **-log(p-value)** | **Ratio** | **z-score** |
| --- | --- | --- | --- |
| Role of Hypercytokinemia/hyperchemokinemia in the Pathogenesis of Influenza | 1.70E+01 | 1.98E-01 | 4.123 |
| Interferon Signaling | 1.35E+01 | 3.06E-01 | 2.714 |
| Role of Pattern Recognition Receptors in Recognition of Bacteria and Viruses | 1.03E+01 | 9.62E-02 | 2.828 |
| Activation of IRF by Cytosolic Pattern Recognition Receptors | 7.81E+00 | 1.38E-01 | 1.667 |
| Granulocyte Adhesion and Diapedesis | 6.36E+00 | 6.35E-02 | NaN |
| Coronavirus Pathogenesis Pathway | 6.03E+00 | 5.91E-02 | -2.111 |
| Complement System | 4.51E+00 | 1.35E-01 | 1 |
| Agranulocyte Adhesion and Diapedesis | 4.24E+00 | 4.67E-02 | NaN |
| Role of PKR in Interferon Induction and Antiviral Response | 4.19E+00 | 5.88E-02 | 2.449 |
| Pathogenesis of Multiple Sclerosis | 4.11E+00 | 3.33E-01 | NaN |
| Necroptosis Signaling Pathway | 3.75E+00 | 5.10E-02 | 2.828 |
| Role of RIG1-like Receptors in Antiviral Innate Immunity | 2.96E+00 | 8.70E-02 | 1 |
| JAK/STAT Signaling | 2.88E+00 | 6.10E-02 | 0.447 |
| Tumor Microenvironment Pathway | 2.68E+00 | 3.91E-02 | 1.89 |
| Role of JAK1, JAK2 and TYK2 in Interferon Signaling | 2.68E+00 | 1.15E-01 | NaN |
| Pyroptosis Signaling Pathway | 2.63E+00 | 5.38E-02 | 2.236 |
| Acute Phase Response Signaling | 2.60E+00 | 3.78E-02 | NaN |
| UVA-Induced MAPK Signaling | 2.53E+00 | 5.10E-02 | NaN |
| Retinoic acid Mediated Apoptosis Signaling | 2.53E+00 | 6.67E-02 | 2 |
| Role of JAK2 in Hormone-like Cytokine Signaling | 2.34E+00 | 8.82E-02 | NaN |
| HMGB1 Signaling | 2.19E+00 | 3.59E-02 | 2 |
| Oncostatin M Signaling | 2.06E+00 | 6.98E-02 | NaN |
| Coronavirus Replication Pathway | 2.00E+00 | 6.67E-02 | NaN |
| Prolactin Signaling | 1.98E+00 | 4.65E-02 | 1 |
| Role of IL-17F in Allergic Inflammatory Airway Diseases | 1.95E+00 | 6.38E-02 | NaN |
| Acute Myeloid Leukemia Signaling | 1.89E+00 | 4.40E-02 | NaN |
| Death Receptor Signaling | 1.82E+00 | 4.17E-02 | 2 |
| Inflammasome pathway | 1.78E+00 | 1.00E-01 | NaN |
| Role of JAK family kinases in IL-6-type Cytokine Signaling | 1.60E+00 | 8.00E-02 | NaN |
| IL-17A Signaling in Gastric Cells | 1.57E+00 | 7.69E-02 | NaN |
| Estrogen-mediated S-phase Entry | 1.57E+00 | 7.69E-02 | NaN |
| IL-13 Signaling Pathway | 1.55E+00 | 3.45E-02 | -1 |
| Systemic Lupus Erythematosus In B Cell Signaling Pathway | 1.55E+00 | 1.80E-02 | 3.051 |
| Th1 and Th2 Activation Pathway | 1.55E+00 | 2.91E-02 | NaN |
| Tetrahydrobiopterin Biosynthesis I | 1.53E+00 | 3.33E-01 | NaN |
| Glycerol-3-phosphate Shuttle | 1.53E+00 | 3.33E-01 | NaN |
| Tetrahydrobiopterin Biosynthesis II | 1.53E+00 | 3.33E-01 | NaN |
| Airway Pathology in Chronic Obstructive Pulmonary Disease | 1.52E+00 | 3.39E-02 | NaN |
| Role of JAK1 and JAK3 in Œ≥c Cytokine Signaling | 1.51E+00 | 4.35E-02 | NaN |
| Th1 Pathway | 1.48E+00 | 3.28E-02 | NaN |
| IL-15 Production | 1.47E+00 | 3.25E-02 | 2 |
| IL-10 Signaling | 1.46E+00 | 4.17E-02 | NaN |
| IL-6 Signaling | 1.41E+00 | 3.12E-02 | 1 |
| IL-17 Signaling | 1.41E+00 | 2.67E-02 | 2.236 |
| Heme Degradation | 1.41E+00 | 2.50E-01 | NaN |
| Melatonin Degradation II | 1.41E+00 | 2.50E-01 | NaN |
| Wound Healing Signaling Pathway | 1.40E+00 | 2.38E-02 | 2.449 |
| TREM1 Signaling | 1.39E+00 | 3.90E-02 | NaN |
| Role of MAPK Signaling in Inhibiting the Pathogenesis of Influenza | 1.39E+00 | 3.90E-02 | NaN |
| Hepatic Cholestasis | 1.38E+00 | 2.62E-02 | NaN |
| Airway Inflammation in Asthma | 1.38E+00 | 6.06E-02 | NaN |
| Toll-like Receptor Signaling | 1.37E+00 | 3.85E-02 | NaN |
| Ferroptosis Signaling Pathway | 1.37E+00 | 3.03E-02 | -1 |
| Pyrimidine Ribonucleotides Interconversion | 1.33E+00 | 5.71E-02 | NaN |
| FLT3 Signaling in Hematopoietic Progenitor Cells | 1.32E+00 | 3.66E-02 | NaN |
| Role of MAPK Signaling in the Pathogenesis of Influenza | 1.32E+00 | 3.66E-02 | NaN |
| ID1 Signaling Pathway | 1.31E+00 | 2.50E-02 | -0.447 |
| Glutathione-mediated Detoxification | 1.29E+00 | 5.41E-02 | NaN |
| PDGF Signaling | 1.27E+00 | 3.49E-02 | NaN |
| Pyrimidine Ribonucleotides De Novo Biosynthesis | 1.27E+00 | 5.26E-02 | NaN |
| Inhibition of Matrix Metalloproteases | 1.25E+00 | 5.13E-02 | NaN |
| Glycerol Degradation I | 1.24E+00 | 1.67E-01 | NaN |
| Tryptophan Degradation to 2-amino-3-carboxymuconate Semialdehyde | 1.24E+00 | 1.67E-01 | NaN |
| TGF-Œ≤ Signaling | 1.15E+00 | 3.12E-02 | NaN |
| Serotonin Receptor Signaling | 1.14E+00 | 4.44E-02 | NaN |
| p53 Signaling | 1.13E+00 | 3.06E-02 | NaN |
| Melanoma Signaling | 1.06E+00 | 4.00E-02 | NaN |
| Erythropoietin Signaling Pathway | 1.00E+00 | 2.26E-02 | -1 |
| Transcriptional Regulatory Network in Embryonic Stem Cells | 1.00E+00 | 3.70E-02 | NaN |
| Bladder Cancer Signaling | 9.67E-01 | 2.59E-02 | NaN |
| Role of Macrophages, Fibroblasts and Endothelial Cells in Rheumatoid Arthritis | 9.67E-01 | 1.83E-02 | NaN |
| CNTF Signaling | 9.63E-01 | 3.51E-02 | NaN |
| Role of IL-17A in Arthritis | 9.63E-01 | 3.51E-02 | NaN |
| MSP-RON Signaling In Macrophages Pathway | 9.43E-01 | 2.52E-02 | NaN |
| Triacylglycerol Degradation | 9.36E-01 | 3.39E-02 | NaN |
| Renin-Angiotensin Signaling | 9.17E-01 | 2.46E-02 | NaN |
| NAD biosynthesis II (from tryptophan) | 9.17E-01 | 7.69E-02 | NaN |
| IL-2 Signaling | 9.14E-01 | 3.28E-02 | NaN |
| Thrombopoietin Signaling | 8.89E-01 | 3.17E-02 | NaN |
| Role of IL-17A in Psoriasis | 8.86E-01 | 7.14E-02 | NaN |
| Phenylalanine Degradation IV (Mammalian, via Side Chain) | 8.86E-01 | 7.14E-02 | NaN |
| Atherosclerosis Signaling | 8.51E-01 | 2.29E-02 | NaN |
| Gustation Pathway | 8.45E-01 | 1.97E-02 | 1 |
| Ephrin Receptor Signaling | 8.45E-01 | 1.97E-02 | NaN |
| Sertoli Cell-Sertoli Cell Junction Signaling | 8.30E-01 | 1.94E-02 | NaN |
| Phagosome Formation | 8.27E-01 | 1.45E-02 | 2.53 |
| SPINK1 General Cancer Pathway | 8.27E-01 | 2.90E-02 | NaN |
| STAT3 Pathway | 8.24E-01 | 2.22E-02 | NaN |
| Agrin Interactions at Neuromuscular Junction | 8.18E-01 | 2.86E-02 | NaN |
| 3-phosphoinositide Biosynthesis | 8.18E-01 | 1.92E-02 | 1 |
| GM-CSF Signaling | 8.18E-01 | 2.86E-02 | NaN |
| Growth Hormone Signaling | 8.07E-01 | 2.82E-02 | NaN |
| Differential Regulation of Cytokine Production in Macrophages and T Helper Cells by IL-17A and IL-17F | 7.85E-01 | 5.56E-02 | NaN |
| Apelin Endothelial Signaling Pathway | 7.85E-01 | 2.13E-02 | NaN |
| ERK5 Signaling | 7.80E-01 | 2.70E-02 | NaN |
| Angiopoietin Signaling | 7.52E-01 | 2.60E-02 | NaN |
| Antiproliferative Role of Somatostatin Receptor 2 | 7.52E-01 | 2.60E-02 | NaN |
| Endocannabinoid Cancer Inhibition Pathway | 7.47E-01 | 2.04E-02 | NaN |
| VDR/RXR Activation | 7.45E-01 | 2.56E-02 | NaN |
| NF-Œ∫B Activation by Viruses | 7.45E-01 | 2.56E-02 | NaN |
| IL-7 Signaling Pathway | 7.45E-01 | 2.56E-02 | NaN |
| IL-3 Signaling | 7.35E-01 | 2.53E-02 | NaN |
| Dopamine Receptor Signaling | 7.35E-01 | 2.53E-02 | NaN |
| Role of BRCA1 in DNA Damage Response | 7.26E-01 | 2.50E-02 | NaN |
| Chemokine Signaling | 7.26E-01 | 2.50E-02 | NaN |
| NAD Signaling Pathway | 7.24E-01 | 1.99E-02 | NaN |
| Putrescine Degradation III | 7.06E-01 | 4.55E-02 | NaN |
| Cyclins and Cell Cycle Regulation | 6.95E-01 | 2.38E-02 | NaN |
| Role of Lipids/Lipid Rafts in the Pathogenesis of Influenza | 6.90E-01 | 4.35E-02 | NaN |
| Differential Regulation of Cytokine Production in Intestinal Epithelial Cells by IL-17A and IL-17F | 6.90E-01 | 4.35E-02 | NaN |
| Pyrimidine Deoxyribonucleotides De Novo Biosynthesis I | 6.90E-01 | 4.35E-02 | NaN |
| Neuroinflammation Signaling Pathway | 6.86E-01 | 1.58E-02 | 2.236 |
| Superpathway of Inositol Phosphate Compounds | 6.82E-01 | 1.69E-02 | 1 |
| IL-22 Signaling | 6.74E-01 | 4.17E-02 | NaN |
| TCA Cycle II (Eukaryotic) | 6.74E-01 | 4.17E-02 | NaN |
| BMP signaling pathway | 6.72E-01 | 2.30E-02 | NaN |
| Regulation of Cellular Mechanics by Calpain Protease | 6.58E-01 | 2.25E-02 | NaN |
| Crosstalk between Dendritic Cells and Natural Killer Cells | 6.44E-01 | 2.20E-02 | NaN |
| D-myo-inositol (1,4,5)-Trisphosphate Biosynthesis | 6.42E-01 | 3.85E-02 | NaN |
| IL-4 Signaling | 6.29E-01 | 2.15E-02 | NaN |
| Breast Cancer Regulation by Stathmin1 | 6.27E-01 | 1.35E-02 | 2.121 |
| Tryptophan Degradation X (Mammalian, via Tryptamine) | 6.27E-01 | 3.70E-02 | NaN |
| Tryptophan Degradation III (Eukaryotic) | 6.27E-01 | 3.70E-02 | NaN |
| Melanocyte Development and Pigmentation Signaling | 5.97E-01 | 2.04E-02 | NaN |
| CREB Signaling in Neurons | 5.97E-01 | 1.32E-02 | 2.121 |
| Salvage Pathways of Pyrimidine Ribonucleotides | 5.97E-01 | 2.04E-02 | NaN |
| D-myo-inositol (1,4,5,6)-Tetrakisphosphate Biosynthesis | 5.70E-01 | 1.65E-02 | NaN |
| D-myo-inositol (3,4,5,6)-tetrakisphosphate Biosynthesis | 5.70E-01 | 1.65E-02 | NaN |
| Dopamine Degradation | 5.64E-01 | 3.12E-02 | NaN |
| Mouse Embryonic Stem Cell Pluripotency | 5.61E-01 | 1.92E-02 | NaN |
| IGF-1 Signaling | 5.61E-01 | 1.92E-02 | NaN |
| Colorectal Cancer Metastasis Signaling | 5.51E-01 | 1.48E-02 | 1 |
| PD-1, PD-L1 cancer immunotherapy pathway | 5.50E-01 | 1.89E-02 | NaN |
| Chronic Myeloid Leukemia Signaling | 5.44E-01 | 1.87E-02 | NaN |
| Telomerase Signaling | 5.44E-01 | 1.87E-02 | NaN |
| PPAR Signaling | 5.44E-01 | 1.87E-02 | NaN |
| Inhibition of Angiogenesis by TSP1 | 5.42E-01 | 2.94E-02 | NaN |
| IL-9 Signaling | 5.32E-01 | 2.86E-02 | NaN |
| Regulation Of The Epithelial Mesenchymal Transition By Growth Factors Pathway | 5.29E-01 | 1.56E-02 | NaN |
| Leukocyte Extravasation Signaling | 5.24E-01 | 1.55E-02 | NaN |
| 3-phosphoinositide Degradation | 5.21E-01 | 1.55E-02 | NaN |
| PPARŒ±/RXRŒ± Activation | 5.17E-01 | 1.54E-02 | NaN |
| Noradrenaline and Adrenaline Degradation | 5.11E-01 | 2.70E-02 | NaN |
| Prostate Cancer Signaling | 5.07E-01 | 1.75E-02 | NaN |
| D-myo-inositol-5-phosphate Metabolism | 5.06E-01 | 1.52E-02 | NaN |
| Docosahexaenoic Acid (DHA) Signaling | 5.02E-01 | 2.63E-02 | NaN |
| IL-17A Signaling in Fibroblasts | 5.02E-01 | 2.63E-02 | NaN |
| Notch Signaling | 5.02E-01 | 2.63E-02 | NaN |
| Cholecystokinin/Gastrin-mediated Signaling | 4.81E-01 | 1.68E-02 | NaN |
| Role of NANOG in Mammalian Embryonic Stem Cell Pluripotency | 4.78E-01 | 1.67E-02 | NaN |
| Neuroprotective Role of THOP1 in Alzheimer's Disease | 4.78E-01 | 1.67E-02 | NaN |
| p38 MAPK Signaling | 4.78E-01 | 1.67E-02 | NaN |
| HIF1Œ± Signaling | 4.70E-01 | 1.44E-02 | NaN |
| Senescence Pathway | 4.67E-01 | 1.34E-02 | 1 |
| LXR/RXR Activation | 4.63E-01 | 1.63E-02 | NaN |
| B Cell Activating Factor Signaling | 4.58E-01 | 2.33E-02 | NaN |
| Glioma Signaling | 4.58E-01 | 1.61E-02 | NaN |
| FXR/RXR Activation | 4.50E-01 | 1.59E-02 | NaN |
| Pancreatic Adenocarcinoma Signaling | 4.50E-01 | 1.59E-02 | NaN |
| Role of OCT4 in Mammalian Embryonic Stem Cell Pluripotency | 4.35E-01 | 2.17E-02 | NaN |
| iNOS Signaling | 4.27E-01 | 2.13E-02 | NaN |
| Retinol Biosynthesis | 4.27E-01 | 2.13E-02 | NaN |
| HGF Signaling | 4.25E-01 | 1.52E-02 | NaN |
| Protein Kinase A Signaling | 4.20E-01 | 1.22E-02 | -2 |
| P2Y Purigenic Receptor Signaling Pathway | 4.20E-01 | 1.50E-02 | NaN |
| EIF2 Signaling | 4.18E-01 | 1.34E-02 | NaN |
| Role of Osteoblasts, Osteoclasts and Chondrocytes in Rheumatoid Arthritis | 4.18E-01 | 1.34E-02 | NaN |
| Axonal Guidance Signaling | 4.09E-01 | 1.18E-02 | NaN |
| IL-12 Signaling and Production in Macrophages | 4.09E-01 | 1.47E-02 | NaN |
| Cell Cycle: G2/M DNA Damage Checkpoint Regulation | 4.07E-01 | 2.00E-02 | NaN |
| G-Protein Coupled Receptor Signaling | 4.05E-01 | 1.14E-02 | 2.121 |
| UVC-Induced MAPK Signaling | 4.00E-01 | 1.96E-02 | NaN |
| Hereditary Breast Cancer Signaling | 3.86E-01 | 1.41E-02 | NaN |
| PI3K Signaling in B Lymphocytes | 3.83E-01 | 1.40E-02 | NaN |
| Osteoarthritis Pathway | 3.83E-01 | 1.27E-02 | NaN |
| Role of Cytokines in Mediating Communication between Immune Cells | 3.81E-01 | 1.85E-02 | NaN |
| EGF Signaling | 3.75E-01 | 1.82E-02 | NaN |
| CSDE1 Signaling Pathway | 3.70E-01 | 1.79E-02 | NaN |
| Role of CHK Proteins in Cell Cycle Checkpoint Control | 3.64E-01 | 1.75E-02 | NaN |
| PTEN Signaling | 3.59E-01 | 1.33E-02 | NaN |
| Polyamine Regulation in Colon Cancer | 3.58E-01 | 1.72E-02 | NaN |
| MSP-RON Signaling Pathway | 3.58E-01 | 1.72E-02 | NaN |
| Cancer Drug Resistance By Drug Efflux | 3.58E-01 | 1.72E-02 | NaN |
| Endometrial Cancer Signaling | 3.47E-01 | 1.67E-02 | NaN |
| GADD45 Signaling | 3.47E-01 | 1.67E-02 | NaN |
| LPS/IL-1 Mediated Inhibition of RXR Function | 3.36E-01 | 1.18E-02 | NaN |
| Sperm Motility | 3.34E-01 | 1.18E-02 | NaN |
| Aryl Hydrocarbon Receptor Signaling | 3.30E-01 | 1.26E-02 | NaN |
| Inhibition of ARE-Mediated mRNA Degradation Pathway | 3.24E-01 | 1.24E-02 | NaN |
| ERB2-ERBB3 Signaling | 3.22E-01 | 1.54E-02 | NaN |
| HOTAIR Regulatory Pathway | 3.18E-01 | 1.23E-02 | NaN |
| Pyridoxal 5'-phosphate Salvage Pathway | 3.17E-01 | 1.52E-02 | NaN |
| IL-17A Signaling in Airway Cells | 3.12E-01 | 1.49E-02 | NaN |
| Superpathway of Melatonin Degradation | 3.12E-01 | 1.49E-02 | NaN |
| T Cell Exhaustion Signaling Pathway | 3.10E-01 | 1.06E-02 | 2.236 |
| ERBB4 Signaling | 3.08E-01 | 1.47E-02 | NaN |
| Cell Cycle: G1/S Checkpoint Regulation | 3.08E-01 | 1.47E-02 | NaN |
| Eicosanoid Signaling | 3.08E-01 | 1.47E-02 | NaN |
| Mitochondrial Dysfunction | 2.96E-01 | 1.17E-02 | NaN |
| Germ Cell-Sertoli Cell Junction Signaling | 2.96E-01 | 1.17E-02 | NaN |
| Glioblastoma Multiforme Signaling | 2.96E-01 | 1.17E-02 | NaN |
| Serotonin Degradation | 2.95E-01 | 1.41E-02 | NaN |
| Insulin Secretion Signaling Pathway | 2.94E-01 | 1.10E-02 | NaN |
| Ephrin B Signaling | 2.91E-01 | 1.39E-02 | NaN |
| Protein Ubiquitination Pathway | 2.90E-01 | 1.09E-02 | NaN |
| Glucocorticoid Receptor Signaling | 2.88E-01 | 1.03E-02 | NaN |
| Glioma Invasiveness Signaling | 2.87E-01 | 1.37E-02 | NaN |
| Tight Junction Signaling | 2.78E-01 | 1.12E-02 | NaN |
| GDNF Family Ligand-Receptor Interactions | 2.75E-01 | 1.32E-02 | NaN |
| Hypoxia Signaling in the Cardiovascular System | 2.75E-01 | 1.32E-02 | NaN |
| Macropinocytosis Signaling | 2.75E-01 | 1.32E-02 | NaN |
| CLEAR Signaling Pathway | 2.70E-01 | 1.05E-02 | NaN |
| Neurotrophin/TRK Signaling | 2.68E-01 | 1.28E-02 | NaN |
| Maturity Onset Diabetes of Young (MODY) Signaling | 2.64E-01 | 1.27E-02 | NaN |
| Thyroid Cancer Signaling | 2.64E-01 | 1.27E-02 | NaN |
| Renal Cell Carcinoma Signaling | 2.60E-01 | 1.25E-02 | NaN |
| Xenobiotic Metabolism Signaling | 2.60E-01 | 1.03E-02 | NaN |
| Estrogen-Dependent Breast Cancer Signaling | 2.56E-01 | 1.23E-02 | NaN |
| Heparan Sulfate Biosynthesis (Late Stages) | 2.56E-01 | 1.23E-02 | NaN |
| BEX2 Signaling Pathway | 2.56E-01 | 1.23E-02 | NaN |
| VEGF Family Ligand-Receptor Interactions | 2.46E-01 | 1.19E-02 | NaN |
| PEDF Signaling | 2.46E-01 | 1.19E-02 | NaN |
| BAG2 Signaling Pathway | 2.46E-01 | 1.19E-02 | NaN |
| Endothelin-1 Signaling | 2.46E-01 | 1.04E-02 | NaN |
| FcŒ≥RIIB Signaling in B Lymphocytes | 2.43E-01 | 1.18E-02 | NaN |
| LPS-stimulated MAPK Signaling | 2.43E-01 | 1.18E-02 | NaN |
| Xenobiotic Metabolism PXR Signaling Pathway | 2.43E-01 | 1.04E-02 | NaN |
| Hepatic Fibrosis / Hepatic Stellate Cell Activation | 2.41E-01 | 1.03E-02 | NaN |
| Oxytocin In Brain Signaling Pathway | 2.35E-01 | 1.02E-02 | NaN |
| Heparan Sulfate Biosynthesis | 2.34E-01 | 1.14E-02 | NaN |
| Pulmonary Healing Signaling Pathway | 2.31E-01 | 1.01E-02 | NaN |
| Natural Killer Cell Signaling | 2.31E-01 | 1.01E-02 | NaN |
| PI3K/AKT Signaling | 2.31E-01 | 1.01E-02 | NaN |
| Adrenomedullin signaling pathway | 2.31E-01 | 1.01E-02 | NaN |
| Ceramide Signaling | 2.28E-01 | 1.11E-02 | NaN |
| Actin Nucleation by ARP-WASP Complex | 2.19E-01 | 1.08E-02 | NaN |
| Non-Small Cell Lung Cancer Signaling | 2.16E-01 | 1.06E-02 | NaN |
| ERBB Signaling | 2.16E-01 | 1.06E-02 | NaN |
| ATM Signaling | 2.03E-01 | 1.01E-02 | NaN |
| VEGF Signaling | 2.03E-01 | 1.01E-02 | NaN |
| Neurovascular Coupling Signaling Pathway | 0.00E+00 | 4.44E-03 | NaN |
| Oxytocin Signaling Pathway | 0.00E+00 | 3.55E-03 | NaN |
| Pulmonary Fibrosis Idiopathic Signaling Pathway | 0.00E+00 | 9.20E-03 | NaN |
| Neuregulin Signaling | 0.00E+00 | 8.55E-03 | NaN |
| Circadian Rhythm Signaling | 0.00E+00 | 3.72E-03 | NaN |
| Synaptic Long Term Potentiation | 0.00E+00 | 7.63E-03 | NaN |
| Fc Epsilon RI Signaling | 0.00E+00 | 8.47E-03 | NaN |
| Actin Cytoskeleton Signaling | 0.00E+00 | 4.08E-03 | NaN |
| Synaptic Long Term Depression | 0.00E+00 | 5.13E-03 | NaN |
| Huntington's Disease Signaling | 0.00E+00 | 3.53E-03 | NaN |
| NRF2-mediated Oxidative Stress Response | 0.00E+00 | 4.22E-03 | NaN |
| RAR Activation | 0.00E+00 | 4.90E-03 | NaN |
| 14-3-3-mediated Signaling | 0.00E+00 | 7.87E-03 | NaN |
| Œ±-Adrenergic Signaling | 0.00E+00 | 9.26E-03 | NaN |
| Clathrin-mediated Endocytosis Signaling | 0.00E+00 | 5.18E-03 | NaN |
| IL-8 Signaling | 0.00E+00 | 9.52E-03 | NaN |
| Role of NFAT in Regulation of the Immune Response | 0.00E+00 | 1.70E-03 | NaN |
| fMLP Signaling in Neutrophils | 0.00E+00 | 7.63E-03 | NaN |
| CXCR4 Signaling | 0.00E+00 | 5.92E-03 | NaN |
| T Helper Cell Differentiation | 0.00E+00 | 2.12E-03 | NaN |
| CCR3 Signaling in Eosinophils | 0.00E+00 | 7.41E-03 | NaN |
| IL-15 Signaling | 0.00E+00 | 1.89E-03 | NaN |
| Virus Entry via Endocytic Pathways | 0.00E+00 | 9.62E-03 | NaN |
| Dendritic Cell Maturation | 0.00E+00 | 6.76E-03 | 0 |
| Factors Promoting Cardiogenesis in Vertebrates | 0.00E+00 | 6.54E-03 | NaN |
| Thrombin Signaling | 0.00E+00 | 4.44E-03 | NaN |
| Cardiac Hypertrophy Signaling | 0.00E+00 | 3.83E-03 | NaN |
| CDK5 Signaling | 0.00E+00 | 8.77E-03 | NaN |
| Molecular Mechanisms of Cancer | 0.00E+00 | 4.47E-03 | NaN |
| Lipid Antigen Presentation by CD1 | 0.00E+00 | 2.40E-03 | NaN |
| GNRH Signaling | 0.00E+00 | 5.21E-03 | NaN |
| Human Embryonic Stem Cell Pluripotency | 0.00E+00 | 6.02E-03 | NaN |
| Type I Diabetes Mellitus Signaling | 0.00E+00 | 3.93E-03 | NaN |
| Graft-versus-Host Disease Signaling | 0.00E+00 | 2.24E-03 | NaN |
| Type II Diabetes Mellitus Signaling | 0.00E+00 | 6.54E-03 | NaN |
| Production of Nitric Oxide and Reactive Oxygen Species in Macrophages | 0.00E+00 | 5.24E-03 | NaN |
| GŒ±12/13 Signaling | 0.00E+00 | 7.52E-03 | NaN |
| p70S6K Signaling | 0.00E+00 | 7.58E-03 | NaN |
| mTOR Signaling | 0.00E+00 | 4.72E-03 | NaN |
| G Beta Gamma Signaling | 0.00E+00 | 7.75E-03 | NaN |
| Communication between Innate and Adaptive Immune Cells | 0.00E+00 | 4.31E-03 | NaN |
| Sphingosine-1-phosphate Signaling | 0.00E+00 | 8.33E-03 | NaN |
| Systemic Lupus Erythematosus Signaling | 0.00E+00 | 6.43E-03 | NaN |
| FAK Signaling | 0.00E+00 | 8.65E-03 | 1.667 |
| AMPK Signaling | 0.00E+00 | 4.10E-03 | NaN |
| PAK Signaling | 0.00E+00 | 8.47E-03 | NaN |
| RAC Signaling | 0.00E+00 | 7.25E-03 | NaN |
| RHOA Signaling | 0.00E+00 | 8.06E-03 | NaN |
| Phospholipase C Signaling | 0.00E+00 | 1.47E-03 | NaN |
| Ovarian Cancer Signaling | 0.00E+00 | 6.33E-03 | NaN |
| HER-2 Signaling in Breast Cancer | 0.00E+00 | 8.81E-03 | NaN |
| Altered T Cell and B Cell Signaling in Rheumatoid Arthritis | 0.00E+00 | 6.10E-03 | NaN |
| Regulation of eIF4 and p70S6K Signaling | 0.00E+00 | 5.59E-03 | NaN |
| Role of NFAT in Cardiac Hypertrophy | 0.00E+00 | 9.01E-03 | NaN |
| Regulation of IL-2 Expression in Activated and Anergic T Lymphocytes | 0.00E+00 | 2.17E-03 | NaN |
| NUR77 Signaling in T Lymphocytes | 0.00E+00 | 1.95E-03 | NaN |
| PKCŒ∏ Signaling in T Lymphocytes | 0.00E+00 | 1.79E-03 | NaN |
| Antiproliferative Role of TOB in T Cell Signaling | 0.00E+00 | 2.35E-03 | NaN |
| Role of Tissue Factor in Cancer | 0.00E+00 | 8.62E-03 | NaN |
| NGF Signaling | 0.00E+00 | 8.33E-03 | NaN |
| Paxillin Signaling | 0.00E+00 | 9.26E-03 | NaN |
| Signaling by Rho Family GTPases | 0.00E+00 | 3.73E-03 | NaN |
| Gap Junction Signaling | 0.00E+00 | 5.05E-03 | NaN |
| Hematopoiesis from Pluripotent Stem Cells | 0.00E+00 | 2.26E-03 | NaN |
| eNOS Signaling | 0.00E+00 | 6.29E-03 | NaN |
| Epithelial Adherens Junction Signaling | 0.00E+00 | 6.37E-03 | NaN |
| GŒ±i Signaling | 0.00E+00 | 7.14E-03 | NaN |
| Regulation of the Epithelial-Mesenchymal Transition Pathway | 0.00E+00 | 5.13E-03 | NaN |
| TEC Kinase Signaling | 0.00E+00 | 5.18E-03 | NaN |
| Adipogenesis pathway | 0.00E+00 | 7.41E-03 | NaN |
| Estrogen Receptor Signaling | 0.00E+00 | 9.78E-03 | 0 |
| ERK/MAPK Signaling | 0.00E+00 | 9.26E-03 | NaN |
| SAPK/JNK Signaling | 0.00E+00 | 1.99E-03 | NaN |
| Cardiac Œ≤-adrenergic Signaling | 0.00E+00 | 5.59E-03 | NaN |
| B Cell Receptor Signaling | 0.00E+00 | 1.58E-03 | NaN |
| Insulin Receptor Signaling | 0.00E+00 | 7.14E-03 | NaN |
| Integrin Signaling | 0.00E+00 | 4.69E-03 | NaN |
| cAMP-mediated signaling | 0.00E+00 | 4.26E-03 | NaN |
| Apoptosis Signaling | 0.00E+00 | 9.62E-03 | NaN |
| NF-Œ∫B Signaling | 0.00E+00 | 8.77E-03 | 2.236 |
| T Cell Receptor Signaling | 0.00E+00 | 3.26E-03 | NaN |
| GPCR-Mediated Nutrient Sensing in Enteroendocrine Cells | 0.00E+00 | 8.47E-03 | NaN |
| Phagosome Maturation | 0.00E+00 | 6.29E-03 | NaN |
| Sumoylation Pathway | 0.00E+00 | 9.71E-03 | NaN |
| Th2 Pathway | 0.00E+00 | 7.30E-03 | NaN |
| Sirtuin Signaling Pathway | 0.00E+00 | 6.85E-03 | NaN |
| Opioid Signaling Pathway | 0.00E+00 | 3.57E-03 | NaN |
| Iron homeostasis signaling pathway | 0.00E+00 | 7.19E-03 | NaN |
| Endocannabinoid Developing Neuron Pathway | 0.00E+00 | 7.87E-03 | NaN |
| Cardiac Hypertrophy Signaling (Enhanced) | 0.00E+00 | 7.37E-03 | NaN |
| Synaptogenesis Signaling Pathway | 0.00E+00 | 9.46E-03 | NaN |
| Systemic Lupus Erythematosus In T Cell Signaling Pathway | 0.00E+00 | 3.12E-03 | NaN |
| White Adipose Tissue Browning Pathway | 0.00E+00 | 7.25E-03 | NaN |
| Hepatic Fibrosis Signaling Pathway | 0.00E+00 | 9.48E-03 | 2 |
| Xenobiotic Metabolism General Signaling Pathway | 0.00E+00 | 6.99E-03 | NaN |
| Semaphorin Neuronal Repulsive Signaling Pathway | 0.00E+00 | 6.62E-03 | NaN |
| MSP-RON Signaling In Cancer Cells Pathway | 0.00E+00 | 7.14E-03 | NaN |
| Role of MAPK Signaling in Promoting the Pathogenesis of Influenza | 0.00E+00 | 8.85E-03 | NaN |

**Supplemental Table 9. Complete IPA analysis, day 0 vs day 10**

| **Ingenuity Canonical Pathways** | **-log(p-value)** | **Ratio** | **z-score** |
| --- | --- | --- | --- |
| Systemic Lupus Erythematosus In B Cell Signaling Pathway | 2.14E+01 | 5.67E-02 | 2.828 |
| IL-15 Signaling | 2.04E+01 | 6.63E-02 | NaN |
| B Cell Receptor Signaling | 1.79E+01 | 5.55E-02 | NaN |
| Communication between Innate and Adaptive Immune Cells | 1.35E+01 | 3.88E-02 | NaN |
| Interferon Signaling | 1.24E+01 | 2.78E-01 | 2.53 |
| Role of Hypercytokinemia/hyperchemokinemia in the Pathogenesis of Influenza | 1.10E+01 | 1.40E-01 | 3.464 |
| Primary Immunodeficiency Signaling | 7.59E+00 | 1.43E-01 | NaN |
| Role of Pattern Recognition Receptors in Recognition of Bacteria and Viruses | 4.20E+00 | 5.13E-02 | 2 |
| Activation of IRF by Cytosolic Pattern Recognition Receptors | 3.61E+00 | 7.69E-02 | 0.447 |
| Pyrimidine Deoxyribonucleotides De Novo Biosynthesis I | 3.01E+00 | 1.30E-01 | NaN |
| Coronavirus Pathogenesis Pathway | 2.72E+00 | 3.45E-02 | -1.633 |
| NAD Signaling Pathway | 2.70E+00 | 3.97E-02 | 2.449 |
| Pathogenesis of Multiple Sclerosis | 2.60E+00 | 2.22E-01 | NaN |
| Complement System | 2.41E+00 | 8.11E-02 | NaN |
| IL-7 Signaling Pathway | 2.34E+00 | 5.13E-02 | NaN |
| B Cell Development | 2.19E+00 | 6.82E-02 | NaN |
| Coronavirus Replication Pathway | 2.17E+00 | 6.67E-02 | NaN |
| Pyroptosis Signaling Pathway | 2.07E+00 | 4.30E-02 | 2 |
| Cell Cycle Control of Chromosomal Replication | 1.91E+00 | 5.36E-02 | NaN |
| Phagosome Formation | 1.78E+00 | 1.74E-02 | 2.714 |
| Role of JAK family kinases in IL-6-type Cytokine Signaling | 1.71E+00 | 8.00E-02 | NaN |
| IL-17A Signaling in Gastric Cells | 1.68E+00 | 7.69E-02 | NaN |
| Role of JAK1, JAK2 and TYK2 in Interferon Signaling | 1.68E+00 | 7.69E-02 | NaN |
| Estrogen-mediated S-phase Entry | 1.68E+00 | 7.69E-02 | NaN |
| Role of Macrophages, Fibroblasts and Endothelial Cells in Rheumatoid Arthritis | 1.63E+00 | 2.13E-02 | NaN |
| Granulocyte Adhesion and Diapedesis | 1.62E+00 | 2.65E-02 | NaN |
| Role of PKR in Interferon Induction and Antiviral Response | 1.52E+00 | 2.94E-02 | 2 |
| Inhibition of Angiogenesis by TSP1 | 1.46E+00 | 5.88E-02 | NaN |
| Role of JAK2 in Hormone-like Cytokine Signaling | 1.46E+00 | 5.88E-02 | NaN |
| Pyrimidine Ribonucleotides Interconversion | 1.44E+00 | 5.71E-02 | NaN |
| Hematopoiesis from Pluripotent Stem Cells | 1.42E+00 | 1.81E-02 | NaN |
| Pyrimidine Ribonucleotides De Novo Biosynthesis | 1.37E+00 | 5.26E-02 | NaN |
| dTMP De Novo Biosynthesis | 1.37E+00 | 2.00E-01 | NaN |
| Inhibition of Matrix Metalloproteases | 1.35E+00 | 5.13E-02 | NaN |
| Necroptosis Signaling Pathway | 1.33E+00 | 2.55E-02 | 2 |
| Salvage Pathways of Pyrimidine Ribonucleotides | 1.28E+00 | 3.06E-02 | NaN |
| UVA-Induced MAPK Signaling | 1.28E+00 | 3.06E-02 | NaN |
| Oncostatin M Signaling | 1.28E+00 | 4.65E-02 | NaN |
| Role of RIG1-like Receptors in Antiviral Innate Immunity | 1.23E+00 | 4.35E-02 | NaN |
| Role of IL-17F in Allergic Inflammatory Airway Diseases | 1.21E+00 | 4.26E-02 | NaN |
| Cell Cycle: G2/M DNA Damage Checkpoint Regulation | 1.16E+00 | 4.00E-02 | NaN |
| UVB-Induced MAPK Signaling | 1.13E+00 | 3.85E-02 | NaN |
| Acute Phase Response Signaling | 1.12E+00 | 2.16E-02 | NaN |
| IL-13 Signaling Pathway | 1.11E+00 | 2.59E-02 | NaN |
| Transcriptional Regulatory Network in Embryonic Stem Cells | 1.10E+00 | 3.70E-02 | NaN |
| p38 MAPK Signaling | 1.07E+00 | 2.50E-02 | NaN |
| Role of CHK Proteins in Cell Cycle Checkpoint Control | 1.06E+00 | 3.51E-02 | NaN |
| Th1 Pathway | 1.06E+00 | 2.46E-02 | NaN |
| IL-15 Production | 1.05E+00 | 2.44E-02 | NaN |
| Triacylglycerol Degradation | 1.04E+00 | 3.39E-02 | NaN |
| Retinoic acid Mediated Apoptosis Signaling | 1.03E+00 | 3.33E-02 | NaN |
| ID1 Signaling Pathway | 1.03E+00 | 2.00E-02 | 0 |
| Gustation Pathway | 1.01E+00 | 1.97E-02 | -1 |
| Mitotic Roles of Polo-Like Kinase | 9.59E-01 | 3.03E-02 | NaN |
| Pyridoxal 5'-phosphate Salvage Pathway | 9.59E-01 | 3.03E-02 | NaN |
| Role of IL-17A in Psoriasis | 9.43E-01 | 7.14E-02 | NaN |
| Role of JAK1 and JAK3 in Œ≥c Cytokine Signaling | 9.24E-01 | 2.90E-02 | NaN |
| Growth Hormone Signaling | 9.07E-01 | 2.82E-02 | NaN |
| Extrinsic Prothrombin Activation Pathway | 8.89E-01 | 6.25E-02 | NaN |
| Parkinson's Signaling | 8.89E-01 | 6.25E-02 | NaN |
| Role of MAPK Signaling in Inhibiting the Pathogenesis of Influenza | 8.48E-01 | 2.60E-02 | NaN |
| VDR/RXR Activation | 8.39E-01 | 2.56E-02 | NaN |
| Granzyme A Signaling | 8.21E-01 | 5.26E-02 | NaN |
| DNA damage-induced 14-3-3œÉ Signaling | 8.21E-01 | 5.26E-02 | NaN |
| JAK/STAT Signaling | 8.04E-01 | 2.44E-02 | NaN |
| Inflammasome pathway | 7.99E-01 | 5.00E-02 | NaN |
| Cyclins and Cell Cycle Regulation | 7.88E-01 | 2.38E-02 | NaN |
| FcŒ≥RIIB Signaling in B Lymphocytes | 7.80E-01 | 2.35E-02 | NaN |
| Prolactin Signaling | 7.72E-01 | 2.33E-02 | NaN |
| PDGF Signaling | 7.72E-01 | 2.33E-02 | NaN |
| Androgen Signaling | 7.50E-01 | 1.78E-02 | NaN |
| Role of Lipids/Lipid Rafts in the Pathogenesis of Influenza | 7.45E-01 | 4.35E-02 | NaN |
| Th1 and Th2 Activation Pathway | 7.33E-01 | 1.74E-02 | NaN |
| IL-22 Signaling | 7.28E-01 | 4.17E-02 | NaN |
| Death Receptor Signaling | 6.99E-01 | 2.08E-02 | NaN |
| TGF-Œ≤ Signaling | 6.99E-01 | 2.08E-02 | NaN |
| ATM Signaling | 6.80E-01 | 2.02E-02 | NaN |
| Protein Ubiquitination Pathway | 6.74E-01 | 1.45E-02 | NaN |
| Sonic Hedgehog Signaling | 6.54E-01 | 3.45E-02 | NaN |
| PD-1, PD-L1 cancer immunotherapy pathway | 6.36E-01 | 1.89E-02 | NaN |
| Sirtuin Signaling Pathway | 6.14E-01 | 1.37E-02 | NaN |
| Kinetochore Metaphase Signaling Pathway | 6.07E-01 | 1.80E-02 | NaN |
| Dendritic Cell Maturation | 6.06E-01 | 1.18E-02 | 2.449 |
| Senescence Pathway | 5.95E-01 | 1.34E-02 | NaN |
| Ephrin Receptor Signaling | 5.95E-01 | 1.48E-02 | NaN |
| Coagulation System | 5.83E-01 | 2.86E-02 | NaN |
| IL-9 Signaling | 5.83E-01 | 2.86E-02 | NaN |
| MSP-RON Signaling In Macrophages Pathway | 5.64E-01 | 1.68E-02 | NaN |
| Glutathione-mediated Detoxification | 5.62E-01 | 2.70E-02 | NaN |
| Protein Kinase A Signaling | 5.59E-01 | 1.22E-02 | 0.447 |
| Agranulocyte Adhesion and Diapedesis | 5.54E-01 | 1.40E-02 | NaN |
| Notch Signaling | 5.53E-01 | 2.63E-02 | NaN |
| ERK/MAPK Signaling | 5.47E-01 | 1.39E-02 | NaN |
| Systemic Lupus Erythematosus Signaling | 5.41E-01 | 1.13E-02 | NaN |
| FXR/RXR Activation | 5.30E-01 | 1.59E-02 | NaN |
| IL-6 Signaling | 5.21E-01 | 1.56E-02 | NaN |
| Intrinsic Prothrombin Activation Pathway | 5.17E-01 | 2.38E-02 | NaN |
| Ferroptosis Signaling Pathway | 5.03E-01 | 1.52E-02 | NaN |
| STAT3 Pathway | 4.91E-01 | 1.48E-02 | NaN |
| iNOS Signaling | 4.76E-01 | 2.13E-02 | NaN |
| Retinol Biosynthesis | 4.76E-01 | 2.13E-02 | NaN |
| Reelin Signaling in Neurons | 4.74E-01 | 1.44E-02 | NaN |
| nNOS Signaling in Skeletal Muscle Cells | 4.70E-01 | 2.08E-02 | NaN |
| PI3K Signaling in B Lymphocytes | 4.58E-01 | 1.40E-02 | NaN |
| Autoimmune Thyroid Disease Signaling | 4.53E-01 | 1.10E-02 | NaN |
| EGF Signaling | 4.23E-01 | 1.82E-02 | NaN |
| Type II Diabetes Mellitus Signaling | 4.21E-01 | 1.31E-02 | NaN |
| CNTF Signaling | 4.10E-01 | 1.75E-02 | NaN |
| Role of IL-17A in Arthritis | 4.10E-01 | 1.75E-02 | NaN |
| Allograft Rejection Signaling | 3.95E-01 | 1.03E-02 | NaN |
| GADD45 Signaling | 3.93E-01 | 1.67E-02 | NaN |
| Circadian Rhythm Signaling | 3.89E-01 | 1.12E-02 | NaN |
| HOTAIR Regulatory Pathway | 3.88E-01 | 1.23E-02 | NaN |
| IL-2 Signaling | 3.87E-01 | 1.64E-02 | NaN |
| Thrombopoietin Signaling | 3.77E-01 | 1.59E-02 | NaN |
| Germ Cell-Sertoli Cell Junction Signaling | 3.64E-01 | 1.17E-02 | NaN |
| IL-17A Signaling in Airway Cells | 3.57E-01 | 1.49E-02 | NaN |
| Cell Cycle: G1/S Checkpoint Regulation | 3.53E-01 | 1.47E-02 | NaN |
| SPINK1 General Cancer Pathway | 3.48E-01 | 1.45E-02 | NaN |
| GM-CSF Signaling | 3.43E-01 | 1.43E-02 | NaN |
| Tumor Microenvironment Pathway | 3.41E-01 | 1.12E-02 | NaN |
| Ephrin B Signaling | 3.34E-01 | 1.39E-02 | NaN |
| Netrin Signaling | 3.34E-01 | 1.39E-02 | NaN |
| D-myo-inositol (1,4,5,6)-Tetrakisphosphate Biosynthesis | 3.33E-01 | 1.10E-02 | NaN |
| D-myo-inositol (3,4,5,6)-tetrakisphosphate Biosynthesis | 3.33E-01 | 1.10E-02 | NaN |
| ERK5 Signaling | 3.26E-01 | 1.35E-02 | NaN |
| Hypoxia Signaling in the Cardiovascular System | 3.17E-01 | 1.32E-02 | NaN |
| Angiopoietin Signaling | 3.13E-01 | 1.30E-02 | NaN |
| Hepatic Cholestasis | 3.10E-01 | 1.05E-02 | NaN |
| NF-Œ∫B Activation by Viruses | 3.10E-01 | 1.28E-02 | NaN |
| Toll-like Receptor Signaling | 3.10E-01 | 1.28E-02 | NaN |
| IL-3 Signaling | 3.05E-01 | 1.27E-02 | NaN |
| Maturity Onset Diabetes of Young (MODY) Signaling | 3.05E-01 | 1.27E-02 | NaN |
| 3-phosphoinositide Degradation | 3.03E-01 | 1.03E-02 | NaN |
| Role of BRCA1 in DNA Damage Response | 3.02E-01 | 1.25E-02 | NaN |
| Heparan Sulfate Biosynthesis (Late Stages) | 2.98E-01 | 1.23E-02 | NaN |
| BEX2 Signaling Pathway | 2.98E-01 | 1.23E-02 | NaN |
| FLT3 Signaling in Hematopoietic Progenitor Cells | 2.94E-01 | 1.22E-02 | NaN |
| Role of MAPK Signaling in the Pathogenesis of Influenza | 2.94E-01 | 1.22E-02 | NaN |
| D-myo-inositol-5-phosphate Metabolism | 2.93E-01 | 1.01E-02 | NaN |
| Neuroinflammation Signaling Pathway | 2.89E-01 | 9.46E-03 | NaN |
| Synaptogenesis Signaling Pathway | 2.89E-01 | 9.46E-03 | NaN |
| Phospholipase C Signaling | 2.76E-01 | 8.84E-03 | NaN |
| Heparan Sulfate Biosynthesis | 2.73E-01 | 1.14E-02 | NaN |
| 3-phosphoinositide Biosynthesis | 2.72E-01 | 9.62E-03 | NaN |
| Regulation of Cellular Mechanics by Calpain Protease | 2.70E-01 | 1.12E-02 | NaN |
| Acute Myeloid Leukemia Signaling | 2.64E-01 | 1.10E-02 | NaN |
| IL-4 Signaling | 2.57E-01 | 1.08E-02 | NaN |
| FcŒ≥ Receptor-mediated Phagocytosis in Macrophages and Monocytes | 2.55E-01 | 1.06E-02 | NaN |
| Role of NFAT in Cardiac Hypertrophy | 2.43E-01 | 9.01E-03 | NaN |
| p53 Signaling | 2.43E-01 | 1.02E-02 | NaN |
| EIF2 Signaling | 2.40E-01 | 8.93E-03 | NaN |
| Role of Osteoblasts, Osteoclasts and Chondrocytes in Rheumatoid Arthritis | 2.40E-01 | 8.93E-03 | NaN |
| Neurovascular Coupling Signaling Pathway | 2.38E-01 | 8.89E-03 | NaN |
| NER (Nucleotide Excision Repair, Enhanced Pathway) | 2.29E-01 | 9.71E-03 | NaN |
| Mouse Embryonic Stem Cell Pluripotency | 2.27E-01 | 9.62E-03 | NaN |
| IGF-1 Signaling | 2.27E-01 | 9.62E-03 | NaN |
| Apoptosis Signaling | 2.27E-01 | 9.62E-03 | NaN |
| Amyotrophic Lateral Sclerosis Signaling | 1.99E-01 | 8.62E-03 | NaN |
| Dilated Cardiomyopathy Signaling Pathway | 0.00E+00 | 6.76E-03 | NaN |
| Oxytocin In Brain Signaling Pathway | 0.00E+00 | 5.08E-03 | NaN |
| Oxytocin Signaling Pathway | 0.00E+00 | 3.55E-03 | NaN |
| Pulmonary Fibrosis Idiopathic Signaling Pathway | 0.00E+00 | 6.13E-03 | NaN |
| Wound Healing Signaling Pathway | 0.00E+00 | 7.94E-03 | NaN |
| Glucocorticoid Receptor Signaling | 0.00E+00 | 6.88E-03 | NaN |
| Axonal Guidance Signaling | 0.00E+00 | 5.89E-03 | NaN |
| Synaptic Long Term Depression | 0.00E+00 | 5.13E-03 | NaN |
| LPS/IL-1 Mediated Inhibition of RXR Function | 0.00E+00 | 3.94E-03 | NaN |
| Hepatic Fibrosis / Hepatic Stellate Cell Activation | 0.00E+00 | 5.15E-03 | NaN |
| Erythropoietin Signaling Pathway | 0.00E+00 | 5.65E-03 | NaN |
| Clathrin-mediated Endocytosis Signaling | 0.00E+00 | 5.18E-03 | NaN |
| IL-8 Signaling | 0.00E+00 | 4.76E-03 | NaN |
| IL-12 Signaling and Production in Macrophages | 0.00E+00 | 7.35E-03 | NaN |
| Role of NFAT in Regulation of the Immune Response | 0.00E+00 | 3.40E-03 | NaN |
| CCR5 Signaling in Macrophages | 0.00E+00 | 2.00E-03 | NaN |
| IL-17 Signaling | 0.00E+00 | 5.35E-03 | NaN |
| Airway Pathology in Chronic Obstructive Pulmonary Disease | 0.00E+00 | 8.47E-03 | NaN |
| T Helper Cell Differentiation | 0.00E+00 | 2.12E-03 | NaN |
| Cellular Effects of Sildenafil (Viagra) | 0.00E+00 | 6.67E-03 | NaN |
| Renin-Angiotensin Signaling | 0.00E+00 | 8.20E-03 | NaN |
| Thrombin Signaling | 0.00E+00 | 4.44E-03 | NaN |
| Cardiac Hypertrophy Signaling | 0.00E+00 | 3.83E-03 | NaN |
| Molecular Mechanisms of Cancer | 0.00E+00 | 4.47E-03 | NaN |
| Corticotropin Releasing Hormone Signaling | 0.00E+00 | 6.62E-03 | NaN |
| HMGB1 Signaling | 0.00E+00 | 5.99E-03 | NaN |
| GNRH Signaling | 0.00E+00 | 5.21E-03 | NaN |
| Role of NANOG in Mammalian Embryonic Stem Cell Pluripotency | 0.00E+00 | 8.33E-03 | NaN |
| CREB Signaling in Neurons | 0.00E+00 | 8.25E-03 | 0.447 |
| Type I Diabetes Mellitus Signaling | 0.00E+00 | 3.93E-03 | NaN |
| Production of Nitric Oxide and Reactive Oxygen Species in Macrophages | 0.00E+00 | 5.24E-03 | NaN |
| GŒ±12/13 Signaling | 0.00E+00 | 7.52E-03 | NaN |
| p70S6K Signaling | 0.00E+00 | 7.58E-03 | NaN |
| Colorectal Cancer Metastasis Signaling | 0.00E+00 | 3.69E-03 | NaN |
| Pancreatic Adenocarcinoma Signaling | 0.00E+00 | 7.94E-03 | NaN |
| G Beta Gamma Signaling | 0.00E+00 | 7.75E-03 | NaN |
| FAK Signaling | 0.00E+00 | 4.80E-03 | 0.447 |
| Hereditary Breast Cancer Signaling | 0.00E+00 | 7.04E-03 | NaN |
| RHOA Signaling | 0.00E+00 | 8.06E-03 | NaN |
| Altered T Cell and B Cell Signaling in Rheumatoid Arthritis | 0.00E+00 | 2.03E-03 | NaN |
| Breast Cancer Regulation by Stathmin1 | 0.00E+00 | 8.42E-03 | 0.447 |
| Neuroprotective Role of THOP1 in Alzheimer's Disease | 0.00E+00 | 8.33E-03 | NaN |
| PKCŒ∏ Signaling in T Lymphocytes | 0.00E+00 | 1.79E-03 | NaN |
| Dopamine-DARPP32 Feedback in cAMP Signaling | 0.00E+00 | 5.43E-03 | NaN |
| Signaling by Rho Family GTPases | 0.00E+00 | 7.46E-03 | NaN |
| RHOGDI Signaling | 0.00E+00 | 4.65E-03 | NaN |
| Sertoli Cell-Sertoli Cell Junction Signaling | 0.00E+00 | 4.85E-03 | NaN |
| Superpathway of Inositol Phosphate Compounds | 0.00E+00 | 8.44E-03 | NaN |
| Epithelial Adherens Junction Signaling | 0.00E+00 | 6.37E-03 | NaN |
| Regulation of the Epithelial-Mesenchymal Transition Pathway | 0.00E+00 | 5.13E-03 | NaN |
| Sperm Motility | 0.00E+00 | 7.84E-03 | NaN |
| TEC Kinase Signaling | 0.00E+00 | 1.73E-03 | NaN |
| Estrogen Receptor Signaling | 0.00E+00 | 4.89E-03 | NaN |
| Nitric Oxide Signaling in the Cardiovascular System | 0.00E+00 | 8.33E-03 | NaN |
| Cardiac Œ≤-adrenergic Signaling | 0.00E+00 | 5.59E-03 | NaN |
| GABA Receptor Signaling | 0.00E+00 | 7.52E-03 | NaN |
| Xenobiotic Metabolism Signaling | 0.00E+00 | 3.45E-03 | NaN |
| WNT/Œ≤-catenin Signaling | 0.00E+00 | 5.78E-03 | NaN |
| NF-Œ∫B Signaling | 0.00E+00 | 1.75E-03 | NaN |
| G-Protein Coupled Receptor Signaling | 0.00E+00 | 5.69E-03 | 1 |
| GPCR-Mediated Nutrient Sensing in Enteroendocrine Cells | 0.00E+00 | 8.47E-03 | NaN |
| Phagosome Maturation | 0.00E+00 | 6.29E-03 | NaN |
| Osteoarthritis Pathway | 0.00E+00 | 8.47E-03 | NaN |
| Opioid Signaling Pathway | 0.00E+00 | 3.57E-03 | NaN |
| Endocannabinoid Neuronal Synapse Pathway | 0.00E+00 | 6.71E-03 | NaN |
| Endocannabinoid Cancer Inhibition Pathway | 0.00E+00 | 6.80E-03 | NaN |
| Apelin Endothelial Signaling Pathway | 0.00E+00 | 7.09E-03 | NaN |
| Cardiac Hypertrophy Signaling (Enhanced) | 0.00E+00 | 3.68E-03 | NaN |
| T Cell Exhaustion Signaling Pathway | 0.00E+00 | 5.30E-03 | NaN |
| White Adipose Tissue Browning Pathway | 0.00E+00 | 7.25E-03 | NaN |
| Hepatic Fibrosis Signaling Pathway | 0.00E+00 | 4.74E-03 | NaN |
| Xenobiotic Metabolism PXR Signaling Pathway | 0.00E+00 | 5.18E-03 | NaN |
| Insulin Secretion Signaling Pathway | 0.00E+00 | 7.33E-03 | NaN |
| Semaphorin Neuronal Repulsive Signaling Pathway | 0.00E+00 | 6.62E-03 | NaN |
| Regulation Of The Epithelial Mesenchymal Transition By Growth Factors Pathway | 0.00E+00 | 5.21E-03 | NaN |
| Calcium Signaling | 0.00E+00 | 4.59E-03 | NaN |

**Supplemental Table 10. IPA analysis, day 0 vs day 14**

| **Ingenuity Canonical Pathways** | **-log(p-value)** | **Ratio** | **z-score** |
| --- | --- | --- | --- |
| IL-15 Signaling | 1.08E+02 | 2.05E-01 | NaN |
| B Cell Receptor Signaling | 9.91E+01 | 1.71E-01 | NaN |
| Systemic Lupus Erythematosus In B Cell Signaling Pathway | 9.24E+01 | 1.49E-01 | NaN |
| Communication between Innate and Adaptive Immune Cells | 8.17E+01 | 1.17E-01 | NaN |
| Kinetochore Metaphase Signaling Pathway | 1.54E+01 | 1.62E-01 | 3.207 |
| Primary Immunodeficiency Signaling | 1.06E+01 | 1.96E-01 | NaN |
| Estrogen-mediated S-phase Entry | 7.95E+00 | 2.69E-01 | 2.646 |
| Mitotic Roles of Polo-Like Kinase | 7.29E+00 | 1.36E-01 | 1.414 |
| Cyclins and Cell Cycle Regulation | 6.37E+00 | 1.07E-01 | 3 |
| Role of CHK Proteins in Cell Cycle Checkpoint Control | 5.48E+00 | 1.23E-01 | -2.236 |
| DNA Methylation and Transcriptional Repression Signaling | 5.22E+00 | 1.46E-01 | NaN |
| Cell Cycle: G2/M DNA Damage Checkpoint Regulation | 4.71E+00 | 1.20E-01 | -1.633 |
| NER (Nucleotide Excision Repair, Enhanced Pathway) | 4.68E+00 | 7.77E-02 | -1.89 |
| Transcriptional Regulatory Network in Embryonic Stem Cells | 4.51E+00 | 1.11E-01 | NaN |
| Cell Cycle Control of Chromosomal Replication | 4.42E+00 | 1.07E-01 | 2.449 |
| DNA damage-induced 14-3-3œÉ Signaling | 4.28E+00 | 2.11E-01 | NaN |
| NAD Signaling Pathway | 3.51E+00 | 5.30E-02 | 2.828 |
| Cell Cycle Regulation by BTG Family Proteins | 3.12E+00 | 1.08E-01 | 2 |
| ATM Signaling | 3.06E+00 | 6.06E-02 | -0.816 |
| Cell Cycle: G1/S Checkpoint Regulation | 3.00E+00 | 7.35E-02 | -2.236 |
| Role of BRCA1 in DNA Damage Response | 2.69E+00 | 6.25E-02 | -0.447 |
| Pancreatic Adenocarcinoma Signaling | 2.52E+00 | 4.76E-02 | NaN |
| Ferroptosis Signaling Pathway | 2.42E+00 | 4.55E-02 | -2.236 |
| GADD45 Signaling | 2.34E+00 | 6.67E-02 | -1 |
| ID1 Signaling Pathway | 2.13E+00 | 3.50E-02 | 1.89 |
| Glutamate Removal from Folates | 1.95E+00 | 1.00E+00 | NaN |
| B Cell Development | 1.88E+00 | 6.82E-02 | NaN |
| BER (Base Excision Repair) Pathway | 1.88E+00 | 6.82E-02 | NaN |
| Mismatch Repair in Eukaryotes | 1.87E+00 | 1.25E-01 | NaN |
| Granzyme A Signaling | 1.73E+00 | 1.05E-01 | NaN |
| Senescence Pathway | 1.71E+00 | 2.68E-02 | -2.646 |
| Hereditary Breast Cancer Signaling | 1.66E+00 | 3.52E-02 | NaN |
| Pyrimidine Deoxyribonucleotides De Novo Biosynthesis I | 1.57E+00 | 8.70E-02 | NaN |
| Hematopoiesis from Pluripotent Stem Cells | 1.55E+00 | 2.26E-02 | NaN |
| Role of Macrophages, Fibroblasts and Endothelial Cells in Rheumatoid Arthritis | 1.49E+00 | 2.44E-02 | NaN |
| Serine Biosynthesis | 1.48E+00 | 3.33E-01 | NaN |
| Sonic Hedgehog Signaling | 1.38E+00 | 6.90E-02 | NaN |
| Glioblastoma Multiforme Signaling | 1.36E+00 | 2.92E-02 | NaN |
| Proline Biosynthesis I | 1.36E+00 | 2.50E-01 | NaN |
| Phagosome Formation | 1.32E+00 | 1.88E-02 | 3.464 |
| Glioma Signaling | 1.29E+00 | 3.23E-02 | NaN |
| Proline Biosynthesis II (from Arginine) | 1.26E+00 | 2.00E-01 | NaN |
| Superpathway of Serine and Glycine Biosynthesis I | 1.26E+00 | 2.00E-01 | NaN |
| dTMP De Novo Biosynthesis | 1.26E+00 | 2.00E-01 | NaN |
| IL-7 Signaling Pathway | 1.24E+00 | 3.85E-02 | NaN |
| Chondroitin and Dermatan Biosynthesis | 1.19E+00 | 1.67E-01 | NaN |
| Arginine Degradation VI (Arginase 2 Pathway) | 1.19E+00 | 1.67E-01 | NaN |
| Glycine Cleavage Complex | 1.19E+00 | 1.67E-01 | NaN |
| Autoimmune Thyroid Disease Signaling | 1.15E+00 | 1.98E-02 | NaN |
| Regulation of Cellular Mechanics by Calpain Protease | 1.11E+00 | 3.37E-02 | NaN |
| p53 Signaling | 1.01E+00 | 3.06E-02 | NaN |
| Salvage Pathways of Pyrimidine Ribonucleotides | 1.01E+00 | 3.06E-02 | NaN |
| FAT10 Cancer Signaling Pathway | 9.67E-01 | 4.00E-02 | NaN |
| UVB-Induced MAPK Signaling | 9.39E-01 | 3.85E-02 | NaN |
| Chronic Myeloid Leukemia Signaling | 9.24E-01 | 2.80E-02 | NaN |
| DNA Double-Strand Break Repair by Homologous Recombination | 8.36E-01 | 7.14E-02 | NaN |
| Parkinson's Signaling | 7.83E-01 | 6.25E-02 | NaN |
| Pyridoxal 5'-phosphate Salvage Pathway | 7.72E-01 | 3.03E-02 | NaN |
| Allograft Rejection Signaling | 7.52E-01 | 1.65E-02 | NaN |
| Coronavirus Pathogenesis Pathway | 7.14E-01 | 1.97E-02 | -2 |
| Sirtuin Signaling Pathway | 6.38E-01 | 1.71E-02 | 1.342 |
| Aryl Hydrocarbon Receptor Signaling | 5.80E-01 | 1.89E-02 | NaN |
| Systemic Lupus Erythematosus Signaling | 5.80E-01 | 1.45E-02 | NaN |
| Role of p14/p19ARF in Tumor Suppression | 5.54E-01 | 3.45E-02 | NaN |
| Small Cell Lung Cancer Signaling | 5.35E-01 | 2.08E-02 | NaN |
| Inhibition of Angiogenesis by TSP1 | 4.98E-01 | 2.94E-02 | NaN |
| Pyrimidine Ribonucleotides Interconversion | 4.87E-01 | 2.86E-02 | NaN |
| Glutathione-mediated Detoxification | 4.67E-01 | 2.70E-02 | NaN |
| Complement System | 4.67E-01 | 2.70E-02 | NaN |
| Dendritic Cell Maturation | 4.62E-01 | 1.35E-02 | 2.646 |
| Breast Cancer Regulation by Stathmin1 | 4.58E-01 | 1.35E-02 | 2.121 |
| Pyrimidine Ribonucleotides De Novo Biosynthesis | 4.58E-01 | 2.63E-02 | NaN |
| Phospholipase C Signaling | 4.55E-01 | 1.33E-02 | NaN |
| Inhibition of Matrix Metalloproteases | 4.49E-01 | 2.56E-02 | NaN |
| Prostate Cancer Signaling | 4.38E-01 | 1.75E-02 | NaN |
| April Mediated Signaling | 4.24E-01 | 2.38E-02 | NaN |
| B Cell Activating Factor Signaling | 4.16E-01 | 2.33E-02 | NaN |
| Molecular Mechanisms of Cancer | 4.16E-01 | 1.34E-02 | NaN |
| p38 MAPK Signaling | 4.10E-01 | 1.67E-02 | NaN |
| Gustation Pathway | 4.01E-01 | 1.48E-02 | NaN |
| Role of IL-17F in Allergic Inflammatory Airway Diseases | 3.86E-01 | 2.13E-02 | NaN |
| FXR/RXR Activation | 3.84E-01 | 1.59E-02 | NaN |
| Melanoma Signaling | 3.66E-01 | 2.00E-02 | NaN |
| Chondroitin Sulfate Biosynthesis (Late Stages) | 3.66E-01 | 2.00E-02 | NaN |
| STAT3 Pathway | 3.50E-01 | 1.48E-02 | NaN |
| Neurovascular Coupling Signaling Pathway | 3.35E-01 | 1.33E-02 | NaN |
| Chondroitin Sulfate Biosynthesis | 3.19E-01 | 1.72E-02 | NaN |
| Protein Kinase A Signaling | 3.15E-01 | 1.22E-02 | 1.342 |
| Dermatan Sulfate Biosynthesis | 3.09E-01 | 1.67E-02 | NaN |
| Ovarian Cancer Signaling | 2.76E-01 | 1.27E-02 | NaN |
| Phospholipases | 2.76E-01 | 1.49E-02 | NaN |
| eNOS Signaling | 2.73E-01 | 1.26E-02 | NaN |
| Eicosanoid Signaling | 2.72E-01 | 1.47E-02 | NaN |
| SPINK1 General Cancer Pathway | 2.67E-01 | 1.45E-02 | NaN |
| HOTAIR Regulatory Pathway | 2.62E-01 | 1.23E-02 | NaN |
| Growth Hormone Signaling | 2.59E-01 | 1.41E-02 | NaN |
| Glioma Invasiveness Signaling | 2.51E-01 | 1.37E-02 | NaN |
| Androgen Signaling | 2.47E-01 | 1.18E-02 | NaN |
| Caveolar-mediated Endocytosis Signaling | 2.43E-01 | 1.33E-02 | NaN |
| Hypoxia Signaling in the Cardiovascular System | 2.40E-01 | 1.32E-02 | NaN |
| Angiopoietin Signaling | 2.37E-01 | 1.30E-02 | NaN |
| Role of MAPK Signaling in Inhibiting the Pathogenesis of Influenza | 2.37E-01 | 1.30E-02 | NaN |
| Thyroid Cancer Signaling | 2.29E-01 | 1.27E-02 | NaN |
| Estrogen-Dependent Breast Cancer Signaling | 2.23E-01 | 1.23E-02 | NaN |
| Role of MAPK Signaling in the Pathogenesis of Influenza | 2.20E-01 | 1.22E-02 | NaN |
| FcŒ≥RIIB Signaling in B Lymphocytes | 2.10E-01 | 1.18E-02 | NaN |
| PDGF Signaling | 2.07E-01 | 1.16E-02 | NaN |
| Oxytocin In Brain Signaling Pathway | 0.00E+00 | 5.08E-03 | NaN |
| Pulmonary Fibrosis Idiopathic Signaling Pathway | 0.00E+00 | 6.13E-03 | NaN |
| Pulmonary Healing Signaling Pathway | 0.00E+00 | 5.03E-03 | NaN |
| CLEAR Signaling Pathway | 0.00E+00 | 3.51E-03 | NaN |
| Glucocorticoid Receptor Signaling | 0.00E+00 | 6.88E-03 | NaN |
| Circadian Rhythm Signaling | 0.00E+00 | 7.43E-03 | NaN |
| Axonal Guidance Signaling | 0.00E+00 | 3.93E-03 | NaN |
| Amyotrophic Lateral Sclerosis Signaling | 0.00E+00 | 8.62E-03 | NaN |
| Synaptic Long Term Depression | 0.00E+00 | 1.03E-02 | NaN |
| Huntington's Disease Signaling | 0.00E+00 | 3.53E-03 | NaN |
| LPS/IL-1 Mediated Inhibition of RXR Function | 0.00E+00 | 3.94E-03 | NaN |
| Acute Phase Response Signaling | 0.00E+00 | 5.41E-03 | NaN |
| Hepatic Cholestasis | 0.00E+00 | 5.24E-03 | NaN |
| Hepatic Fibrosis / Hepatic Stellate Cell Activation | 0.00E+00 | 5.15E-03 | NaN |
| Clathrin-mediated Endocytosis Signaling | 0.00E+00 | 5.18E-03 | NaN |
| Role of PKR in Interferon Induction and Antiviral Response | 0.00E+00 | 7.35E-03 | NaN |
| Role of NFAT in Regulation of the Immune Response | 0.00E+00 | 1.70E-03 | NaN |
| Virus Entry via Endocytic Pathways | 0.00E+00 | 9.62E-03 | NaN |
| Reelin Signaling in Neurons | 0.00E+00 | 7.19E-03 | NaN |
| HIF1Œ± Signaling | 0.00E+00 | 4.81E-03 | NaN |
| Cellular Effects of Sildenafil (Viagra) | 0.00E+00 | 6.67E-03 | NaN |
| Neuropathic Pain Signaling In Dorsal Horn Neurons | 0.00E+00 | 9.90E-03 | NaN |
| Endothelin-1 Signaling | 0.00E+00 | 5.21E-03 | NaN |
| Thrombin Signaling | 0.00E+00 | 4.44E-03 | NaN |
| Cardiac Hypertrophy Signaling | 0.00E+00 | 3.83E-03 | NaN |
| CDK5 Signaling | 0.00E+00 | 8.77E-03 | NaN |
| CREB Signaling in Neurons | 0.00E+00 | 4.95E-03 | NaN |
| Bladder Cancer Signaling | 0.00E+00 | 8.62E-03 | NaN |
| Non-Small Cell Lung Cancer Signaling | 0.00E+00 | 1.06E-02 | NaN |
| p70S6K Signaling | 0.00E+00 | 7.58E-03 | NaN |
| Colorectal Cancer Metastasis Signaling | 0.00E+00 | 3.69E-03 | NaN |
| G Beta Gamma Signaling | 0.00E+00 | 7.75E-03 | NaN |
| FAK Signaling | 0.00E+00 | 1.92E-03 | NaN |
| AMPK Signaling | 0.00E+00 | 4.10E-03 | NaN |
| RHOA Signaling | 0.00E+00 | 8.06E-03 | NaN |
| Role of Osteoblasts, Osteoclasts and Chondrocytes in Rheumatoid Arthritis | 0.00E+00 | 8.93E-03 | NaN |
| HER-2 Signaling in Breast Cancer | 0.00E+00 | 8.81E-03 | NaN |
| Altered T Cell and B Cell Signaling in Rheumatoid Arthritis | 0.00E+00 | 4.07E-03 | NaN |
| Atherosclerosis Signaling | 0.00E+00 | 7.63E-03 | NaN |
| Role of NFAT in Cardiac Hypertrophy | 0.00E+00 | 4.50E-03 | NaN |
| NUR77 Signaling in T Lymphocytes | 0.00E+00 | 1.95E-03 | NaN |
| Antiproliferative Role of TOB in T Cell Signaling | 0.00E+00 | 4.69E-03 | NaN |
| PI3K Signaling in B Lymphocytes | 0.00E+00 | 6.99E-03 | NaN |
| Signaling by Rho Family GTPases | 0.00E+00 | 3.73E-03 | NaN |
| Gap Junction Signaling | 0.00E+00 | 5.05E-03 | NaN |
| Telomerase Signaling | 0.00E+00 | 9.35E-03 | NaN |
| D-myo-inositol-5-phosphate Metabolism | 0.00E+00 | 1.01E-02 | NaN |
| D-myo-inositol (1,4,5,6)-Tetrakisphosphate Biosynthesis | 0.00E+00 | 1.10E-02 | NaN |
| Superpathway of Inositol Phosphate Compounds | 0.00E+00 | 8.44E-03 | NaN |
| D-myo-inositol (3,4,5,6)-tetrakisphosphate Biosynthesis | 0.00E+00 | 1.10E-02 | NaN |
| 3-phosphoinositide Degradation | 0.00E+00 | 1.03E-02 | NaN |
| 3-phosphoinositide Biosynthesis | 0.00E+00 | 9.62E-03 | NaN |
| Antioxidant Action of Vitamin C | 0.00E+00 | 8.93E-03 | NaN |
| GŒ±i Signaling | 0.00E+00 | 7.14E-03 | NaN |
| Granulocyte Adhesion and Diapedesis | 0.00E+00 | 5.29E-03 | NaN |
| Sperm Motility | 0.00E+00 | 3.92E-03 | NaN |
| Estrogen Receptor Signaling | 0.00E+00 | 7.33E-03 | NaN |
| ERK/MAPK Signaling | 0.00E+00 | 9.26E-03 | NaN |
| Nitric Oxide Signaling in the Cardiovascular System | 0.00E+00 | 8.33E-03 | NaN |
| Protein Ubiquitination Pathway | 0.00E+00 | 1.09E-02 | NaN |
| GABA Receptor Signaling | 0.00E+00 | 7.52E-03 | NaN |
| Integrin Signaling | 0.00E+00 | 4.69E-03 | NaN |
| IGF-1 Signaling | 0.00E+00 | 9.62E-03 | NaN |
| TGF-Œ≤ Signaling | 0.00E+00 | 1.04E-02 | NaN |
| Apoptosis Signaling | 0.00E+00 | 9.62E-03 | NaN |
| NF-Œ∫B Signaling | 0.00E+00 | 1.75E-03 | NaN |
| G-Protein Coupled Receptor Signaling | 0.00E+00 | 4.27E-03 | NaN |
| Autophagy | 0.00E+00 | 9.30E-03 | NaN |
| Sumoylation Pathway | 0.00E+00 | 9.71E-03 | NaN |
| Osteoarthritis Pathway | 0.00E+00 | 4.24E-03 | NaN |
| Neuroinflammation Signaling Pathway | 0.00E+00 | 3.15E-03 | NaN |
| Adrenomedullin signaling pathway | 0.00E+00 | 5.03E-03 | NaN |
| Endocannabinoid Cancer Inhibition Pathway | 0.00E+00 | 6.80E-03 | NaN |
| Cardiac Hypertrophy Signaling (Enhanced) | 0.00E+00 | 1.84E-03 | NaN |
| T Cell Exhaustion Signaling Pathway | 0.00E+00 | 1.77E-03 | NaN |
| Synaptogenesis Signaling Pathway | 0.00E+00 | 3.15E-03 | NaN |
| Semaphorin Neuronal Repulsive Signaling Pathway | 0.00E+00 | 6.62E-03 | NaN |
| Tumor Microenvironment Pathway | 0.00E+00 | 5.59E-03 | NaN |
| Role of MAPK Signaling in Promoting the Pathogenesis of Influenza | 0.00E+00 | 8.85E-03 | NaN |

**Supplemental Table 11. Peptides used in IFN-γ ELISPOT assay**

| **Virus** | **Antigen** | **Supplier** | **Catalog #** |
| --- | --- | --- | --- |
| DENV-3: Sleman/1978 | E (partial) | BEI | NR-511 |
| DENV-3: Philippines/H87/1956 | E (partial) | BEI | NR-9228 |
| DENV-3: Philippines/H87/1956 | NS1 | BEI | NR-2753 |
| DENV-3: Philippines/H87/1956 | NS3 | BEI | NR-2754 |
| DENV-3: Philippines/H87/1956 | NS5 | BEI | NR-4204 |

**Supplemental Table 12.** Antibodies used for flow cytometry

| **Antibody** | **Clone** | **Manufacturer** | **Catalog #** | **Lot #** | **Dilution** |
| --- | --- | --- | --- | --- | --- |
| CD38 PE-Cy7 | HIT2 | Biolegend | 303516 | B335356 | 1:50 |
| CD27 AF647 | O323 | Biolegend | 302812 | B243264 | 1:200 |
| CD56 BV510 | 5.1H11 | Biolegend | 362534 | B340656 | 1:50 |
| CD14 BV510 | M5E2 | Biolegend | 301841 | B332900 | 1:100 |
| CD3 BV510 | UCHT1 | Biolegend | 300449 | B349804 | 1:50 |
| CD19 BV785 | HIB19 | Biolegend | 302239 | B300756 | 1:200 |
